# Cost-effectiveness and budget impact of decentralising childhood tuberculosis diagnosis: a mathematical modelling study in six high tuberculosis incidence countries

**DOI:** 10.1101/2023.09.26.23295962

**Authors:** Marc d’Elbée, Martin Harker, Nyashadzaishe Mafirakureva, Mastula Nanfuka, Minh Huyen Ton Nu Nguyet, Jean-Voisin Taguebue, Raoul Moh, Celso Khosa, Ayeshatu Mustapha, Juliet Mwanga-Amumpere, Laurence Borand, Sylvie Kwedi Nolna, Eric Komena, Saniata Cumbe, Jacob Mugisha, Naome Natukunda, Tan Eang Mao, Jérôme Wittwer, Antoine Bénard, Tanguy Bernard, Hojoon Sohn, Maryline Bonnet, Eric Wobudeya, Olivier Marcy, Peter J. Dodd, TB-Speed Health Economics Study Group

## Abstract

**Background:** The burden of childhood tuberculosis remains high globally, largely due to under-diagnosis. Decentralising childhood tuberculosis diagnosis services to lower health system levels could improve case detection, but there is little empirically based evidence on cost-effectiveness or budget impact.

**Methods:** We assessed the cost-effectiveness and budget impact of decentralising a comprehensive diagnosis package for childhood tuberculosis to district hospitals (DH-focused) or primary health centres (PHC-focused) compared to standard of care (SOC) in Cambodia, Cameroon, Côte d’Ivoire, Mozambique, Sierra Leone, and Uganda (NCT04038632). A mathematical model was developed to assess the health and economic outcomes of the intervention from a health system perspective. Estimated outcomes were tuberculosis cases, deaths, disability-adjusted life years and incremental cost-effectiveness ratios (ICERs). We also calculated the budget impact of nationwide implementation.

**Findings:** For the DH-focused strategy versus SOC, ICERs ranged between $263 (Cambodia) and $342 (Côte d’Ivoire) per disability-adjusted life-year (DALY) averted. For the PHC-focused strategy versus SOC, ICERs ranged between $477 (Cambodia) and $599 (Côte d’Ivoire) per DALY averted. Results were sensitive to TB prevalence and the discount rate used. The additional costs of implementing the DH-focused strategy ranged between $13M (Cambodia) and $50M (Mozambique), and between $14M (Sierra Leone) and $135M (Uganda) for the PHC-focused strategy.

**Interpretation:** The DH-focused strategy may be cost-effective in some countries, depending on the cost-effectiveness threshold used for policy making. Either intervention would require substantial early investment.

**Funding:** Unitaid

## Research in context

### Evidence before the study

We searched the PubMed database using (tuberculosis) AND ((paediatric) OR (pediatric) OR (child*)) AND ((Costs and Cost Analysis) OR (“budget impact analysis”)) between January 2000 and February 2023, without language restrictions.

We found 16 articles assessing the cost-effectiveness of a broad range of interventions for child tuberculosis such as vaccine programs, the use of GeneXpert machines, systematic screening, directly observed therapy, short-course preventive therapy, addition of sampling (stool) method to national algorithms, isoniazid preventive therapy delivery, and contact tracing. We did not find any budget impact analysis on tuberculosis care intervention for children. A study published by Thompson et al in 2023 investigated the costs and cost-effectiveness of a decentralised molecular testing strategy for adult tuberculosis in Uganda. It did not include chest radiography as part of the diagnostic strategy. Overall, the authors recommended decentralised Xpert testing, however, the study addressed adult patients, and no budget impact analysis was conducted to inform scale-up.

In 2022, WHO recommended decentralised models of care to deliver tuberculosis services to children. The WHO guideline development group rated the overall certainty of evidence found through systematic review as “very low”, with no evidence on cost-effectiveness, and recognised that adequate investment was critical to enable the acceptability, equity and feasibility of decentralised approaches. To date, the TB-Speed Decentralization study is the closest assessment of the intervention recommended by the WHO.

### Added value of this study

Unlike all of the published studies, our intervention investigated a decentralised comprehensive package of care for children as developed by the TB-Speed project which includes systematic tuberculosis screening for all sick children <15 years, clinical evaluation, Xpert MTB/RIF Ultra-testing on respiratory and stool samples, and chest radiography for children with presumptive tuberculosis.

This study found that, compared to the standard of care, decentralising a comprehensive diagnosis package for childhood tuberculosis to district hospitals is potentially cost-effective from a health systems perspective while decentralising to primary health centres is unlikely to be cost-effective. Decentralisation would require substantial financial investment in the early implementation phase for equipment purchases.

### Implications of all the available evidence

Decentralisation of tuberculosis diagnostic services could be cost-effective in some settings with high prevalence of tuberculosis in children seeking healthcare. The main factors affecting cost-effectiveness are the level of decentralisation (district hospital versus primary health centre), local tuberculosis prevalence, and facility testing volumes. Substantial financial commitment is needed in the early implementation phase. Following WHO recommendations, countries should consider scaling up locally-adapted interventions to improve diagnosis of tuberculosis and other diseases using Xpert machines to improve cost-effectiveness, prioritising areas with highest tuberculosis prevalence, and include such plans when identifying domestic or donor sources of funding.

## Introduction

Tuberculosis mortality remains high in children globally,^1^ with 209,000 deaths estimated by the World Health Organization (WHO) for 1.1 million paediatric cases in 2021.^2^ Modelling suggests the majority (96%) of these deaths are occurring among children not receiving treatment for tuberculosis.^3^ In 2021, only 38.5% of childhood tuberculosis cases were reported to the WHO, largely because of substantial underdiagnosis which prevents children from receiving treatment.

Diagnosing tuberculosis in children is challenging largely due to difficulties in collecting expectorated sputum samples and insufficient bacteria in samples to test positive due to the paucibacillary nature of pulmonary tuberculosis in children.^4–6^ Alternative specimen collection methods such as induced sputum and gastric aspirate require equipment and experienced personnel, which are often lacking at primary health centre (PHC) level where most sick children seek care, and may be unavailable at district hospital (DH) level. Consequently, most children with presumptive tuberculosis (symptoms suggestive of tuberculosis) do not access appropriate diagnostic tests for tuberculosis, even when seen at DH level.^7^ Referrals introduce potential delays, risk losses to follow-up, and do not align with the ambition of providing patient-centred tuberculosis care.^8^

Developments in molecular diagnostics and novel sample collection procedures provide opportunities to decentralise diagnostic capacity for paediatric tuberculosis to the primary care level. While more costly than smear microscopy, the higher sensitivity, robustness and low training requirements of the WHO-endorsed GeneXpert-operated rapid molecular diagnostic assays for tuberculosis^9^ allow their deployment at PHC level. Stool samples can be collected in young children regardless of setting or equipment compared to respiratory samples and can be used to identify *Mycobacterium tuberculosis* using Xpert MTB/RIF.^10,11^ Nasopharyngeal aspirates (NPA) are easier to collect than gastric aspirate or induced sputum sampling, and in combination with stool samples can provide similar sensitivity to Xpert MTB/RIF testing on two gastric aspirates or induced sputa.^10^ The WHO has now recommended these sample collection methods with Xpert MTB/RIF Ultra, the latest generation of molecular tests, for paediatric tuberculosis diagnosis.^9^

The recent revision to the WHO guidelines for child and adolescent tuberculosis recommended decentralised models of care to notably increase case detection in children.^9^ However, this recommendation was provisional due to the low quality of evidence, including evidence on cost-effectiveness, for such approaches. Few published studies evaluate models of care for paediatric tuberculosis, and even fewer include economic evaluation; none to our knowledge has included a budget impact analysis. For many countries with limited resources and high tuberculosis incidence, objectively weighing trade-offs between policy options and considering their affordability is crucial, and is explicitly required when applying for donor support including Global Fund.

The TB-Speed Decentralization study (NCT04038632) used a pre-/post-intervention approach to assess the impact on tuberculosis case detection of decentralising a comprehensive childhood tuberculosis diagnosis package at DH or PHC level.^12^ The package included systematic outpatient screening, Xpert MTB/RIF Ultra on stool and NPA samples for those with presumptive tuberculosis, enhanced healthcare worker training and clinical mentoring to improve clinical skills, and digital chest X-ray at DH level. The study was conducted in 12 DHs and 47 PHCs across six high tuberculosis incidence countries: Cambodia, Cameroon, Côte d’Ivoire, Mozambique, Sierra Leone, and Uganda. In each country, two rural or semi-urban districts were randomly allocated to either a PHC-focused or DH-focused decentralisation strategy. In the DH-focused strategy, children with presumptive tuberculosis at PHC level were referred to DH for diagnosis; in the PHC-focused strategy, diagnostic evaluations were performed at PHC, except chest X-ray done at DH if required. Baseline ‘pre’ data comprised 9 months of retrospective data and at least 3 months of prospective data. Comparator ‘post’ data were collected for 12 months after introducing the intervention.

In this analysis we sought to fill the evidence gaps on the economic evaluation of real-world strategies for improving paediatric tuberculosis care by undertaking a cost-effectiveness and budget impact analysis of the TB-Speed Decentralization study^12^. We used empirical data on costs and the cascades of tuberculosis care (screening, clinical assessment, microbiological/radiological testing, and treatment) from the study with mathematical models of patient pathways to compare the health and cost consequences of the different models of care.

## Methods

### Patient pathways

Conceptual models were developed through iterative consultation with country experts and the TB-Speed Decentralization study team to represent detailed patient care pathways for three comparator arms: a standard of care (SOC), the DH-focused strategy and the PHC-focused strategy (**Figure 1** and **Appendix**). Diagrammatic representations of patient pathways formed the basis of the decision-analytic model structure; accompanying narrative descriptions of activities at each stage informed quantification of resource use. We sought to develop pathways that were general enough to include common elements across all included countries. The SOC pathway represented expert consensus on typical care available in high tuberculosis incidence countries, informed by the baseline facility assessment from the included countries before the start of the TB-Speed Decentralization intervention. DH-focused and PHC-focused pathways reflected the TB-Speed Decentralization protocol, to represent the expected patient pathway if these interventions were implemented widely across the target countries.

**Figure 1.**
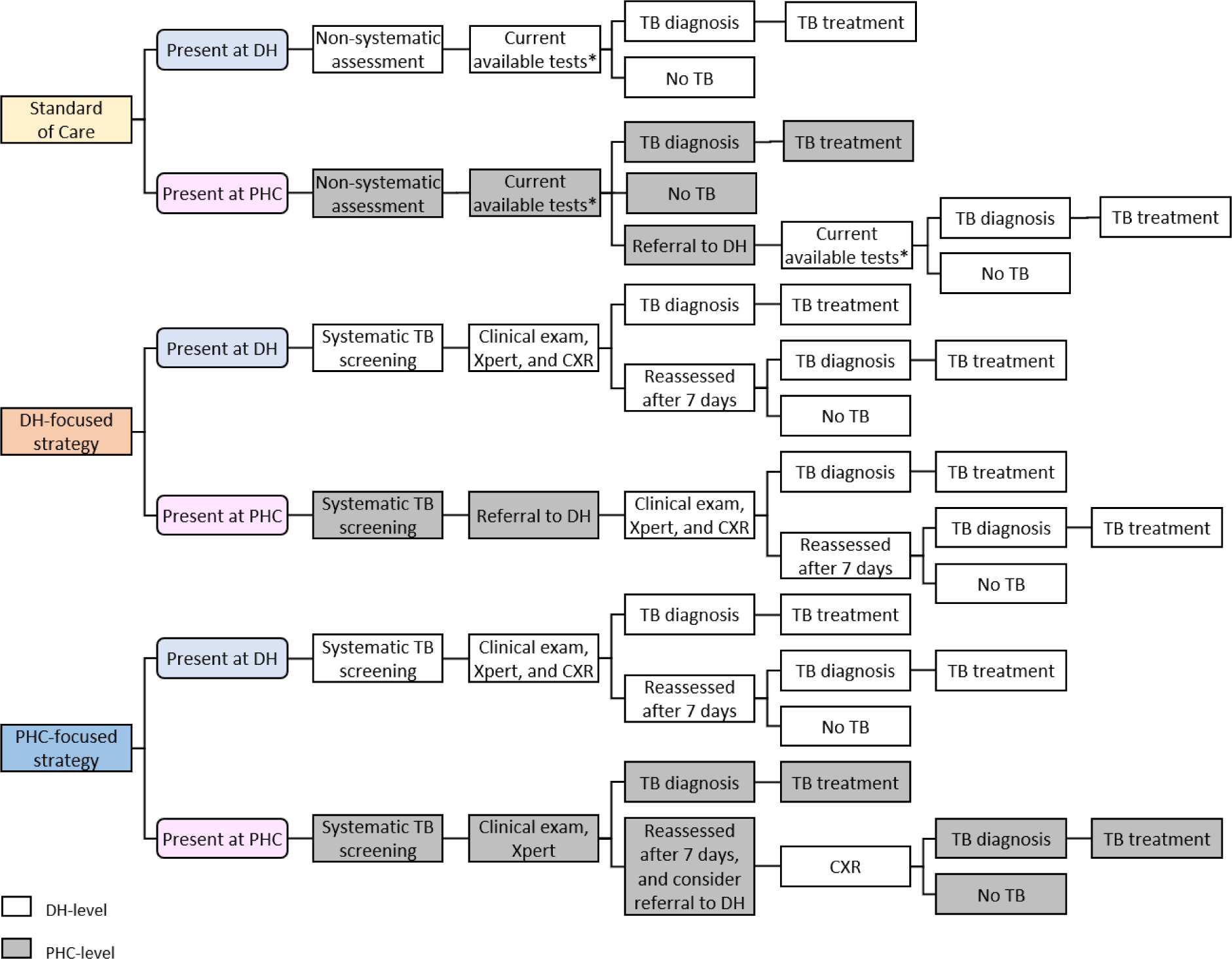
Simplified patient care pathways for the diagnosis and treatment of tuberculosis in children *Clinical exam, and, for a proportion of patients, Xpert on sputum or gastric aspirate, smear microscopy and CXR (in DH) TB: tuberculosis, DH: district hospital, PHC: primary health centre, CXR: chest X-ray

### Costing approach

#### Cost data collection

Cost data collection tools were adapted from the Value TB costing tool suite developed in conjunction with the Global Health Cost Consortium (GHCC), with reference to the GHCC/WHO guidance ‘Costing Guidelines for Tuberculosis Interventions’.^13^

Labour costs were sourced from national pay scales and project accounts, medications from the Stop TB partnership Global Drug Facility catalogue,^14^ consumables including materials for diagnostic tests, staff training, and equipment from project accounts, and hospitalisation cost of an inpatient bed day from the WHO-CHOICE unit cost estimates for service delivery.^15^ As this intervention did not change facility infrastructure, we excluded facility-associated overhead costs from the analysis. More information on the scope of the costing can be found in **Appendix**.

Routine health facility-level aggregated data on patient numbers were collected by field research assistants from outpatient, tuberculosis, and laboratory registers during the observation, preparatory and intervention phases.

To calculate the proportionate use of major equipment such as X-ray or GeneXpert machines we used the expected lifespan and annual number of uses of each item. These data were obtained through key informants at MSF-Logistique (https://www.msflogistique.org/) for expected lifespan and from laboratory managers for number of uses over the course of 12 months at PHCs and at DHs.

#### Time and motion study

To value the contribution of labour we conducted a time and motion study to estimate the length of time that staff spent on each patient care task under TB-Speed interventions. Timesheets were used to record the length of tuberculosis-related consultations, appointments and diagnostic tests. Healthcare workers were invited to take part voluntarily and no personal data were collected, only the site name and the role of the healthcare worker. On the self-completed timesheets, participants recorded all activities related to TB-Speed patients and the length of their time they spent on each activity. Participants included doctors, clinical officers, nurses, laboratory technicians and radiographers, at DH and PHC levels in all countries, with 179 participants in total.

#### Cost analysis

Unit costs were estimated using an ingredient-based costing approach, in which the expected resources required for each child enrolled into the study were listed, costed, and summed to estimate direct health service utilisation and cost varying by patient characteristics and the route taken on the pathway. Services (number and type of diagnostic and treatment procedures, materials, laboratory investigations and medications) provided to patients were valued by multiplying the quantities required by their unit costs. The value of all time spent by staff for each patient was estimated as the product of ‘hours spent’ and ‘hourly labour costs’. Costs were estimated in 2021 US dollars (USD), using a discount rate of 3% for the annualisation of the economic costs of equipment following guidelines.^16,17^

### Modelling approach

A decision analytic mathematical model was developed in R software to assess the clinical benefits, cost-effectiveness, and budget impact of the intervention from a health system perspective. The attributes of children flowing through the tree were: age (0-4 years or 5-14 years); HIV and antiretroviral treatment status (each positive or negative); and true tuberculosis status (bacteriologically-confirmed tuberculosis, bacteriologically-unconfirmed tuberculosis, not tuberculosis). The attribute ‘bacteriologically confirmed tuberculosis’ refers to tuberculosis that would be bacteriologically positive under ideal circumstances and with all samples available.

The probabilities of following each route through the tree depended on attributes, and were parameterized using a mixture of literature, TB-Speed Decentralization study data, and expert opinion. Literature was used to parametrize the accuracy of diagnostic algorithms and availability of samples. Study data on child characteristics, level of presentation, and the care cascade were used to calibrate tuberculosis prevalence, the level of initial care seeking (given true tuberculosis status), and the likelihoods of being assessed and being considered to have presumptive tuberculosis. Expert opinion was used to inform unobserved features of typical care such as referral loss to follow-up and the frequency with which certain diagnostic tests were used under standard of care. Modelling of the cascade of care and intervention effect was based on calibration of unobserved parameters to meta-analytic summaries, representing a typical cascade. We undertook sensitivity analyses exploring the influence of tuberculosis prevalence, assumed constant across countries in the model, and of the discount rate applied. See **Appendix** for more details.

Country-specific unit costs associated with resource use at each step of care were accumulated to produce total mean costs. Health benefits for those with tuberculosis were modelled with a previously published approach that used case-fatality ratio from systematic literature reviews to quantify mortality reductions from treating more tuberculosis.^3^ Country-specific life expectancy from United Nations estimates was used to calculate the mean life-years lost over a lifetime horizon (with and without 3% discounting). We disregarded the contribution of morbidity to disability adjusted life-years (DALYs). All results were calculated using a probabilistic sensitivity analysis with 1,000 replicates.

We report the total and incremental (to SOC) number of children treated for tuberculosis, costs, number of deaths and deaths averted, number of DALYs and DALYs averted, per 100,000 children presenting as outpatients, and the incremental cost-effectiveness ratios (ICERs) for both interventions in each country. ICERs were compared to various options for cost-effectiveness thresholds (presented as a range) in each country to assess potential cost-effectiveness.^18,19^ We complied with the Consolidated Health Economic Evaluation Reporting Standards (CHEERS 2022) reporting guidelines (**Appendix**).^20^

To project the 5-year (2022-2026) budget impact of adopting these interventions nationally in each country, we fitted a cost function in Excel using parameters related to costs of equipment and its delivery, installation and maintenance, training, supplies, and personnel costs. Input quantities were the national number of DHs, PHCs, eligible healthcare workers, and pre-intervention (2019) annual rate of tuberculosis notification in children 0-14 years old from the WHO Global TB Programme. See **Appendix** for additional details. Costs were combined with the scale of deployment to generate the required implementation budget in each year for each country. We assumed a 3-year period (2022-2024) to provide facilities with equipment, to train all eligible staff, and rollout supply chain delivery to all sites. A sensitivity analysis is presented in **Appendix**.

A health economic analysis plan was developed jointly by economists, modellers, international and country investigators. The economic analyses were nested within the main study protocol approved by the WHO Ethical Review Committee, Inserm’s ethics review committee (IRB00003888), as well as all national ethics committees prior to the start of the study (See **Appendix**).

## Results

Overall, the modelled cascade of care data showed that the tuberculosis screening rate among children presenting to health facilities increased from 21% (SOC) to 89% (DH-focused and PHC-focused), the rate of children identified with presumptive tuberculosis among screened children remained constant (3%), and the rate of children initiated on treatment among the children identified with presumptive tuberculosis increased from 12% (SOC) to 18% (DH- focused) and 14% (PHC-focused) (**Figure 2**). Results were similar between 0-4 and 5-14 years age groups (**Appendix**).

**Figure 2.**
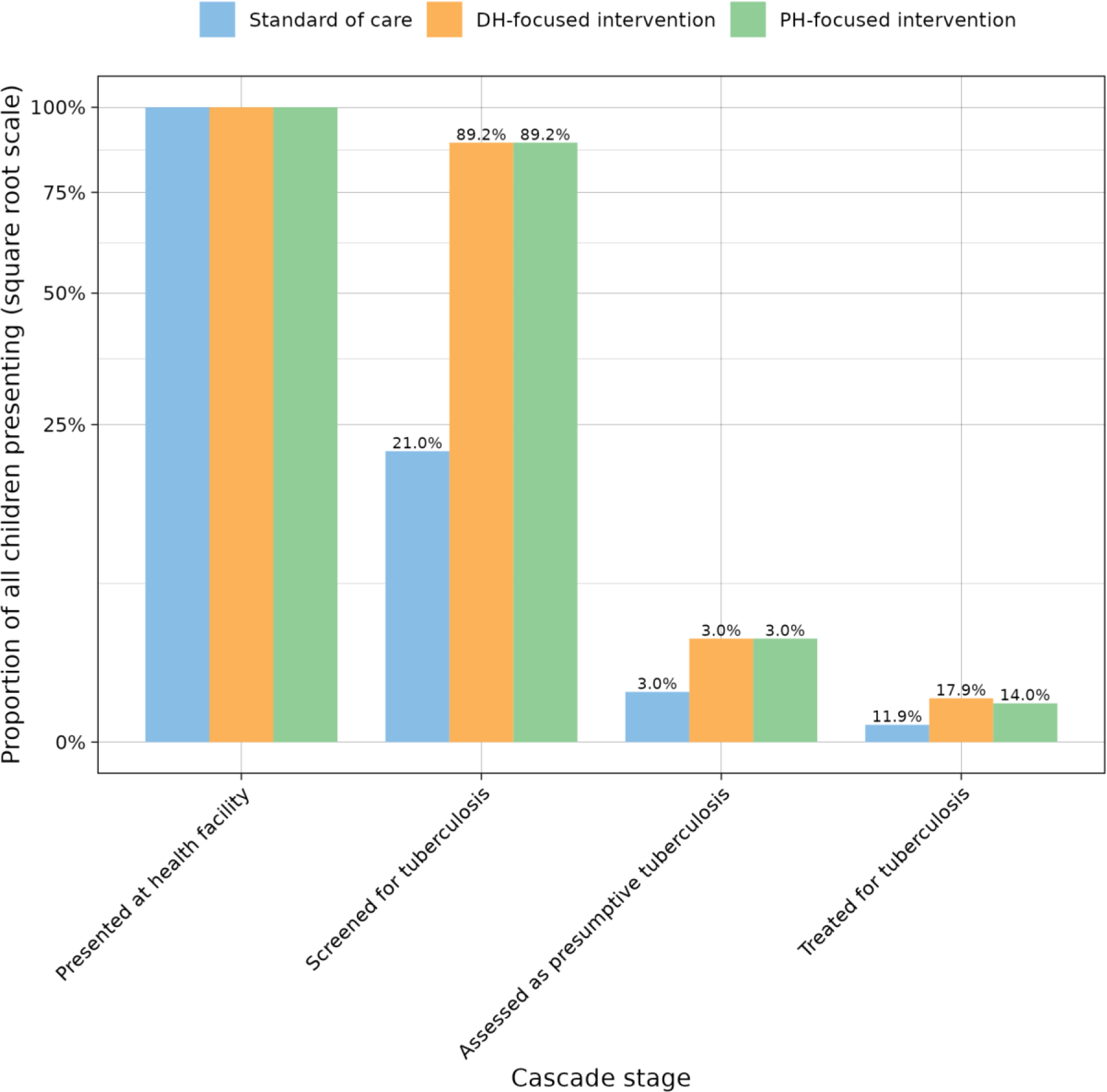
Screening to diagnosis cascades during the intervention period by decentralisation approach. Percentages are bar height as a proportion of the corresponding bar at the previous stage.

Results from the cost analysis and the time and motion study, as well as costs per child treated disaggregated by arm, cost category, and tuberculosis care stage are presented in the **Appendix**. Compared to 74 (95% uncertainty interval [UI]: 6 to 257) children treated for tuberculosis in the SOC (per 100,000 presenting), we estimated 476 (95% UI: 104 to 1,210) treated in the DH- focused arm and 373 (95% UI: 80 to 925) treated in the PHC-focused arm (**Table 1**). Compared with the SOC at 44 (6 to 131) deaths per 100,000 presenting, the number of deaths averted was 21 (2 to 69) and 18 (2 to 57), for the DH-focused and PHC-focused interventions, respectively. For the DH-focused intervention costs per 100,000 increased by between $158K ($41-357K) in Cambodia and $190K ($54-416K) in Côte d’Ivoire, while DALYs averted ranged from 547 (54-1,754) in Sierra Leone to 600 (59-1,916) in Cambodia.

**Table 1.**
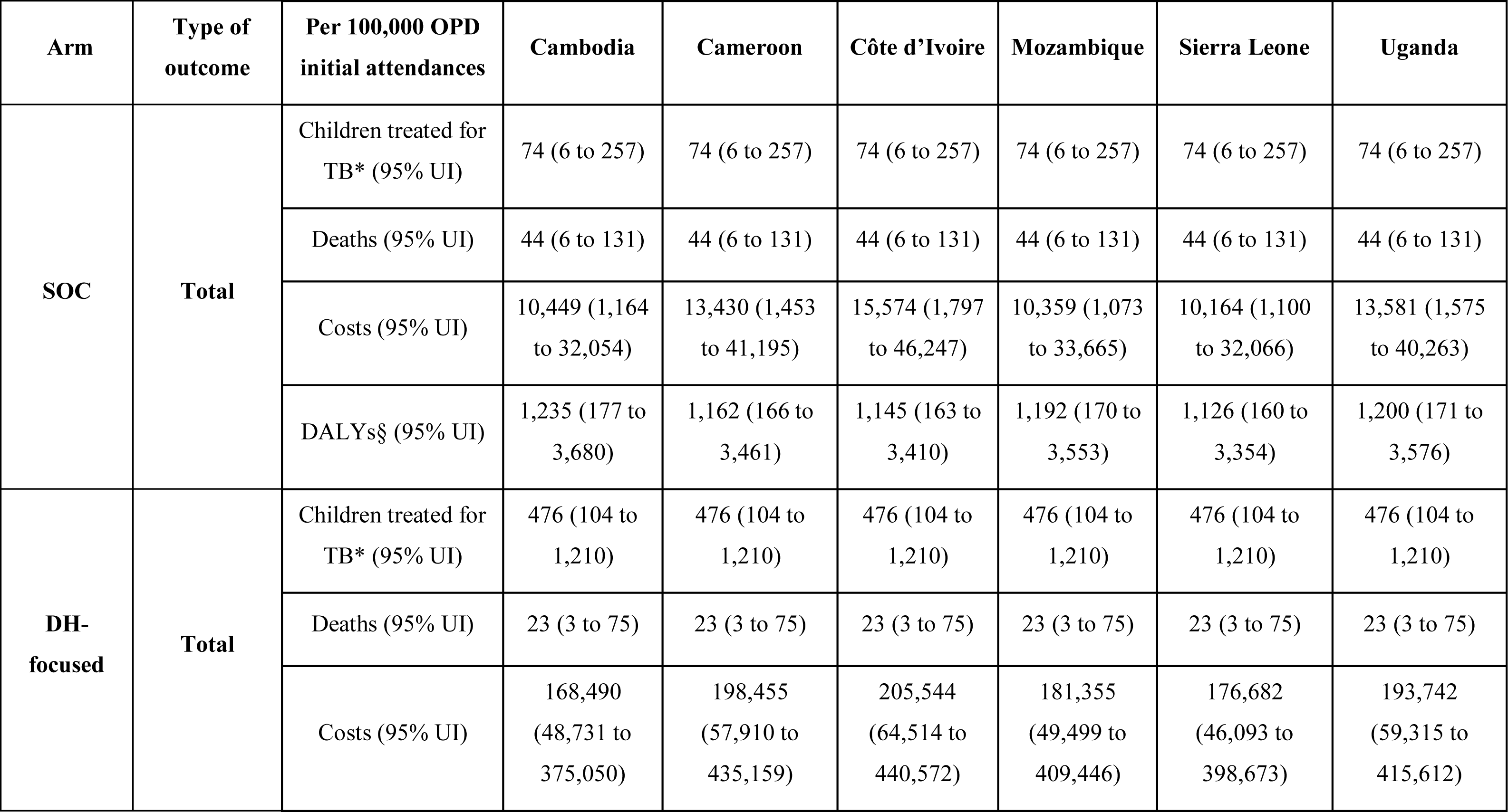
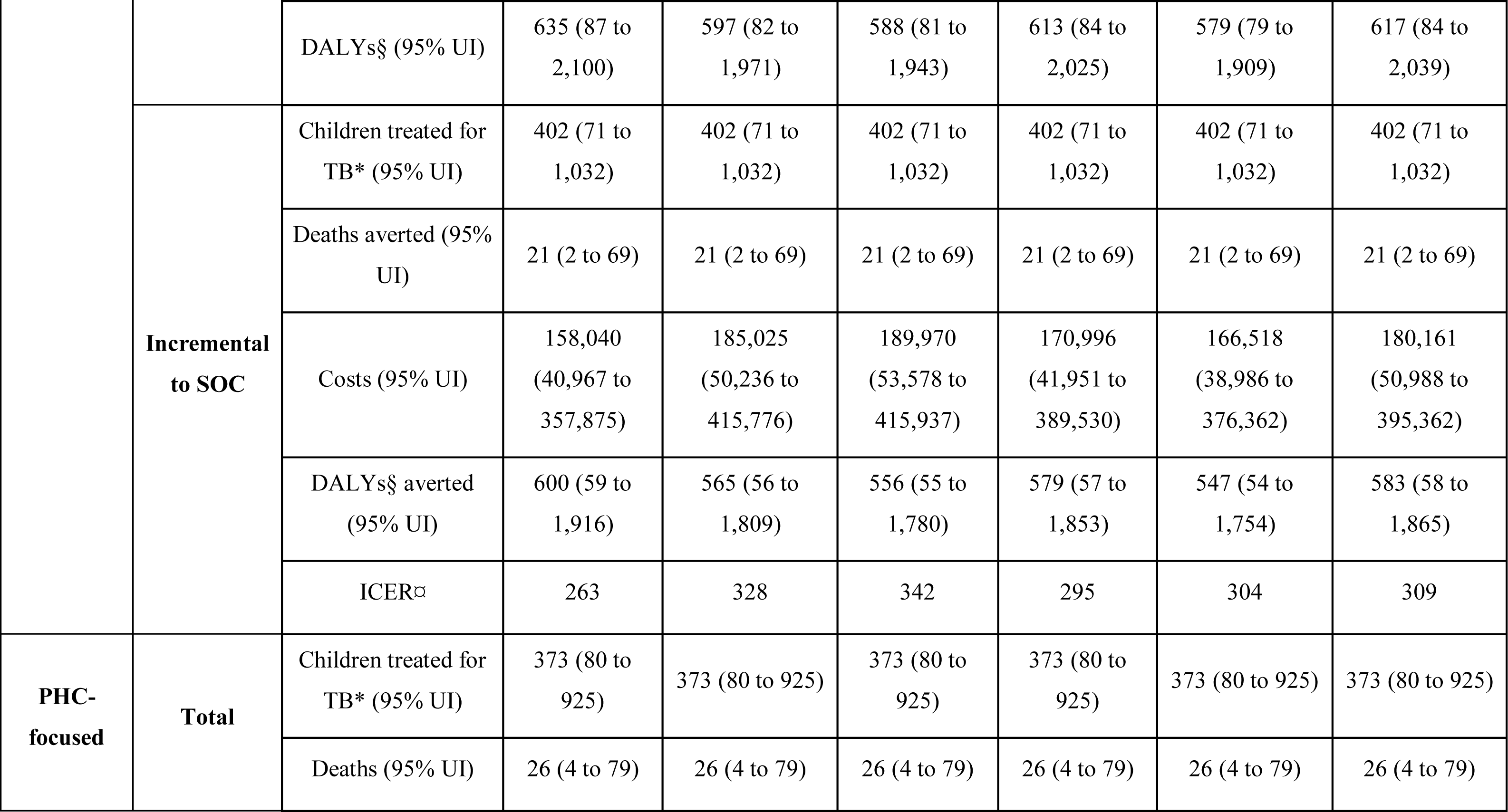
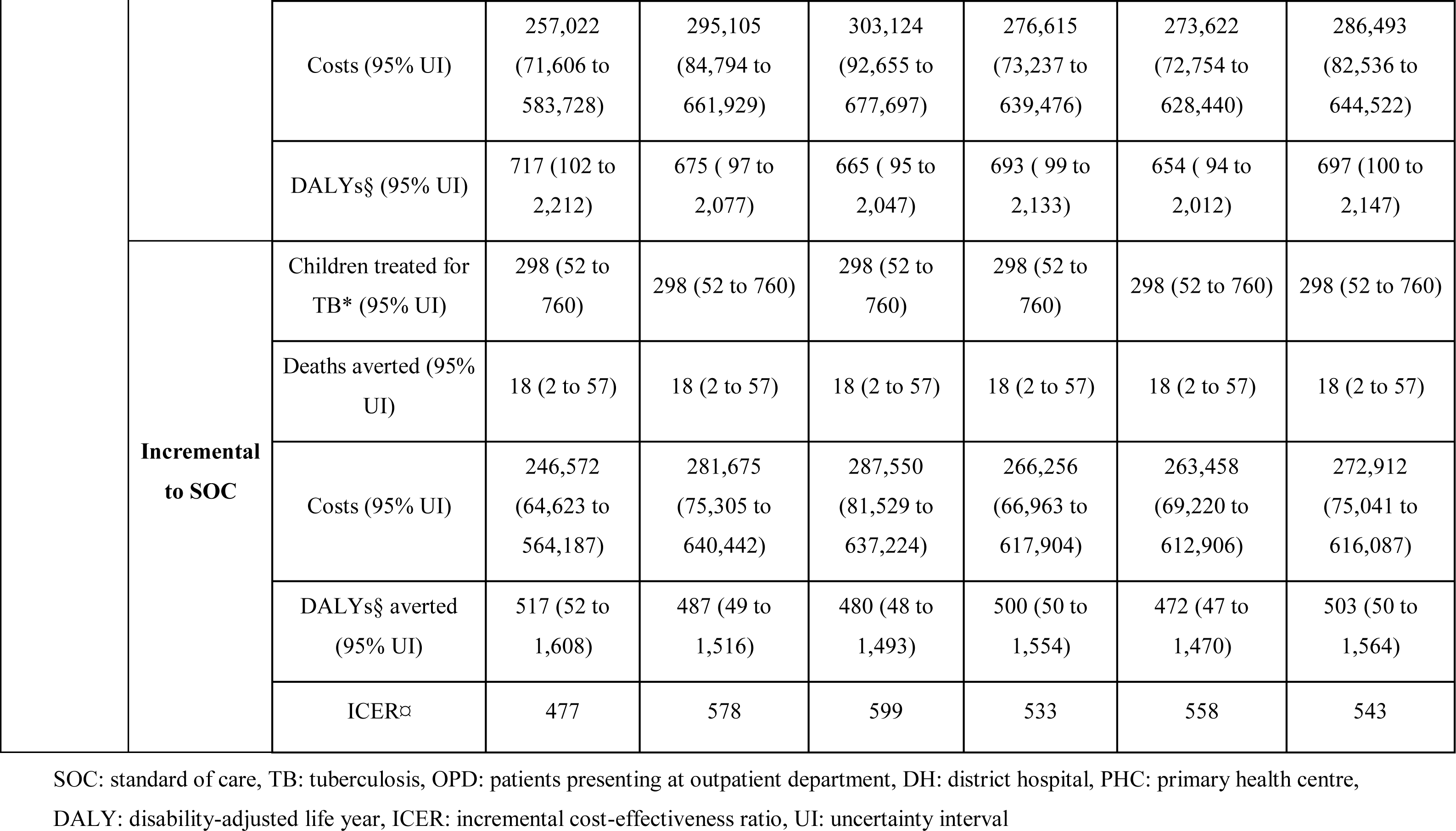
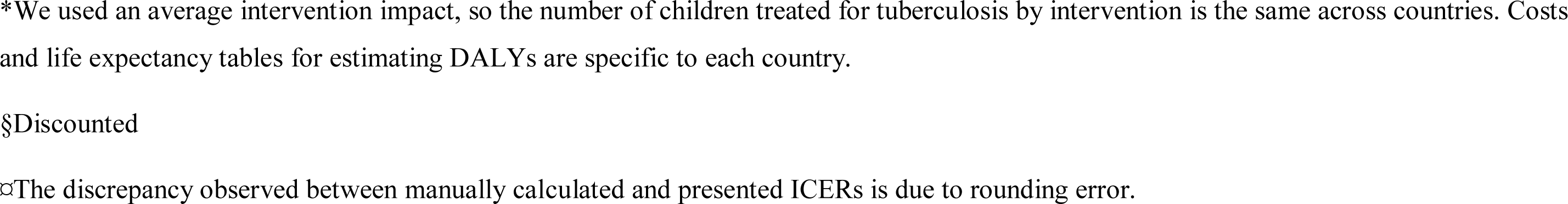
Health impact, costs and cost-effectiveness.

| Arm | Type of outcome | Per 100,000 OPD initial attendances | Cambodia | Cameroon | Côte d'Ivoire | Mozambique | Sierra Leone | Uganda |
| --- | --- | --- | --- | --- | --- | --- | --- | --- |
| **SOC** | **Total** | Children treated for TB* (95% UI) | 74 (6 to 257) | 74 (6 to 257) | 74 (6 to 257) | 74 (6 to 257) | 74 (6 to 257) | 74 (6 to 257) |
|  |  | Deaths (95% UI) | 44 (6 to 131) | 44 (6 to 131) | 44 (6 to 131) | 44 (6 to 131) | 44 (6 to 131) | 44 (6 to 131) |
|  |  | Costs (95% UI) | 10,449 (1,164 to 32,054) | 13,430 (1,453 to 41,195) | 15,574 (1,797 to 46,247) | 10,359 (1,073 to 33,665) | 10,164 (1,100 to 32,066) | 13,581 (1,575 to 40,263) |
|  |  | DALYs§ (95% UI) | 1,235 (177 to 3,680) | 1,162 (166 to 3,461) | 1,145 (163 to 3,410) | 1,192 (170 to 3,553) | 1,126 (160 to 3,354) | 1,200 (171 to 3,576) |
| **DH-focused** | **Total** | Children treated for TB* (95% UI) | 476 (104 to 1,210) | 476 (104 to 1,210) | 476 (104 to 1,210) | 476 (104 to 1,210) | 476 (104 to 1,210) | 476 (104 to 1,210) |
|  |  | Deaths (95% UI) | 23 (3 to 75) | 23 (3 to 75) | 23 (3 to 75) | 23 (3 to 75) | 23 (3 to 75) | 23 (3 to 75) |
|  |  | Costs (95% UI) | 168,490 (48,731 to 375,050) | 198,455 (57,910 to 435,159) | 205,544 (64,514 to 440,572) | 181,355 (49,499 to 409,446) | 176,682 (46,093 to 398,673) | 193,742 (59,315 to 415,612) |
|  |  | DALYs§ (95% UI) | 635 (87 to 2,100) | 597 (82 to 1,971) | 588 (81 to 1,943) | 613 (84 to 2,025) | 579 (79 to 1,909) | 617 (84 to 2,039) |
|  | **Incremental to SOC** | Children treated for TB* (95% UI) | 402 (71 to 1,032) | 402 (71 to 1,032) | 402 (71 to 1,032) | 402 (71 to 1,032) | 402 (71 to 1,032) | 402 (71 to 1,032) |
|  |  | Deaths averted (95% UI) | 21 (2 to 69) | 21 (2 to 69) | 21 (2 to 69) | 21 (2 to 69) | 21 (2 to 69) | 21 (2 to 69) |
|  |  | Costs (95% UI) | 158,040 (40,967 to 357,875) | 185,025 (50,236 to 415,776) | 189,970 (53,578 to 415,937) | 170,996 (41,951 to 389,530) | 166,518 (38,986 to 376,362) | 180,161 (50,988 to 395,362) |
|  |  | DALYs§ averted (95% UI) | 600 (59 to 1,916) | 565 (56 to 1,809) | 556 (55 to 1,780) | 579 (57 to 1,853) | 547 (54 to 1,754) | 583 (58 to 1,865) |
|  |  | ICER☒ | 263 | 328 | 342 | 295 | 304 | 309 |
| **PHC-focused** | **Total** | Children treated for TB* (95% UI) | 373 (80 to 925) | 373 (80 to 925) | 373 (80 to 925) | 373 (80 to 925) | 373 (80 to 925) | 373 (80 to 925) |
|  |  | Deaths (95% UI) | 26 (4 to 79) | 26 (4 to 79) | 26 (4 to 79) | 26 (4 to 79) | 26 (4 to 79) | 26 (4 to 79) |
|  |  | Costs (95% UI) | 257,022<br>(71,606 to<br>583,728) | 295,105<br>(84,794 to<br>661,929) | 303,124<br>(92,655 to<br>677,697) | 276,615<br>(73,237 to<br>639,476) | 273,622<br>(72,754 to<br>628,440) | 286,493<br>(82,536 to<br>644,522) |
|  |  | DALYs§ (95% UI) | 717 (102 to<br>2,212) | 675 ( 97 to<br>2,077) | 665 ( 95 to<br>2,047) | 693 ( 99 to<br>2,133) | 654 ( 94 to<br>2,012) | 697 (100 to<br>2,147) |
|  | **Incremental**<br>**to SOC** | Children treated for<br>TB* (95% UI) | 298 (52 to<br>760) | 298 (52 to 760) | 298 (52 to<br>760) | 298 (52 to<br>760) | 298 (52 to 760) | 298 (52 to 760) |
|  |  | Deaths averted (95%<br>UI) | 18 (2 to 57) | 18 (2 to 57) | 18 (2 to 57) | 18 (2 to 57) | 18 (2 to 57) | 18 (2 to 57) |
|  |  | Costs (95% UI) | 246,572<br>(64,623 to<br>564,187) | 281,675<br>(75,305 to<br>640,442) | 287,550<br>(81,529 to<br>637,224) | 266,256<br>(66,963 to<br>617,904) | 263,458<br>(69,220 to<br>612,906) | 272,912<br>(75,041 to<br>616,087) |
|  |  | DALYs§ averted<br>(95% UI) | 517 (52 to<br>1,608) | 487 (49 to<br>1,516) | 480 (48 to<br>1,493) | 500 (50 to<br>1,554) | 472 (47 to<br>1,470) | 503 (50 to<br>1,564) |
|  |  | ICERα | 477 | 578 | 599 | 533 | 558 | 543 |

SOC: standard of care, TB: tuberculosis, OPD: patients presenting at outpatient department, DH: district hospital, PHC: primary health centre, DALY: disability-adjusted life year, ICER: incremental cost-effectiveness ratio, UI: uncertainty interval
\*We used an average intervention impact, so the number of children treated for tuberculosis by intervention is the same across countries. Costs and life expectancy tables for estimating DALYs are specific to each country.
§Discounted
⌘The discrepancy observed between manually calculated and presented ICERs is due to rounding error.

The ICERs compared with the SOC ranged between $263 (Cambodia) and $342 (Côte d’Ivoire) per DALY averted for the DH-focused strategy, and between $477 (Cambodia) and $599 (Côte d’Ivoire) per DALY averted for the PHC-focused strategy. The DH-focused strategy dominated the PHC-focused strategy, being less expensive and more effective. Sensitivity analysis showed that both paediatric tuberculosis prevalence and discount rate applied on life years had a strong impact on the estimated ICERs (**Appendix**). At a constant TB prevalence of 200/100,000 across age groups, applying no discount rate (0% instead of 3%) led to ICERs decreasing by 57%- 60% across countries and strategies. Increasing TB prevalence from 200/100,000 to 500/100,000 (with fixed 3% discounting) led to ICERs decreasing by 55%-56%.

The cost-effectiveness acceptability curves (**Figure 3** and **Appendix**) show the decision uncertainty surrounding the adoption of the strategies, depending on the cost-effectiveness thresholds which decision-makers may select in each country. Using estimated thresholds from Ochalek et al.^18^, the highest estimated probabilities of the DH-focused strategy being cost-effective compared to SOC were for Cambodia (40%-69%) and Côte d’Ivoire (49%-65%) and the lowest for Sierra Leone (0%-1%). For the PHC-focused strategy versus SOC, the highest probabilities were for Cambodia (7%-27%) and Côte d’Ivoire (11%-26%) and the lowest for Cameroon, Sierra Leone and Uganda (<1%).

**Figure 3.**
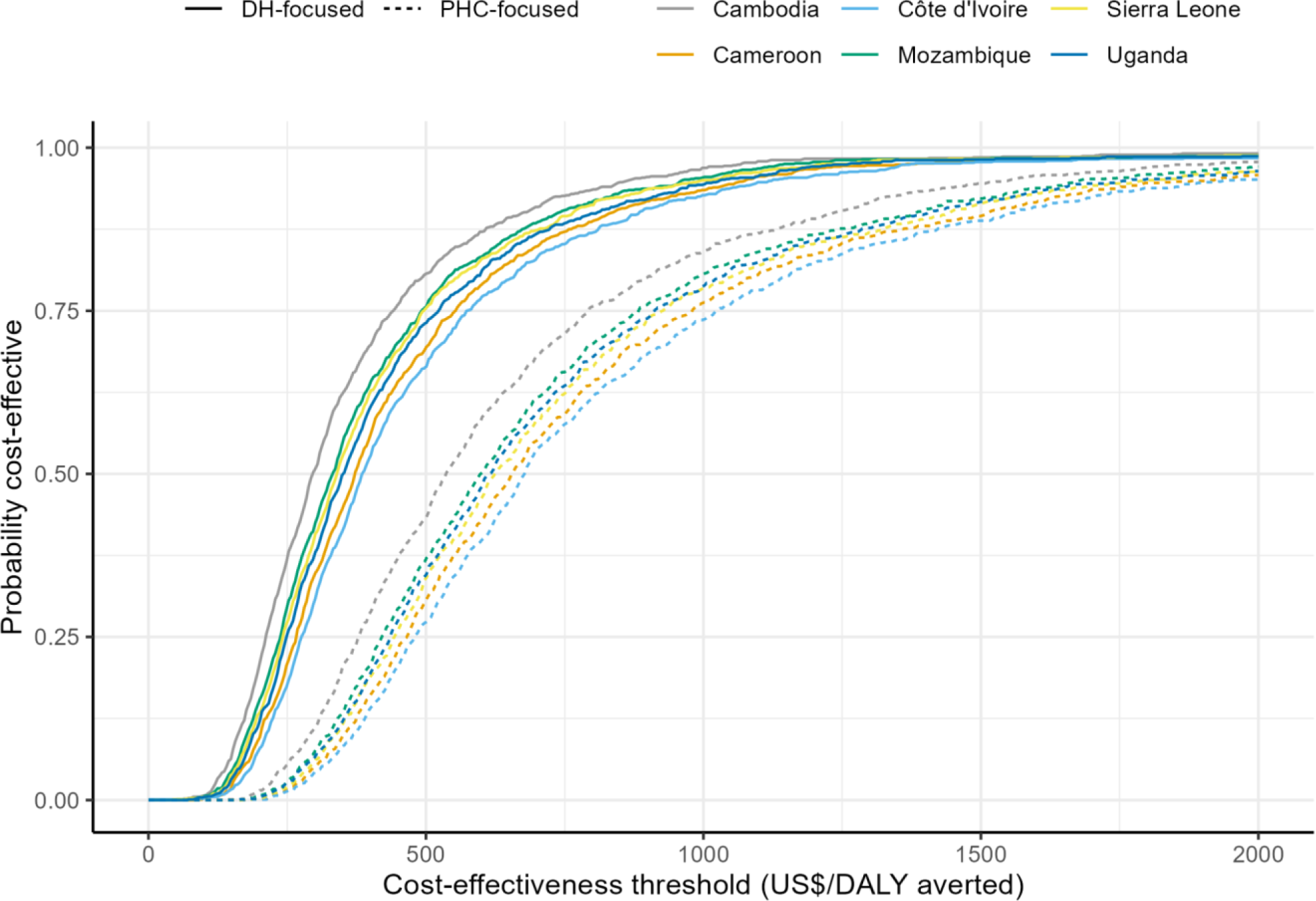
Cost-effectiveness acceptability curves by country for the district hospital-focused and primary health centre-focused strategies, each compared to the standard of care (in US$ per DALY averted). DALY: disability-adjusted life year

Additional costs of implementing the decentralisation intervention at DH level over five years were estimated between $13M in Cambodia and Sierra Leone and $50M in Mozambique, whereas decentralising to PHC level would cost between $14M in Sierra Leone and $135M in Uganda (**Table 2**). Countries with higher numbers of health facilities to equip (Cambodia, Mozambique, Sierra Leone) and/or higher tuberculosis notification rates (Cambodia, Mozambique, Sierra Leone, Uganda) had higher implementation costs.

**Table 2.**
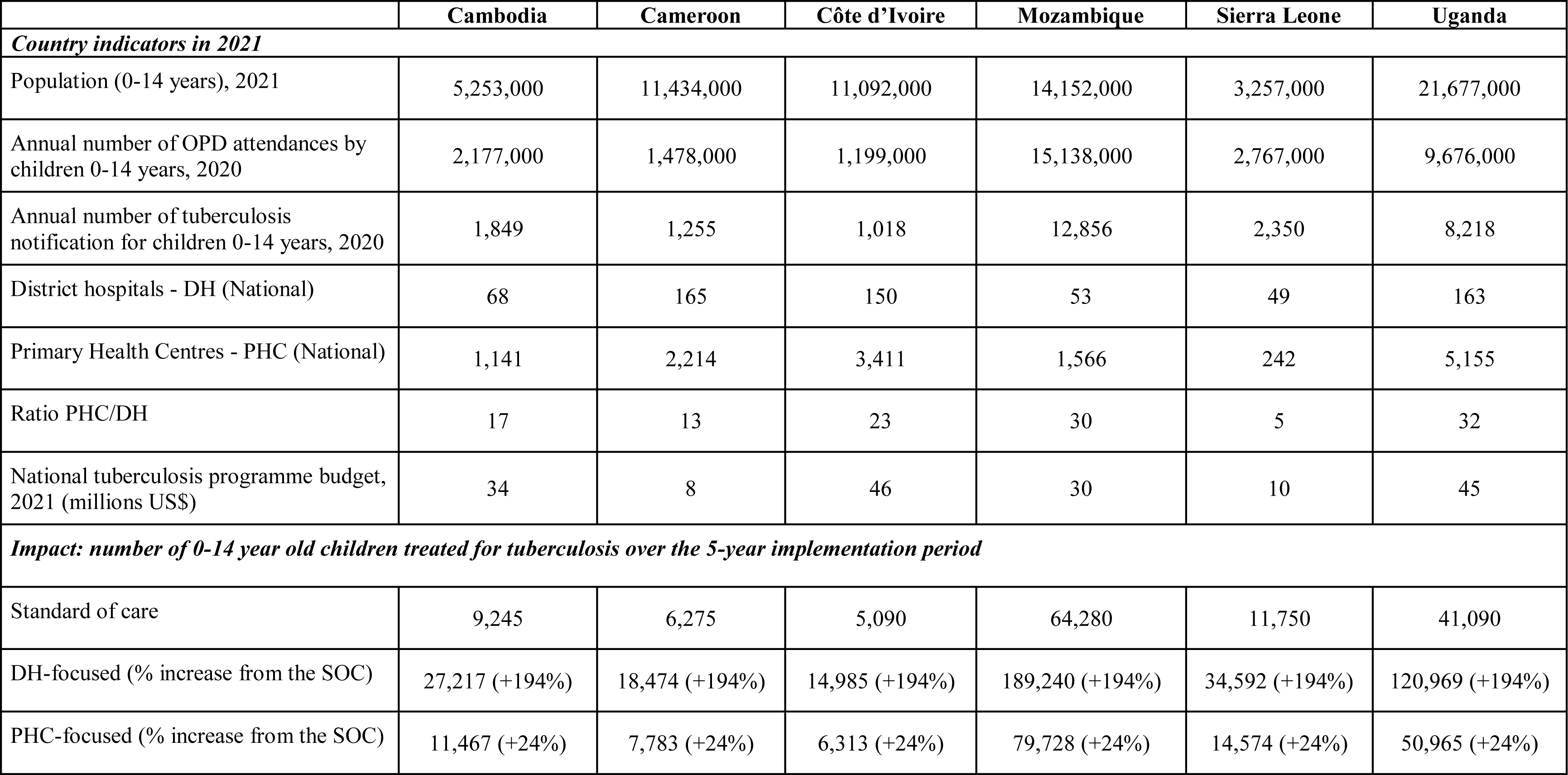
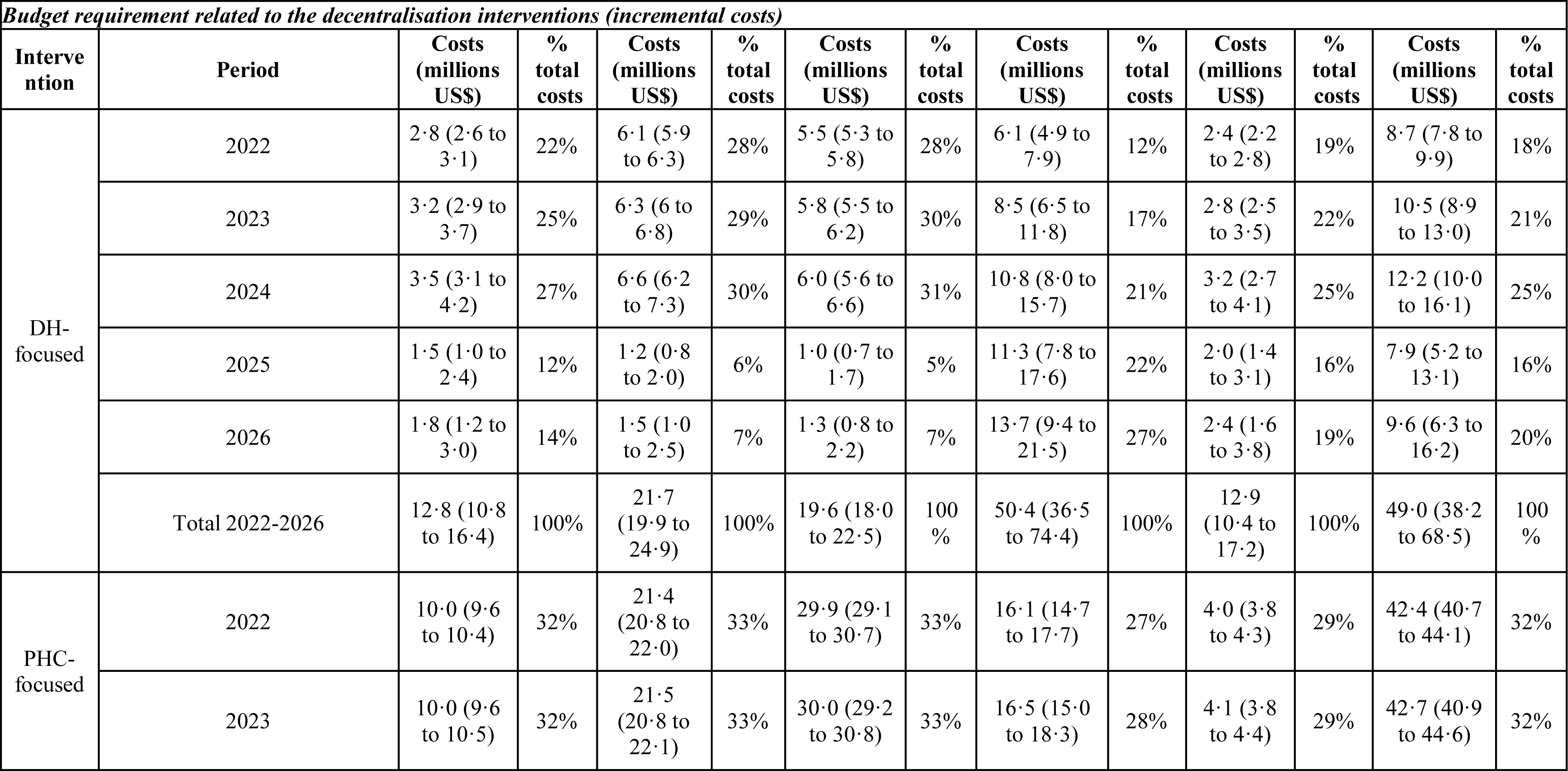
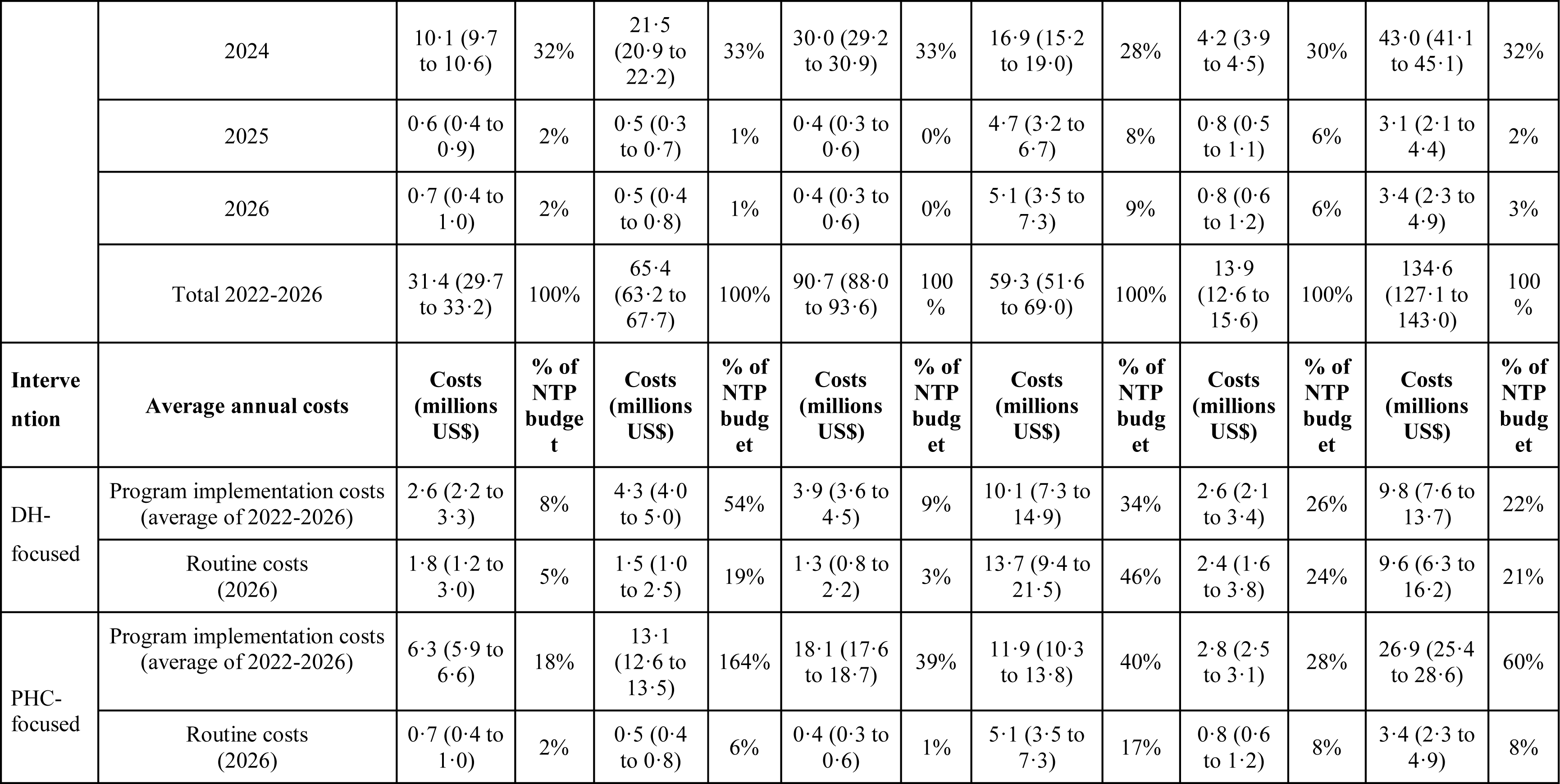
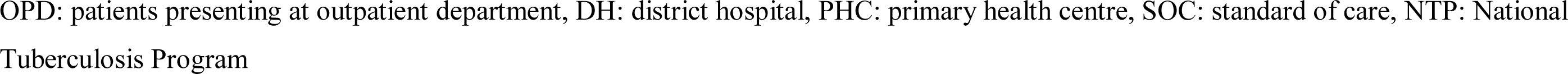
Projected budget impact of adopting the decentralisation intervention by strategy and by country for years 2022 to 2026. Costs are in 2021 US$.

|  | **Cambodia** | **Cameroon** | **Côte d'Ivoire** | **Mozambique** | **Sierra Leone** | **Uganda** |
| --- | --- | --- | --- | --- | --- | --- |
| ***Country indicators in 2021*** |  |  |  |  |  |  |
| Population (0-14 years), 2021 | 5,253,000 | 11,434,000 | 11,092,000 | 14,152,000 | 3,257,000 | 21,677,000 |
| Annual number of OPD attendances by children 0-14 years, 2020 | 2,177,000 | 1,478,000 | 1,199,000 | 15,138,000 | 2,767,000 | 9,676,000 |
| Annual number of tuberculosis notification for children 0-14 years, 2020 | 1,849 | 1,255 | 1,018 | 12,856 | 2,350 | 8,218 |
| District hospitals - DH (National) | 68 | 165 | 150 | 53 | 49 | 163 |
| Primary Health Centres - PHC (National) | 1,141 | 2,214 | 3,411 | 1,566 | 242 | 5,155 |
| Ratio PHC/DH | 17 | 13 | 23 | 30 | 5 | 32 |
| National tuberculosis programme budget, 2021 (millions US\$) | 34 | 8 | 46 | 30 | 10 | 45 |
| ***Impact: number of 0-14 year old children treated for tuberculosis over the 5-year implementation period*** |  |  |  |  |  |  |
| Standard of care | 9,245 | 6,275 | 5,090 | 64,280 | 11,750 | 41,090 |
| DH-focused (% increase from the SOC) | 27,217 (+194%) | 18,474 (+194%) | 14,985 (+194%) | 189,240 (+194%) | 34,592 (+194%) | 120,969 (+194%) |
| PHC-focused (% increase from the SOC) | 11,467 (+24%) | 7,783 (+24%) | 6,313 (+24%) | 79,728 (+24%) | 14,574 (+24%) | 50,965 (+24%) |

| *Budget requirement related to the decentralisation interventions (incremental costs)* |  |  |  |  |  |  |  |  |  |  |  |  |  |
| --- | --- | --- | --- | --- | --- | --- | --- | --- | --- | --- | --- | --- | --- |
| Interve<br>ntion | Period | Costs<br>(millions<br>US\$) | %<br>total<br>costs | Costs<br>(millions<br>US\$) | %<br>total<br>costs | Costs<br>(millions<br>US\$) | %<br>total<br>costs | Costs<br>(millions<br>US\$) | %<br>total<br>costs | Costs<br>(millions<br>US\$) | %<br>total<br>costs | Costs<br>(millions<br>US\$) | %<br>total<br>costs |
| DH-focused | 2022 | 2.8 (2.6 to 3.1) | 22% | 6.1 (5.9 to 6.3) | 28% | 5.5 (5.3 to 5.8) | 28% | 6.1 (4.9 to 7.9) | 12% | 2.4 (2.2 to 2.8) | 19% | 8.7 (7.8 to 9.9) | 18% |
|  | 2023 | 3.2 (2.9 to 3.7) | 25% | 6.3 (6 to 6.8) | 29% | 5.8 (5.5 to 6.2) | 30% | 8.5 (6.5 to 11.8) | 17% | 2.8 (2.5 to 3.5) | 22% | 10.5 (8.9 to 13.0) | 21% |
|  | 2024 | 3.5 (3.1 to 4.2) | 27% | 6.6 (6.2 to 7.3) | 30% | 6.0 (5.6 to 6.6) | 31% | 10.8 (8.0 to 15.7) | 21% | 3.2 (2.7 to 4.1) | 25% | 12.2 (10.0 to 16.1) | 25% |
|  | 2025 | 1.5 (1.0 to 2.4) | 12% | 1.2 (0.8 to 2.0) | 6% | 1.0 (0.7 to 1.7) | 5% | 11.3 (7.8 to 17.6) | 22% | 2.0 (1.4 to 3.1) | 16% | 7.9 (5.2 to 13.1) | 16% |
|  | 2026 | 1.8 (1.2 to 3.0) | 14% | 1.5 (1.0 to 2.5) | 7% | 1.3 (0.8 to 2.2) | 7% | 13.7 (9.4 to 21.5) | 27% | 2.4 (1.6 to 3.8) | 19% | 9.6 (6.3 to 16.2) | 20% |
|  | Total 2022-2026 | 12.8 (10.8 to 16.4) | 100% | 21.7 (19.9 to 24.9) | 100% | 19.6 (18.0 to 22.5) | 100 % | 50.4 (36.5 to 74.4) | 100% | 12.9 (10.4 to 17.2) | 100% | 49.0 (38.2 to 68.5) | 100 % |
| PHC-focused | 2022 | 10.0 (9.6 to 10.4) | 32% | 21.4 (20.8 to 22.0) | 33% | 29.9 (29.1 to 30.7) | 33% | 16.1 (14.7 to 17.7) | 27% | 4.0 (3.8 to 4.3) | 29% | 42.4 (40.7 to 44.1) | 32% |
|  | 2023 | 10.0 (9.6 to 10.5) | 32% | 21.5 (20.8 to 22.1) | 33% | 30.0 (29.2 to 30.8) | 33% | 16.5 (15.0 to 18.3) | 28% | 4.1 (3.8 to 4.4) | 29% | 42.7 (40.9 to 44.6) | 32% |
|  | 2024 | 10.1 (9.7 to 10.6) | 32% | 21.5 (20.9 to 22.2) | 33% | 30.0 (29.2 to 30.9) | 33% | 16.9 (15.2 to 19.0) | 28% | 4.2 (3.9 to 4.5) | 30% | 43.0 (41.1 to 45.1) | 32% |
|  | 2025 | 0.6 (0.4 to 0.9) | 2% | 0.5 (0.3 to 0.7) | 1% | 0.4 (0.3 to 0.6) | 0% | 4.7 (3.2 to 6.7) | 8% | 0.8 (0.5 to 1.1) | 6% | 3.1 (2.1 to 4.4) | 2% |
|  | 2026 | 0.7 (0.4 to 1.0) | 2% | 0.5 (0.4 to 0.8) | 1% | 0.4 (0.3 to 0.6) | 0% | 5.1 (3.5 to 7.3) | 9% | 0.8 (0.6 to 1.2) | 6% | 3.4 (2.3 to 4.9) | 3% |
|  | Total 2022-2026 | 31.4 (29.7 to 33.2) | 100% | 65.4 (63.2 to 67.7) | 100% | 90.7 (88.0 to 93.6) | 100% | 59.3 (51.6 to 69.0) | 100% | 13.9 (12.6 to 15.6) | 100% | 134.6 (127.1 to 143.0) | 100% |
| **Intervention** | **Average annual costs** | **Costs (millions US\$)** | **% of NTP budget** | **Costs (millions US\$)** | **% of NTP budget** | **Costs (millions US\$)** | **% of NTP budget** | **Costs (millions US\$)** | **% of NTP budget** | **Costs (millions US\$)** | **% of NTP budget** | **Costs (millions US\$)** | **% of NTP budget** |
| DH-focused | Program implementation costs (average of 2022-2026) | 2.6 (2.2 to 3.3) | 8% | 4.3 (4.0 to 5.0) | 54% | 3.9 (3.6 to 4.5) | 9% | 10.1 (7.3 to 14.9) | 34% | 2.6 (2.1 to 3.4) | 26% | 9.8 (7.6 to 13.7) | 22% |
|  | Routine costs (2026) | 1.8 (1.2 to 3.0) | 5% | 1.5 (1.0 to 2.5) | 19% | 1.3 (0.8 to 2.2) | 3% | 13.7 (9.4 to 21.5) | 46% | 2.4 (1.6 to 3.8) | 24% | 9.6 (6.3 to 16.2) | 21% |
| PHC-focused | Program implementation costs (average of 2022-2026) | 6.3 (5.9 to 6.6) | 18% | 13.1 (12.6 to 13.5) | 164% | 18.1 (17.6 to 18.7) | 39% | 11.9 (10.3 to 13.8) | 40% | 2.8 (2.5 to 3.1) | 28% | 26.9 (25.4 to 28.6) | 60% |
|  | Routine costs (2026) | 0.7 (0.4 to 1.0) | 2% | 0.5 (0.4 to 0.8) | 6% | 0.4 (0.3 to 0.6) | 1% | 5.1 (3.5 to 7.3) | 17% | 0.8 (0.6 to 1.2) | 8% | 3.4 (2.3 to 4.9) | 8% |

OPD: patients presenting at outpatient department, DH: district hospital, PHC: primary health centre, SOC: standard of care, NTP: National Tuberculosis Program

The average annual cost of a five-year scale-up plan of the DH-focused intervention ranged between $3M (Cambodia) and $10M (Mozambique), corresponding to between 8% (Cambodia) and 54% (Cameroon) of countries’ national tuberculosis programme (NTP) budgets in 2021, and, for the PHC-focused intervention, between $3M (Sierra Leone) and $27M (Uganda), or 18% (Cambodia) to 164% (Cameroon) of NTP budget. Once the health facilities are equipped, the average annual routine costs (corresponding to year 2026) ranged between $1·3M (3% NTP budget) in Côte d’Ivoire and $13·7M (46%) in Mozambique for DH-focused, and between $0·4M (1%) in Côte d’Ivoire and $5·1M (17%) in Mozambique for the PHC-focused strategy.

We assumed facilities saw a sustained increase in diagnoses following implementation. We projected an increase in the cumulative number treated for tuberculosis over the five-year implementation period of 194% for the DH-focused, and 24% for the PHC-focused strategy, compared to SOC. Sensitivity analyses suggested that important cost reductions could be achieved through purchase price reduction of the cartridge kits and mucus aspirators, and economies of scale on delivery chain costs (**Appendix**).

## Discussion

Benchmarked against estimates of country cost-effectiveness thresholds,^18^ this study found that the decentralisation of paediatric tuberculosis services to DH level could be cost-effective compared to the SOC from a health systems perspective in Cambodia and Côte d’Ivoire, whereas decentralisation to PHC level was unlikely to be cost-effective in any country. Ultimately, choice of cost-effectiveness thresholds is a judgement for decision-makers, and other considerations, notably the budget impact, also need to be considered. Sensitivity analysis suggests that decentralisation targeted to geographical areas with very high tuberculosis prevalence would be highly likely to be cost-effective in all countries. Implementation would require substantial financial investment in the early phase, particularly for the PHC-focused intervention.

The PHC-focused strategy was, as expected, more costly than the DH-focused strategy due to spending on diagnostic equipment for a larger number of facilities, but unexpectedly was also less effective than the DH-focused strategy. The reasons for this lower effectiveness (fewer tuberculosis diagnoses) are not clear, but may include differences in staff cadres composition and greater experience in diagnosing paediatric tuberculosis at DH level due to higher patient volumes.^12^ The higher tuberculosis diagnosis rate in children seeking care at DH level may indicate care-givers prefer to take more severely ill children to this level first or because children are referred from PHC with more advanced disease. We did not model differences in tuberculosis severity by location which may have favoured the DH-focused strategy. Finally, all children enrolled in DH had a CXR performed while in PHCs, only children with persisting symptoms after 7 days were referred for CXR at DH; this is likely to have contributed to higher tuberculosis detection in the DH-focused strategy. We did not include patient costs in our analysis; the PHC-focused strategy is likely to have reduced costs to patients due to fewer referrals.

We used a single typical SOC as a comparator across the six countries. It was not practical or transparent to represent every variation that exists within and across these countries. In our iterative conceptual modelling, we agreed on a structure clinicians from all countries felt adequately represented patient pathways in their country. However, paediatric tuberculosis care was already partially decentralised to PHC level in Uganda and Mozambique, meaning our modelled SOC is more of an approximation in these settings. Some parameters were derived by calibrating to cascade data, and our estimate of the true tuberculosis prevalence in children seeking care across all countries was uncertain. In reality tuberculosis prevalence will vary between countries, and higher prevalence values substantially improved cost-effectiveness (**Appendix**).

We were also limited by the nature of the TB-Speed Decentralization study and data. The pre-/post-intervention primary outcome means results are vulnerable to confounding by factors that changed over time. In particular, the study period overlapped with the COVID-19 pandemic. Restrictions introduced by countries affected transportation (less available and more costly) and these barriers may have increased patient losses during referral to hospitals. Fear of COVID-19 also reduced facility attendance. Facility-level overheads costs were recently reported to be a significant contributor to total costs in adult tuberculosis costing estimates from the Value-TB project (the largest tuberculosis costing effort to date).^21^ These were excluded in our study because it was anticipated that the adoption of a new diagnostic strategy without change to the number of health facilities or staff would not significantly affect overheads, but this assumption should be carefully considered alongside roll-out plans. Data collection, as well as our analysis, was focused on children, which ignores the additional benefits from decentralisation of tuberculosis diagnostic capacity in improving the detection of tuberculosis in adults, as well as detection of rifampicin-resistant tuberculosis and other pathogens using the GeneXpert platform.

A major strength of this study is the collection and analysis of primary data to inform costs, impacts, and care cascades in six high tuberculosis incidence countries in different regions of Africa and in South-East Asia. This diversity in economic and health system context suggests the generalisability of these results across comparable settings. Underlying modelling assumptions were based on a number of previously published studies, and applied within a framework that captured considerable complexity in patient pathways, including reassessments and referrals between levels.

To our knowledge, this is the first multi-country study to assess the cost-effectiveness or budget impact of decentralising childhood tuberculosis services, and will add to the evidence base for the interim WHO recommendation on decentralised models of care. Systematic reviews of the cost and cost-effectiveness of tuberculosis screening have found ICERs of between US$281 and US$698 per DALY averted in the general population,^22^ but for screening in high risk groups ICERs as low as US$51 per DALY have been reported.^23^ An analysis of the cost-effectiveness of Xpert on stool for paediatric tuberculosis in Ethiopia and Indonesia found ICERs of US$132 and US$94 per DALY, respectively.^24^ However, the majority of these analyses were not based on empirical data from implemented interventions, and restricted to a small component of tuberculosis diagnosis given that the majority of children are clinically diagnosed.

A number of studies have explored the effects of decentralisation of general tuberculosis services and costs. A 2017 study assessed the cost per tuberculosis diagnosis of implementing Xpert testing regardless of age at PHCs in Uganda.^25^ Average costs per new tuberculosis diagnosis using Xpert averaged US$119, but were as high as US$885 in the lowest-volume centre. The authors did not attempt to estimate cost-effectiveness in terms of cost per DALY averted. Thompson and colleagues estimated the cost-effectiveness of an adult tuberculosis diagnostic strategy decentralised in community health centres in Uganda.^26^ The authors recommended decentralised testing services with ICERs ranging US$687 per additional treatment initiation in 14 days and US$1,332 per additional tuberculosis diagnosis. Cost-effectiveness notably increased with high testing volumes and lower equipment costs.

TB-Speed Decentralization and other studies have demonstrated the potential for decentralised diagnostic approaches to find more children with tuberculosis.^12,27^ Our analysis shows that this is possible in a way that could be considered cost-effective across a range of settings, depending on strategy and tuberculosis prevalence, but that large-scale implementation would still incur substantial costs relative to existing NTP budgets. Countries should consider scaling up locally-adapted interventions to improve tuberculosis diagnosis while monitoring their performance, potentially prioritising areas with the highest tuberculosis prevalence, and including such plans when identifying domestic or donor sources of funding.

## Contributors

MH, NM, PD, OM, MB, EW conceived the health economics analysis plan. PD developed the cost-effectiveness model and MDE developed the budget impact model. MDE conducted the cost, the cost-effectiveness and the budget impact analyses. MH coordinated international economic data collection and design of the patient pathways. MDE, PD, MH, NM interpreted economic results. TB, AB, HS provided scientific expertise and guidance.

OM, MB, EW conceived and designed the TB-Speed Decentralization study. MN coordinated and contributed to the implementation of the study protocol. OM, MB, and EW led the study at international level. LB led the study in Cambodia, J-VT led the study in Cameroon, RM led the study in Côte d’Ivoire, CK led the study in Mozambique, JM led the study in Sierra Leone, EW and JM-A led the study in Uganda, MN coordinated study implementation at international level. MHTNN did the statistical analysis of the decentralisation study. OM, MN, MH, MB, EW and MDE contributed to the interpretation of the results.

MDE wrote the first draft and all authors reviewed, edited and approved the final version of the manuscript. MDE, PD, OM, MB, EW were responsible for the decision to submit the manuscript.

TB-Speed Health Economics Study Group.

TB-Speed Health Economics Study Group
| Name | Affiliation |
| --- | --- |
| Arlt-Hilares, Doris | MSF-Logistique, Merignac, France |
| Balestre, Eric | University of Bordeaux, National Institute for Health and Medical Research UMR 1219, Research Institute for Sustainable Development EMR 271, Bordeaux, France |
| Banga, Marie-France | Programme PAC-CI, CHU de Treichville, Abidjan, Côte d'Ivoire |
| Bénard, Antoine | CHU Bordeaux, Service d'information Médicale, USMR & CIC-EC 14-01, Bordeaux, France |
| Bernard, Tanguy | Bordeaux School of Economics, Bordeaux, France |
| Bonnet, Maryline | TransVIHMI, University of Montpellier, IRD /INSERM, Montpellier, France |
| Borand, Laurence | Center for Tuberculosis Research, Division of Infectious Diseases, Johns Hopkins University School of Medicine, Baltimore, MD USA<br>Epidemiology and Public Health Unit, Clinical Research Group, Institut Pasteur du Cambodge, Phnom Penh, Cambodia |
| Breton, Guillaume | SOLTHIS, Paris, France |
| Bunnet, Dim | Epidemiology and Public Health Unit, Clinical Research Group, Institut Pasteur du Cambodge, Phnom Penh, Cambodia |
| Chateau, Paul-Damien | MSF-Logistique, Merignac, France |
| Cumbe, Saniata | Instituto Nacional de Saúde, Marracuene, Mozambique |
| d'Elbée, Marc | University of Bordeaux, National Institute for Health and Medical Research UMR 1219, Research Institute for Sustainable Development EMR 271, Bordeaux, France<br>Ceped UMR 196, Université Paris Cité, Research Institute for Sustainable Development (IRD), Inserm, Paris, France |
| de Lauzanne, Agathe | Epidemiology and Public Health Unit, Clinical Research Group, Institut Pasteur du Cambodge, Phnom Penh, Cambodia |
| Dodd, Peter J. | School of Health & Related Research, University of Sheffield, Sheffield, United Kingdom |
| Harker, Martin | TB Modelling Group, TB Centre, and Global Centre for Health Economics, London School of Hygiene and Tropical Medicine, London, United Kingdom |
| Huyen Ton Nu Nguyet, Minh | University of Bordeaux, National Institute for Health and Medical Research UMR 1219, Research Institute for Sustainable Development EMR 271, Bordeaux, France |
| Kaing, Sanary | Epidemiology and Public Health Unit, Clinical Research Group, Institut Pasteur du Cambodge, Phnom Penh, Cambodia |
| Khosa, Celso | Instituto Nacional de Saúde, Marracuene, Mozambique |
| Komena, Eric | Programme PAC-CI, CHU de Treichville, Abidjan, Côte d'Ivoire |
| Koroma, Monica | SOLTHIS, Sierra Leone |
| Kwedi Nolna, Sylvie | University of Montpellier, TransVIHMI, IRD /INSERM, Montpellier, France |
| Mafirakureva, Nyashadzaishe | School of Health & Related Research, University of Sheffield, Sheffield, United Kingdom |
| Mao, Tan Eang | Cambodia National Tuberculosis Programme |
| Marcy, Olivier | University of Bordeaux, National Institute for Health and Medical Research UMR 1219, Research Institute for Sustainable Development EMR 271, Bordeaux, France |
| Mbang Masson, Douglas | Mother and Child Center, Chantal Biya Foundation, Yaoundé, Cameroon |
| Moh, Raoul | Teaching Unit of Dermatology and Infectiology, UFR of Medical Sciences, Félix-Houphouët Boigny University, Abidjan, Côte d'Ivoire<br>Programme PAC-CI, CHU de Treichville, Abidjan, Côte d'Ivoire |
| Mugisha, Jacob | SOLTHIS, Sierra Leone |
| Mustapha, Ayeshatu | Ola During Children's Hospital, Freetown, Sierra Leone |
| Mwanga-Amumpere, Juliet | Epicentre Mbarara Research Centre, Mbarara, Uganda |
| Nafuka, Mastula | MU-JHU Care Ltd, MUJHU Research Collaboration, Kampala, Uganda |
| Natukunda, Naome | Epicentre Mbarara Research Centre, Mbarara, Uganda |
| Orne-Gliemann, Joanna | University of Bordeaux, National Institute for Health and Medical Research UMR 1219, Research Institute for Sustainable Development EMR 271, Bordeaux, France |
| Ouattara, Eric | University of Bordeaux, National Institute for Health and Medical Research UMR 1219, |
|  | Research Institute for Sustainable Development EMR 271, Bordeaux, France |
| Poublan, Julien | University of Bordeaux, National Institute for Health and Medical Research UMR 1219, Research Institute for Sustainable Development EMR 271, Bordeaux, France |
| Sohn, Hojoon | Seoul National University College of Medicine, Seoul, South Korea |
| Taguebue, Jean-Voisin | Mother and Child Center, Chantal Biya Foundation, Yaoundé, Cameroon |
| Tulinawe, Immaculate | Epicentre Mbarara Research Centre, Mbarara, Uganda |
| Voss de Lima, Yara | Instituto Nacional de Saúde, Marracuene, Mozambique |
| Wittwer, Jérôme | University of Bordeaux, National Institute for Health and Medical Research UMR 1219, Bordeaux, France |
| Wobudeya, Eric | MU-JHU Care Ltd, MUJHU Research Collaboration, Kampala, Uganda |

## Data sharing

Aggregated data for all analyses will be publicly available with the publication on a Github repository under a Creative Commons Attribution (CC BY) licence (URL: https://github.com/petedodd/TBSdecent).

## Data Availability

Aggregated data for all analyses will be publicly available with the publication on a Github repository under a Creative Commons Attribution (CC BY) licence

https://github.com/petedodd/TBSdecent

## Acknowledgements

We thank all the children and their families who participated in the study and the healthcare workers of the participating hospitals and laboratories. We thank the Ministries of Health and National Tuberculosis Programmes of participating countries for their support. We thank members of the TB-Speed Scientific Advisory Board who gave technical advice on the design of the study and approved the protocol: Anneke Hesseling (Stellenbosch University, Cape Town, South Africa), Luis Cuevas (Liverpool School of Tropical Medicine, Liverpool, UK), Malgorzata Grzemska and Sabine Verkuijl (WHO, Geneva, Switzerland), Philippa Musoke (Makerere University, Kampala, Uganda), and Mark Nicol (University of Western Australia, Perth, WA, Australia). This work received support from the University of Bordeaux, Bordeaux, France, and the University Félix Houphouët Boigny, Abidjan, Côte d’Ivoire, within the IPORA (Interdisciplinary Policy-Oriented Research for Africa) network.

## References

1 Perin J, Mulick A, Yeung D, et al. Global, regional, and national causes of under-5 mortality in 2000-19: an updated systematic analysis with implications for the Sustainable Development Goals. Lancet Child Adolesc Health 2021; published online Nov 17. DOI:10.1016/S2352-4642(21)00311-4.

2 World Health Organization. Global tuberculosis report 2022. https://apps.who.int/iris/rest/bitstreams/1474924/retrieve.

3 Dodd PJ, Yuen CM, Sismanidis C, Seddon JA, Jenkins HE. The global burden of tuberculosis mortality in children: a mathematical modelling study. Lancet Glob Health 2017; 5: e898–906.

4 Wobudeya E, Bonnet M, Walters EG, et al. Diagnostic Advances in Childhood Tuberculosis-Improving Specimen Collection and Yield of Microbiological Diagnosis for Intrathoracic Tuberculosis. Pathogens 2022; 11. DOI:10.3390/pathogens11040389.

5 Zar HJ, Workman L, Isaacs W, et al. Rapid molecular diagnosis of pulmonary tuberculosis in children using nasopharyngeal specimens. Clin Infect Dis 2012; 55: 1088–95.

6 Gous N, Scott LE, Khan S, Reubenson G, Coovadia A, Stevens W. Diagnosing childhood pulmonary tuberculosis using a single sputum specimen on Xpert MTB/RIF at point of care. S Afr Med J 2015; 105: 1044–8.

7 Oliwa JN, Gathara D, Ogero M, et al. Diagnostic practices and estimated burden of tuberculosis among children admitted to 13 government hospitals in Kenya: An analysis of two years’ routine clinical data. PLoS One 2019; 14: e0221145.

8 Reid MJA, Goosby E. Patient-Centered Tuberculosis Programs Are Necessary to End the Epidemic. J Infect Dis 2017; 216: S673–4.

9 WHO consolidated guidelines on tuberculosis. Module 5: management of tuberculosis in children and adolescents. World Health Organization, 2022.

10 Marcy O, Ung V, Goyet S, et al. Performance of Xpert MTB/RIF and Alternative Specimen Collection Methods for the Diagnosis of Tuberculosis in HIV-Infected Children. Clinical Infectious Diseases. 2016; 62: 1161–8.

11 Song R, Click ES, McCarthy KD, et al. Sensitive and Feasible Specimen Collection and Testing Strategies for Diagnosing Tuberculosis in Young Children. JAMA Pediatr 2021;: e206069.

12 Wobudeya E, Nanfuka M, Huyen Ton Nu Nguyet M, et al. Impact of decentralizing childhood tuberculosis diagnosis at primary health center and district hospital level - A pre-post study in six high tuberculosis burden countries.

13 Cunnama L, Garcia Baena I, Gomez G, et al. Costing guidelines for tuberculosis interventions. World Health Organization, 2020.

14 Global Drug Facility. August 2022 Medicines Catalog. https://www.stoptb.org/sites/default/files/gdfmedicinescatalog_1.pdf.

15 World Health Organization. WHO-CHOICE estimates of cost for inpatient and outpatient health service delivery. https://www.who.int/publications/m/item/who-choice-estimates-of-cost-for-inpatient-and-outpatient-health-service-delivery.

16 Wilkinson T, Sculpher MJ, Claxton K, et al. The International Decision Support Initiative Reference Case for Economic Evaluation: An Aid to Thought. Value Health 2016; 19: 921–8.

17 Tan-Torres Edejer T, Baltussen R, Adam T, et al., editors. Making choices in health: WHO guide to cost-effectiveness. World Health Organization.

18 Ochalek J, Lomas J, Claxton K. Estimating health opportunity costs in low-income and middle-income countries: a novel approach and evidence from cross-country data. BMJ Glob Health 2018; 3: e000964.

19 Woods B, Revill P, Sculpher M, Claxton K. Country-Level Cost-Effectiveness Thresholds: Initial Estimates and the Need for Further Research. Value Health 2016; 19: 929–35.

20 Husereau D, Drummond M, Augustovski F, et al. Consolidated Health Economic Evaluation Reporting Standards 2022 (CHEERS 2022) Statement: Updated Reporting Guidance for Health Economic Evaluations. Value Health 2022; 25: 3–9.

21 Sweeney S, Laurence YV, Cunnama L, et al. Cost of TB services: approach and summary findings of a multi-country study (Value TB). Int J Tuberc Lung Dis 2022; 26: 1006–15.

22 Empringham B, Alsdurf H, Miller C, Zwerling A. How much does TB screening cost? A systematic review of economic evaluations. Int J Tuberc Lung Dis 2022; 26: 38–43.

23 Alsdurf H, Empringham B, Miller C, Zwerling A. Tuberculosis screening costs and cost-effectiveness in high-risk groups: a systematic review. BMC Infect Dis 2021; 21: 935.

24 Mafirakureva N, Klinkenberg E, Spruijt I, et al. Xpert Ultra stool testing to diagnose tuberculosis in children in Ethiopia and Indonesia: a model-based cost-effectiveness analysis. BMJ Open 2022; 12: e058388.

25 Hsiang E, Little KM, Haguma P, et al. Higher cost of implementing Xpert(®) MTB/RIF in Ugandan peripheral settings: implications for cost-effectiveness. Int J Tuberc Lung Dis 2016; 20: 1212–8.

26 Thompson RR, Nalugwa T, Oyuku D, et al. Multicomponent strategy with decentralised molecular testing for tuberculosis in Uganda: a cost and cost-effectiveness analysis. Lancet Glob Health 2023; 11: e278–86.

27 Zawedde-Muyanja S, Nakanwagi A, Dongo JP, et al. Decentralisation of child tuberculosis services increases case finding and uptake of preventive therapy in Uganda. Int J Tuberc Lung Dis 2018; 22: 1314–21.

