## Supplementary materials for "Cost-effectiveness and budget impact of decentralising childhood tuberculosis diagnosis: a mathematical modelling study in six high tuberculosis incidence countries"


**Abbreviations** **2**

**Study intervention and patient care pathways** **3**

**Costing analysis** **9**

- Scope of costing 9
- Supplementary results - Costing analysis 9
- Supplementary results - Time and motion study 14

**Cost-effectiveness modelling** **16**

- Overview of model design and implementation 16
- Cost-effectiveness analysis 17
- Modelling of diagnostic accuracy 18
- Model parameters 18
- Calibration to cascade data 21
- CHEERS 2022 - Checklist 36
- Country level probabilities of cost-effectiveness and cost-effectiveness thresholds 39
- Supplementary results - Screening to diagnosis cascades during the intervention period by decentralisation approach and age group 41
- Supplementary results - Other modelled health outcomes and cost per health outcome 42
- Supplementary results - Cost per child treated by cost input and by care stage 44
- Supplementary results - Sensitivity analysis 46

**Budget impact analysis** **48**

- Methodological approach 48
- Supplementary results - Scale-up costs by intervention, year, cost input, and country 51
- Supplementary results - Sensitivity analysis 62

**Ethics committees and review boards for the study** **68**

**References** **69**

### Abbreviations

CFR: Case-Fatality Rate
CXR: Chest X-Ray
DALY: Disability-Adjusted Life Year
DH: District Hospital
ERC: Ethics Review Committee
GA: Gastric Aspirate
GDP: Growth Domestic Product
GLMM: Generalised Linear Mixed Model
HCW: Health Care Worker
ICER: Incremental Cost-Effectiveness Ratio
IDH: DH-focused Intervention
IPH: PHC-focused Intervention
IQR: InterQuartile Range
NPA:  NasoPharyngeal Aspirate
OPD: OutPatient Department
OR: Odds Ratio
PHC: Primary Health Centre
PSA: Probabilistic Sensitivity Analysis
RifR-TB: Rifampicin-Resistant Tuberculosis
RifS-TB: Rifampicin-Sensitive Tuberculosis
SD: Standard Deviation
SOC: Standard Of Care
TB: Tuberculosis
WHO: World Health Organisation

### Study intervention and patient care pathways

The intervention was implemented at two levels: at patient level with the childhood tuberculosis diagnosis package, and at health system level with the decentralisation approaches. The patient level intervention consisted of:

1. systematic tuberculosis screening (four simple specific screening questions by a HCW at healthcare OPD entry point or during consultation aiming to identify children with symptoms representing presumptive tuberculosis),
   and then, for all children identified with presumptive tuberculosis:
2. a clinical evaluation (detailed history taking, relevant physical examination, and severity of illness assessment),
3. a microbiological evaluation based on NPA and stool or sputum testing using Xpert Ultra (but not a precondition to diagnosis), and
4. a radiological evaluation with an optimised CXR reading approach (using digital radiography, improvement of reading skills, simplified reading tool, and quality control of CXR reading).

The health system level intervention consisted of two decentralisation approaches compared to a status quo (**Figure 1**). In the DH-focused approach, the comprehensive childhood tuberculosis diagnosis package was implemented at the DH while only systematic tuberculosis screening was implemented at the PHC (**Figure 2**). All children identified as having presumptive tuberculosis by screening at PHC were referred to the DH for tuberculosis clinical evaluation, Xpert Ultra testing on NPA and stool or expectorated sputum, and CXR. Referral transportation to the DH was not covered by the study. Tracking of referrals was set-up by the study. In the PHC-focused approach, the comprehensive childhood tuberculosis diagnosis package was implemented at both the DH and the PHC, with the exception of CXR at PHC (**Figure 3**). Children not diagnosed with tuberculosis at enrolment had a tuberculosis clinical assessment after 7 days. They were referred to the DH for CXR if still symptomatic at 7 days.

In participating countries (Mozambique and Uganda) that had already partially decentralised tuberculosis diagnosis and treatment initiation in children at PHC level using clinical evaluation and sputum collection, these PHCs continued to provide these services instead of the pure DH-focused approach (**Figure 4**). Only children whom clinicians decided not to treat for tuberculosis according to national guidelines were referred to the DH for further investigations and in those cases the decision to initiate treatment was taken at the DH. Although these two countries had some partially decentralised tuberculosis care services, these were considered to be minor compared to the overall impact of the decentralisation interventions assessed across countries, and so effectiveness results from these countries are included as part of the DH-focused approach in the results for this paper. The impact assessment on tuberculosis case detection of the decentralisation intervention is presented by Wobudeya et al.[1]

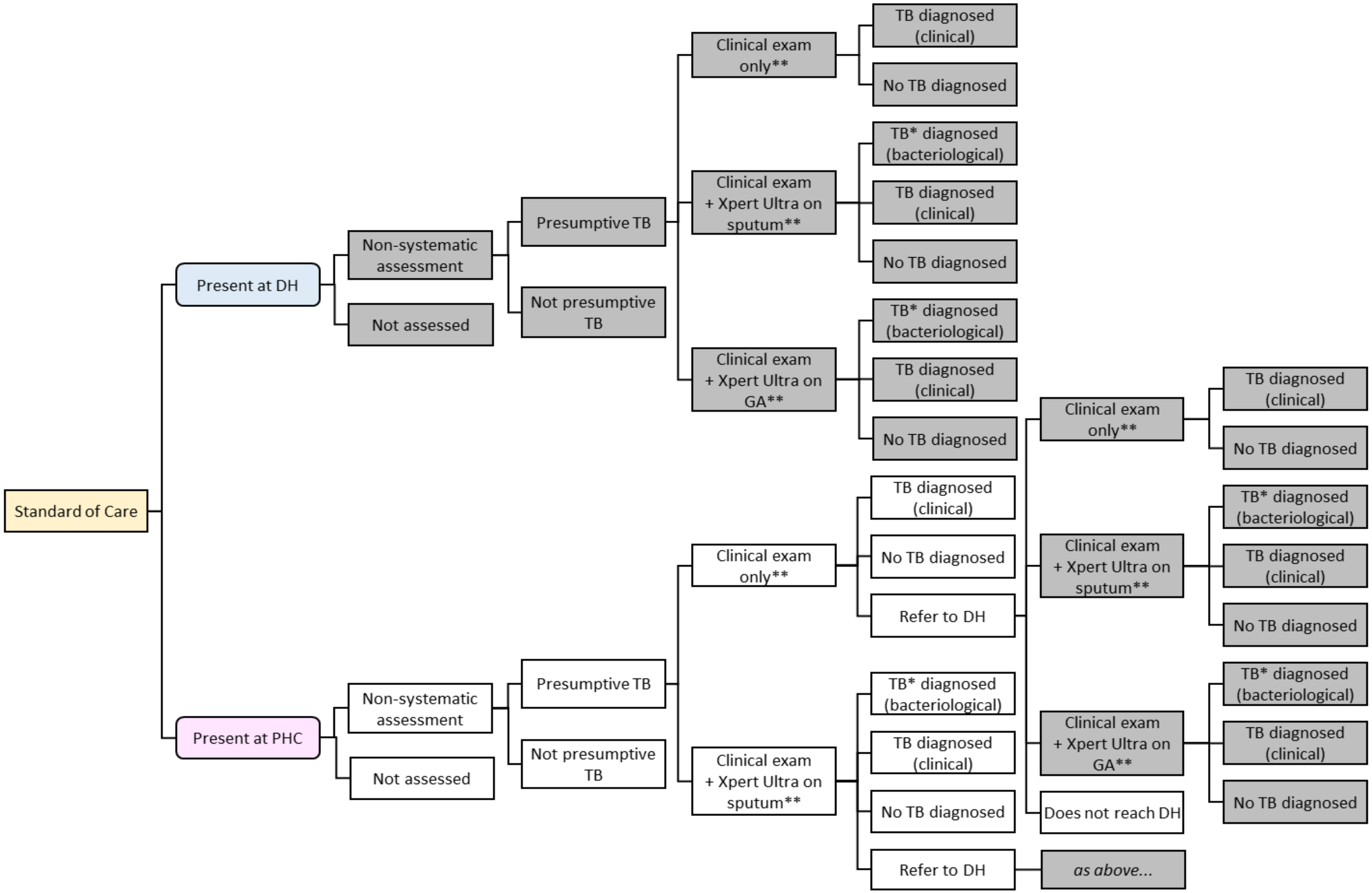


**Figure 1**. Standard of care pathway for the diagnosis and treatment of tuberculosis in children

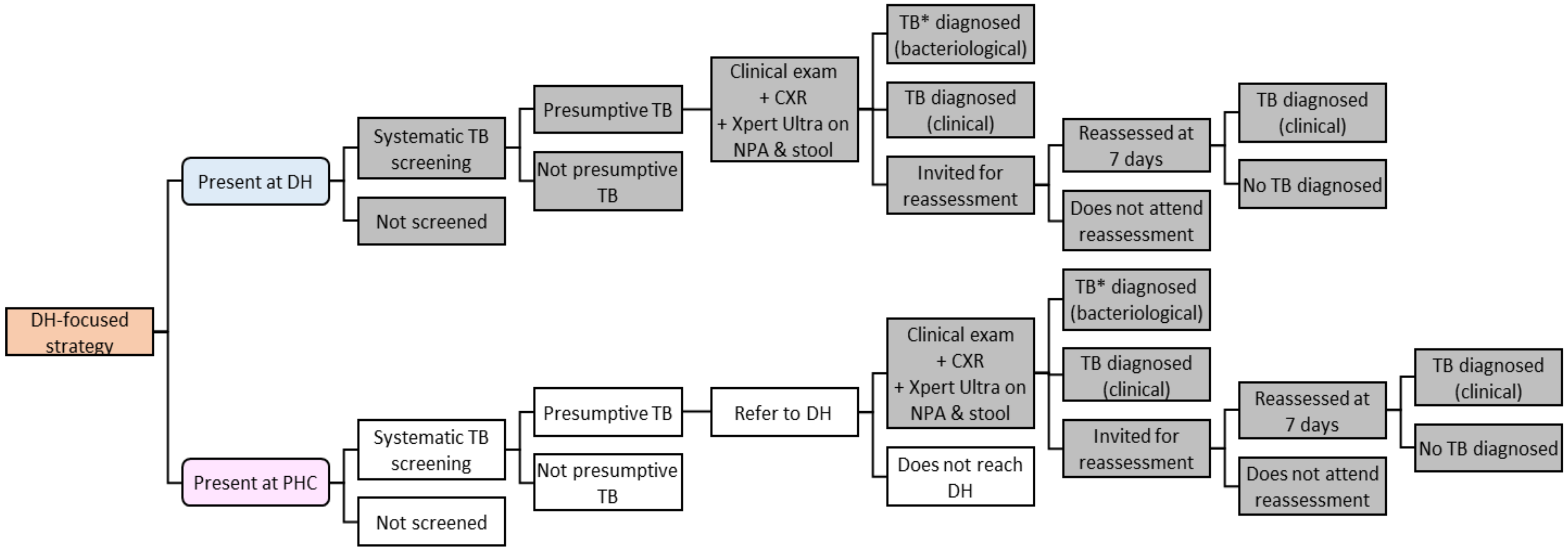


**Figure 2**. DH-focused pathway for the diagnosis and treatment of tuberculosis in children

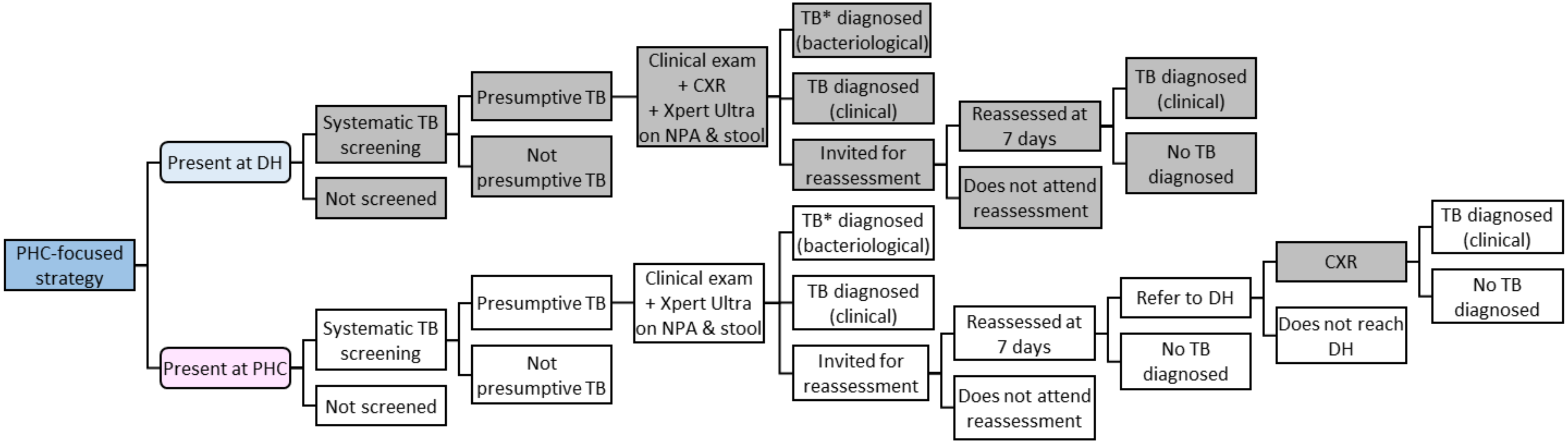


**Figure 3**. PHC-focused pathway for the diagnosis and treatment of tuberculosis in children

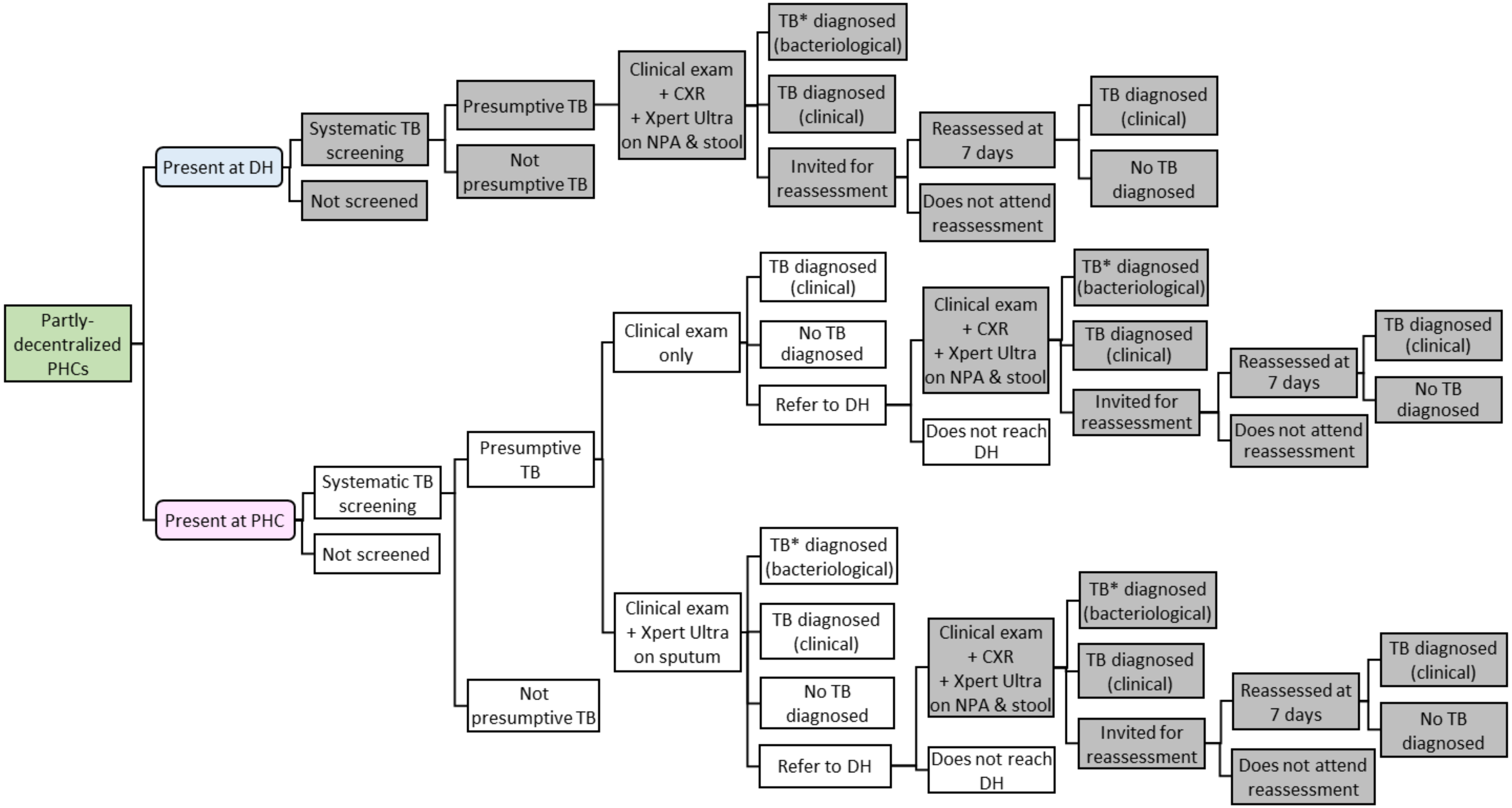


**Figure 4**. Partially decentralised strategy for the diagnosis and treatment of tuberculosis in children

**Notes**

- Boxes shaded in **grey** show activities undertaken at DH. Unshaded boxes show activities undertaken at PHC
- * Tuberculosis diagnosis includes the possibility of RifR-TB (see below)
- **Smear microscopy and/or (in DH only) CXR also occurs for a proportion of patients. This is incorporated in the costs, but does not lead to differences in diagnosis rates, treatment rates or outcomes
- "Clinical" diagnosis includes diagnosis partially or wholly based on the findings of chest X-ray (CXR)
- Activity following presentation at PHC is different in all 4 pathways. Activity following presentation at DH is the same in all non-SOC pathways

**Pathway endings**

Where Xpert MTB/RIF Ultra testing is undertaken, tuberculosis diagnosis may be of rifampicin-sensitive tuberculosis (RifS-TB) or rifampicin-resistant tuberculosis (RifR-TB). Each is treated using the appropriate treatment programme in parallel strategies.

Where tuberculosis is diagnosed based on clinical examination (alone, or with CXR results) without positive Xpert results, tuberculosis is assumed to be RifS-TB and so RifS-TB treatment is given (which may be inappropriate due to RifR-TB being unrecognised).

Each branch ending in 'No TB diagnosed' in the main figure is followed by the ending presented in **Figure 5**.

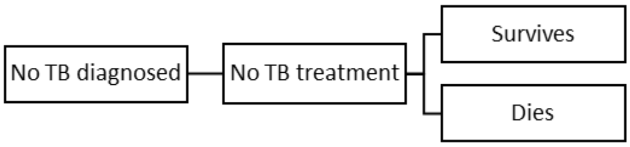


**Figure 5**. Branch ending in 'No TB diagnosed'

Each branch ending in 'TB diagnosed (clinical)' in the main figure is followed by the ending presented in **Figure 6**.

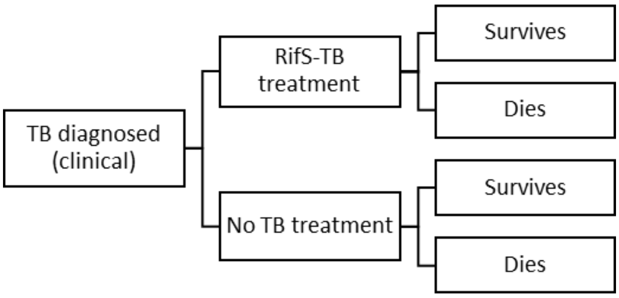


**Figure 6**. Branch ending in 'TB diagnosed (clinical)'

Each branch ending in 'TB* diagnosed (bacteriological)' in the main figure is followed by the ending presented in **Figure 7**.

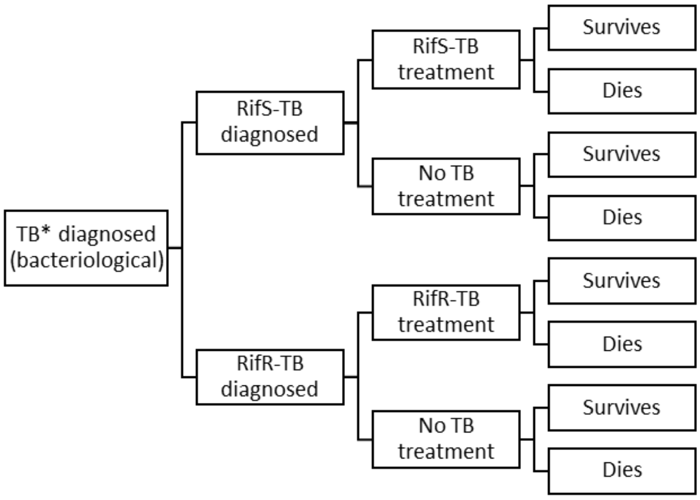


**Figure 7**. Branch ending in 'TB* diagnosed (bacteriological)'

### Costing analysis

#### Scope of costing

This study followed a health service perspective; resource use falling upon patients, such as transportation and loss of earnings were not measured. This study was not expected to increase the number of children presenting to health facilities, and it was not considered reasonable that health services would change the number of facilities in response to adopting a different diagnosis strategy. Consequently, the economic analyses assumed that only current facilities would be used. Accordingly, this analysis did not include costs of running a health centre or hospital, buildings, furniture, administration and management, utilities, cleaning and security. Similarly, there are shared costs of operating the infrastructure of a national health system and national tuberculosis programme. We did not anticipate that these would be materially affected by the adoption of a new diagnostic strategy, and so these costs were also excluded. As a result, the resource use and costs collected for the cost-effectiveness analysis did not intend to reflect the full total costs to the health system of diagnosing and treating each patient. However, the difference in costs between alternative strategies (the incremental cost) was intended to reflect the true incremental cost. Costs are presented with low and high estimates accounting for heterogeneity in cost inputs (such as the higher salary of a senior nurse doing the same activity as a more junior nurse, or variation of unit costs for supplies).

#### Supplementary results - Costing analysis

Cost analysis results are presented by country, type of facility, TB-Speed activity, and cost inputs in **Table 1**.

**Table 1.** Average cost of TB-Speed activities per 100 patients, by cost inputs, type of facility, and country (in 2021 US$)

| Activity | | Cambodia | | Cameroon | | Côte d'Ivoire | | Mozambique | | Sierra Leone | | Uganda | |
|---|---|---|---|---|---|---|---|---|---|---|---|---|---|
| | Health facility | DH | PHC | DH | PHC | DH | PHC | DH | PHC | DH | PHC | DH | PHC |
| **Systematic or non-systematic tuberculosis screening** | **Total** | **20.48** | **20.48** | **27.69** | **27.69** | **36.60** | **36.60** | **15.82** | **15.82** | **15.82** | **15.82** | **30.44** | **30.44** |
| | Personnel | 20.30 | 20.30 | 27.51 | 27.51 | 36.42 | 36.42 | 15.64 | 15.64 | 15.64 | 15.64 | 30.26 | 30.26 |
| | Training | 0.18 | 0.18 | 0.18 | 0.18 | 0.18 | 0.18 | 0.18 | 0.18 | 0.18 | 0.18 | 0.18 | 0.18 |
| **Initial clinical examination or second check-up at 7 days: history, symptoms, and physical exam** | **Total** | **97.92** | **96.31** | **188.05** | **178.72** | **208.96** | **157.14** | **187.63** | **143.88** | **228.55** | **93.21** | **166.10** | **163.80** |
| | Personnel | 42.21 | 40.60 | 64.36 | 55.03 | 124.65 | 72.84 | 75.02 | 31.27 | 166.61 | 31.27 | 62.82 | 60.53 |
| | Supplies | 39.93 | 39.93 | 39.93 | 39.93 | 40.02 | 40.02 | 40.02 | 40.02 | 40.02 | 40.02 | 40.02 | 40.02 |
| | Training | 0.36 | 0.36 | 0.36 | 0.36 | 0.36 | 0.36 | 0.36 | 0.36 | 0.36 | 0.36 | 0.36 | 0.36 |
| | Supply chain | 15.42 | 15.42 | 83.40 | 83.40 | 43.93 | 43.93 | 72.23 | 72.23 | 21.57 | 21.57 | 62.90 | 62.90 |
| **Initial clinical examination: vital signs and measurements** | **Total** | **95.82** | **95.82** | **162.79** | **162.79** | **153.00** | **153.00** | **128.18** | **128.18** | **90.18** | **90.18** | **153.59** | **153.59** |
| | Personnel | 44.98 | 44.98 | 60.96 | 60.96 | 80.69 | 80.69 | 34.64 | 34.64 | 34.64 | 34.64 | 67.05 | 67.05 |
| | Supplies | 38.88 | 38.88 | 38.88 | 38.88 | 38.96 | 38.96 | 38.96 | 38.96 | 38.96 | 38.96 | 38.96 | 38.96 |
| | Training | 0.40 | 0.40 | 0.40 | 0.40 | 0.40 | 0.40 | 0.40 | 0.40 | 0.40 | 0.40 | 0.40 | 0.40 |
| | Supply chain | 11.56 | 11.56 | 62.55 | 62.55 | 32.95 | 32.95 | 54.18 | 54.18 | 16.17 | 16.17 | 47.18 | 47.18 |
| **Collecting an NPA sample** | **Total** | **1268.39** | **1845.16** | **1582.89** | **2261.97** | **1503.86** | **2344.32** | **1449.03** | **2181.00** | **1397.20** | **2316.82** | **1386.13** | **2025.24** |
| | Personnel | 40.11 | 40.11 | 40.11 | 40.11 | 65.56 | 65.56 | 42.74 | 42.74 | 42.74 | 42.74 | 40.11 | 40.11 |
| | Equipment | 816.33 | 1328.48 | 816.33 | 1318.62 | 816.33 | 1503.89 | 816.33 | 1434.72 | 816.33 | 1609.60 | 816.33 | 1366.28 |
| | Supplies | 266.08 | 266.08 | 266.08 | 266.08 | 266.08 | 266.08 | 266.08 | 266.08 | 266.08 | 266.08 | 266.08 | 266.08 |

| Activity | | Cambodia | | Cameroon | | Côte d'Ivoire | | Mozambique | | Sierra Leone | | Uganda | |
|---|---|---|---|---|---|---|---|---|---|---|---|---|---|
| | **Health facility** | **DH** | **PHC** | **DH** | **PHC** | **DH** | **PHC** | **DH** | **PHC** | **DH** | **PHC** | **DH** | **PHC** |
| | Training | 0.32 | 0.32 | 0.32 | 0.32 | 0.32 | 0.32 | 0.32 | 0.32 | 0.32 | 0.32 | 0.32 | 0.32 |
| | Supply chain | 145.55 | 210.17 | 460.05 | 636.84 | 355.58 | 508.48 | 323.56 | 437.14 | 271.73 | 398.08 | 263.29 | 352.46 |
| **Collecting a GA sample** | **Total** | **1268.39** | **1845.16** | **1582.89** | **2261.97** | **1503.86** | **2344.32** | **1449.03** | **2181.00** | **1397.20** | **2316.82** | **1386.13** | **2071.96** |
| | Personnel | 40.11 | 40.11 | 40.11 | 40.11 | 65.56 | 65.56 | 42.74 | 42.74 | 42.74 | 42.74 | 40.11 | 40.11 |
| | Equipment | 816.33 | 1328.48 | 816.33 | 1318.62 | 816.33 | 1503.89 | 816.33 | 1434.72 | 816.33 | 1609.60 | 816.33 | 1412.99 |
| | Supplies | 266.08 | 266.08 | 266.08 | 266.08 | 266.08 | 266.08 | 266.08 | 266.08 | 266.08 | 266.08 | 266.08 | 266.08 |
| | Training | 0.32 | 0.32 | 0.32 | 0.32 | 0.32 | 0.32 | 0.32 | 0.32 | 0.32 | 0.32 | 0.32 | 0.32 |
| | Supply chain | 145.55 | 210.17 | 460.05 | 636.84 | 355.58 | 508.48 | 323.56 | 437.14 | 271.73 | 398.08 | 263.29 | 352.46 |
| **Collecting and preparing a stool sample** | **Total** | **121.29** | **330.50** | **182.62** | **425.65** | **147.17** | **404.96** | **160.85** | **398.08** | **110.62** | **372.79** | **164.83** | **391.83** |
| | Personnel | 19.91 | 19.91 | 24.11 | 24.11 | 28.89 | 28.89 | 15.64 | 15.64 | 15.64 | 15.64 | 29.81 | 29.81 |
| | Equipment | 39.90 | 249.11 | 43.33 | 286.36 | 42.60 | 300.38 | 41.40 | 278.62 | 41.79 | 303.96 | 40.65 | 267.65 |
| | Supplies | 49.45 | 49.45 | 51.66 | 51.66 | 41.87 | 41.87 | 48.95 | 48.95 | 36.28 | 36.28 | 46.61 | 46.61 |
| | Training | 0.18 | 0.18 | 0.18 | 0.18 | 0.18 | 0.18 | 0.18 | 0.18 | 0.18 | 0.18 | 0.18 | 0.18 |
| | Supply chain | 11.85 | 11.85 | 63.35 | 63.35 | 33.63 | 33.63 | 54.69 | 54.69 | 16.73 | 16.73 | 47.58 | 47.58 |
| **Collecting a sputum sample** | **Total** | **83.73** | **83.86** | **122.77** | **122.88** | **96.61** | **97.00** | **107.20** | **107.48** | **76.24** | **76.79** | **117.32** | **117.57** |
| | Personnel | 20.12 | 20.12 | 22.45 | 22.45 | 25.93 | 25.93 | 15.46 | 15.46 | 22.45 | 22.45 | 32.68 | 32.68 |
| | Equipment | 20.40 | 20.52 | 20.40 | 20.51 | 20.40 | 20.79 | 20.40 | 20.68 | 20.40 | 20.95 | 20.40 | 20.65 |

| Activity | | Cambodia | | Cameroon | | Côte d'Ivoire | | Mozambique | | Sierra Leone | | Uganda | |
|---|---|---|---|---|---|---|---|---|---|---|---|---|---|
| | **Health facility** | **DH** | **PHC** | **DH** | **PHC** | **DH** | **PHC** | **DH** | **PHC** | **DH** | **PHC** | **DH** | **PHC** |
| | Supplies | 31.18 | 31.18 | 16.39 | 16.39 | 16.48 | 16.48 | 16.48 | 16.48 | 16.48 | 16.48 | 16.48 | 16.48 |
| | Training | 0.18 | 0.18 | 0.18 | 0.18 | 0.18 | 0.18 | 0.18 | 0.18 | 0.18 | 0.18 | 0.18 | 0.18 |
| | Supply chain | 11.85 | 11.85 | 63.35 | 63.35 | 33.63 | 33.63 | 54.69 | 54.69 | 16.73 | 16.73 | 47.58 | 47.58 |
| **Processing a sample (NPA, stool, GA or sputum) and conducting Xpert MTB/RIF Ultra test** | **Total** | **1590.43** | **3432.14** | **1764.32** | **3673.50** | **1683.82** | **3625.62** | **1711.84** | **3611.37** | **1599.15** | **3551.50** | **1721.16** | **3599.87** |
| | Personnel | 45.45 | 45.45 | 55.32 | 55.32 | 66.54 | 66.54 | 35.43 | 35.43 | 35.43 | 35.43 | 68.68 | 68.68 |
| | Equipment | 266.32 | 2067.70 | 266.32 | 2065.17 | 266.32 | 2112.70 | 266.32 | 2094.96 | 266.32 | 2139.82 | 266.32 | 2089.38 |
| | Supplies | 1230.71 | 1230.71 | 1211.20 | 1211.20 | 1221.15 | 1221.15 | 1214.07 | 1214.07 | 1226.74 | 1226.74 | 1216.40 | 1216.40 |
| | Training | 1.69 | 1.69 | 1.69 | 1.69 | 1.69 | 1.69 | 1.69 | 1.69 | 1.69 | 1.69 | 1.69 | 1.69 |
| | Supply chain | 46.26 | 86.59 | 229.79 | 340.12 | 128.12 | 223.54 | 194.33 | 265.22 | 68.97 | 147.82 | 168.07 | 223.72 |
| **Conducting smear microscopy (any sample type)** | **Total** | **31.29** | **31.29** | **85.18** | **85.18** | **58.49** | **58.49** | **71.54** | **71.54** | **33.34** | **33.34** | **73.37** | **73.37** |
| | Personnel | 10.88 | 10.88 | 13.51 | 13.51 | 16.51 | 16.51 | 8.21 | 8.21 | 8.21 | 8.21 | 17.08 | 17.08 |
| | Supplies | 7.10 | 7.10 | 7.10 | 7.10 | 7.18 | 7.18 | 7.18 | 7.18 | 7.18 | 7.18 | 7.18 | 7.18 |
| | Training | 1.69 | 1.69 | 1.69 | 1.69 | 1.69 | 1.69 | 1.69 | 1.69 | 1.69 | 1.69 | 1.69 | 1.69 |
| | Supply chain | 11.62 | 11.62 | 62.88 | 62.88 | 33.12 | 33.12 | 54.46 | 54.46 | 16.26 | 16.26 | 47.42 | 47.42 |
| **Conducting a digital chest X-ray** | **Total** | **747.67** | **n/a** | **777.11** | **n/a** | **843.99** | **n/a** | **771.05** | **n/a** | **848.61** | **n/a** | **808.84** | **n/a** |
| | Personnel | 67.05 | | 86.99 | | 155.89 | | 86.28 | | 162.77 | | 126.14 | |
| | Equipment | 674.60 | | 674.60 | | 674.60 | | 674.60 | | 674.60 | | 674.60 | |

| Activity | | Cambodia | | Cameroon | | Côte d'Ivoire | | Mozambique | | Sierra Leone | | Uganda | |
|---|---|---|---|---|---|---|---|---|---|---|---|---|---|
| | **Health facility** | **DH** | **PHC** | **DH** | **PHC** | **DH** | **PHC** | **DH** | **PHC** | **DH** | **PHC** | **DH** | **PHC** |
| | Training | 0.54 | | 0.54 | | 0.54 | | 0.54 | | 0.54 | | 0.54 | |
| | Supply chain | 5.48 | | 14.98 | | 12.96 | | 9.63 | | 10.71 | | 7.56 | |
| **Diagnosis and treatment of drug-sensitive tuberculosis** | **Total** | **3338.75** | **3338.75** | **3258.19** | **3258.19** | **4072.18** | **4072.18** | **3621.71** | **3621.71** | **4278.40** | **4278.40** | **3228.36** | **3228.36** |
| | Personnel | 394.34 | 394.34 | 601.28 | 601.28 | 1164.55 | 1164.55 | 700.90 | 700.90 | 1556.57 | 1556.57 | 586.90 | 586.90 |
| | Supplies - drugs | 2941.04 | 2941.04 | 2653.56 | 2653.56 | 2904.27 | 2904.27 | 2917.45 | 2917.45 | 2718.47 | 2718.47 | 2638.10 | 2638.10 |
| | Training | 3.36 | 3.36 | 3.36 | 3.36 | 3.36 | 3.36 | 3.36 | 3.36 | 3.36 | 3.36 | 3.36 | 3.36 |
| **Diagnosis and treatment of multidrug-resistant tuberculosis** | **Total** | **144,102.67** | **144,102.67** | **146,519.91** | **146,519.91** | **150,619.64** | **150,619.64** | **146,763.33** | **146,763.33** | **154,494.34** | **154,494.34** | **146,677.59** | **146,677.59** |
| | Personnel | 3108.41 | 3108.41 | 4739.53 | 4739.53 | 9179.52 | 9179.52 | 5524.80 | 5524.80 | 12269.60 | 12269.60 | 4626.22 | 4626.22 |
| | Supplies - drugs | 140,967.76 | 140,967.76 | 141,753.88 | 141,753.88 | 141,413.62 | 141,413.62 | 141,212.03 | 141,212.03 | 142,198.25 | 142,198.25 | 142,024.87 | 142,024.87 |
| | Training | 26.50 | 26.50 | 26.50 | 26.50 | 26.50 | 26.50 | 26.50 | 26.50 | 26.50 | 26.50 | 26.50 | 26.50 |

DH: district hospital, PHC: primary health centre, NPA: nasopharyngeal aspirate, GA: gastric aspirate.

#### Supplementary results - Time and motion study

To be able to value the contribution of labour, it was necessary to understand the length of time that staff spend undertaking each task that contributed to the care of a TB-Speed patient. We carried out a time and motion study involving healthcare workers (HCWs). HCWs were invited to take part voluntarily. The time and motion study was nested in the main study protocol approved by the WHO and the sponsor's (Inserm) ethics review committees, as well as the country ethics committees. No personal data related to the HCWs or patients was collected. Research assistants in each country explained the time and motion study to HCWs and ensured that the data were recorded correctly. Each HCW self-completed a timesheet on each of five working days which included some activities related to treating TB-Speed study participants. It was noted that all HCWs were seeing a mixture of TB-Speed participants and other patients, with study participants usually being in a minority. On the timesheet they recorded all activities related to TB-Speed patients and the length of their time they spent on each activity. In the six countries, across tertiary/district hospitals and primary health centres, we recruited 101 nurses/primary care clinicians, 36 doctors, 24 radiographers, and 18 laboratory technicians. Timesheets were used to record the length of tuberculosis-related consultations, appointments and diagnostic tests, but were not used to measure the length of care for inpatients. Inpatient care was valued using standard national costs for bed days, calculated by WHO-CHOICE. Study results are presented in **Table 2**.

**Table 2.** Length of TB-Speed activities (in minutes)

| NURSES/PRIMARY CARE CLINICIANS | | | | |
|---|---|---|---|---|
| **Activity** | **Number of episodes observed** | **Mean** | **SD** | **Median** |
| Systematic tuberculosis screening | 775 | 7 | 8 | 5 |
| Conduct - HIV rapid test | 35 | 16 | 11 | 10 |
| Conduct - Malaria rapid test | 170 | 10 | 5 | 10 |
| Detailed initial clinical examination and assessment | 339 | 15 | 14 | 10 |
| Patient measurements (height, weight, vital signs) | 556 | 11 | 9 | 9 |
| Sample collection - GA | 27 | 21 | 9 | 20 |
| Sample collection - NPA | 159 | 13 | 10 | 9 |
| Sample collection - sputum | 12 | 26 | 15 | 20 |

| | | | | |
|---|---|---|---|---|
| Sample collection - stool | 86 | 14 | 9 | 13 |
| Post-discharge follow-up appointment | 104 | 14 | 14 | 7 |
| Other appointment or consultation | 150 | 16 | 14 | 11 |
| DOCTORS | | | | |
| **Activity** | **Number of episodes observed** | **Mean** | **SD** | **Median** |
| Systematic tuberculosis screening | 67 | 13 | 9 | 10 |
| Detailed initial clinical examination and assessment | 161 | 19 | 18 | 10 |
| Chest X-ray - interpreting | 77 | 16 | 12 | 13 |
| Patient consultation to give results or discuss diagnosis | 108 | 16 | 13 | 11 |
| Post-discharge follow-up appointment | 54 | 21 | 18 | 15 |
| RADIOGRAPHERS | | | | |
| **Activity** | **Number of episodes observed** | **Mean** | **SD** | **Median** |
| Chest X-ray - conducting | 241 | 11 | 12 | 6 |
| Chest X-ray - interpreting | 25 | 9 | 4 | 9 |
| LABORATORY TECHNICIANS | | | | |
| **Activity** | **Number of episodes observed** | **Mean** | **SD** | **Median** |
| Receiving sample | 53 | 5 | 4 | 5 |
| Sorting sample | 41 | 6 | 4 | 6 |
| Technical preparation, initiating test | 65 | 23 | 13 | 20 |
| Recording and returning results | 49 | 7 | 5 | 5 |
| Validation of results | 35 | 5 | 5 | 5 |

SD: standard deviation, GA: gastric aspirate, NPA: nasopharyngeal aspirate

### Cost-effectiveness modelling

#### Overview of model design and implementation

##### *Development of a health economic model*

A decision analytic mathematical model was developed to assess the clinical benefits, cost-effectiveness, and budget impact of the intervention. We developed a ‘core’ model representing the structure of populations, health services and epidemiology in each country, and the natural history of treated and untreated tuberculosis, including adverse events. A decision module was then developed to add on to the core model to evaluate the cost-effectiveness and budget impact of the decentralisation interventions, and its structure was determined by patient care pathways.

The core model containing common evidence and functions for modelling tuberculosis natural history and outcomes was created and is being maintained using the *“HEdtree”* R package.[2] This model is based on previously published work, includes child age, sex, and HIV/antiretroviral treatment status, and models the natural history of progression to tuberculosis disease and tuberculosis treatment outcomes.[3–6]

##### *Development of patient care pathways*

To inform the design of the economic model, detailed patient care pathways were developed to represent the routes that patients would be expected to follow from the point at which tuberculosis screening or assessment could be initiated up to the end of treatment (Appendix Figures 1-4 above). These incorporated each episode and element of healthcare encountered by the patient, including the number of expected appointments, the diagnostic tests or screening undertaken, medications given and inpatient care received. They differentiated between the healthcare provided for those found to be positive or negative at each of the screening or diagnostic stages included, and were replicated for each intervention arms and the comparator standard-of-care arm.

The actual standard of care in current practice differed both between and within countries. In developing the standard-of-care pathways, the intention was to represent the expected typical current standard for care in TB-Speed countries as realistically as possible. The pathways formed the basis of the patient flows in the economic model, and allowed the resources required by patients receiving each intervention to be quantified and thereby costed.

##### *Resource use*

As this is a modelling study and not a within-trial analysis, resource use in the model was not based directly upon the actual resource use of each patient within the TB-Speed intervention study, but based on the resources expected to be required to follow each of the patient care pathways designed to structure the modelling. However, where the pathways corresponded to the experience of patients in TB-Speed studies we used actual resource use recorded by the study wherever possible, for example, annual usage of GeneXpert machines at DH and PHC levels was estimated based on observed data.

The patient care pathways lay out in detail the resources required at each stage of a patient's treatment journey, depending on the intervention they receive and their disease status. This includes the number of appointments expected; the number and nature of staff present at each appointment; the tests undertaken; and the medication given for tuberculosis. The pathways were developed by the health economists in an iterative process of consultation with HCWs involved in the study.

#### Cost-effectiveness analysis

The economic model generated projections for mortality which were used to calculate the expected overall health outcomes for patients following each alternative intervention arm in terms of disability-adjusted life years (DALYs), which are typically quantified as the sum of the years of life lost due to premature mortality in the population and the years lived with disability for people living with the health condition or its consequences.[7] In this case, the effects of morbidity on DALYs were not included in the model due to the short length of time children had tuberculosis compared to the lifetime horizon of the model. The economic model was constructed to take into account differences in the structure of the health system in each country. Life-table and cost data used in the model were also specific to each country. We did not assume any treatment effect beyond the end of treatment at individual level. We assumed that the observed intervention effects do not vary over time in projecting the discounted impacts of long-term deployment. The primary cost-effectiveness analyses compared the costs and health outcomes of each arm extrapolated to a lifetime horizon using mathematical modelling to capture the full benefit of any differences in mortality between alternative interventions. Both the costs and DALYs calculated in the extrapolation of the cost-effectiveness modelling were discounted at an annual rate of 3% as recommended by WHO-CHOICE and the iDSI Reference Case for Economic Evaluation.[8,9] (In practice all costs fell within 1 year, and so discounting was only relevant to DALYs.) The cost and DALY results from the model were combined to calculate incremental cost-effectiveness ratios (ICERs).

To determine if an intervention is likely to be cost-effective compared to the comparator strategy, ICERs can be compared against a benchmark known as a cost-effectiveness threshold for the relevant country.[8] However, cost-effectiveness thresholds are not always considered to inform decisions. At country level, the WHO-CHOICE methodology uses a health maximisation approach with a ranking of all packages of healthcare interventions from the most cost-effective intervention to the least cost-effective to assess value for money, and recommends their adoption until a 'budget' threshold is reached. Other economic aspects such as affordability and non-economic aspects (acceptability, feasibility or health system capacity, sustainability, scalability, equity, and ethics) may also be considered to inform decisions.

WHO 'demand side' thresholds, based on the WHO-CHOICE methodology, of 1 times GDP per capita or 3 times GDP per capita,[9] are recognised as too high and locally irrelevant.[10] These thresholds are used as global, normative guidelines. For instance, these are used for defining WHO 'Best buys' interventions for the prevention and control of noncommunicable diseases.[11]

However, as these thresholds are not evidence-based, ICERs below these level do not guarantee that an intervention would in fact be affordable within a national health system.

'Supply side' thresholds may be defined by a public health system as the maximum financial investment a public payer will commit to generate a unit of health.[12] To inform national TB budget allocation by local decision makers, we assess the cost-effectiveness of the TB-Speed Decentralization intervention using locally adapted supply-side thresholds. Given that there are not official thresholds for the countries covered by this study, and there are a range of approaches that can be considered, we compared our study findings with alternative sources estimating cost-effectiveness thresholds.[10] Ochalek et al. have estimated country level health opportunity costs (that is, what is given up as a consequence of introducing an intervention) and provided ranges of cost-effectiveness thresholds,[13] whereas Woods et al. extrapolated health opportunity costs in low- and middle-income countries from United Kingdom estimates.[14] We present and compare our ICERs to these two approaches to guide recommendations on the interventions' cost-effectiveness and how they compare with the current portfolio of interventions in these countries (**Table 10**). Both sources present a number of options for potential thresholds - we calculate the probability of the interventions being cost-effective compared to both the highest and lowest thresholds proposed by each source for each country, re-valued in line with GDP growth since publication. The presentation of two sets of thresholds aims to emphasise that the choice of cost-effectiveness threshold is not prescribed in these countries. Ultimately, the choice of cost-effectiveness threshold is for local decision makers to make when considering which interventions are cost-effective, and which intervention should be adopted in national health plans.

#### Modelling of diagnostic accuracy

The results of bacteriological tests are considered positive if it is possible to obtain a diagnostic sample *and* the test results are positive (calculated as the product of the probabilities of these two events). The test positivity depends on child true tuberculosis status and the test accuracy, typically derived from systematic reviews with meta-analysis. The probability that a sample is available (feasible and accepted and successful) is based on literature (**Table 4**). If multiple diagnostic tests are employed (for example, Xpert Ultra on an NPA sample and Xpert Ultra on a stool sample), a result is positive if *either* test is positive.

#### Model parameters

Parameters used in the model fall into a number of categories:

1. Based on literature, expert opinion, or non-cascade study data
    a. Non-cascade study data & opinion (**Table 3**)
    b. Diagnostic accuracy from literature (**Table 4**)
    c. Outcomes parameters from literature (**Table 5**)
2. Directly based on study cascade through meta-analysis
3. Based on cascade data via back-calculation

This section presents parameters under category 1 above. Cascade data are discussed in the following section.

**Table 3.** Parameters derived from study data and expert opinion

| NAME | MEDIAN (IQR) | DISTRIBUTION | DESCRIPTION |
|---|---|---|---|
| d.ptltfu | 0.046 (0.031 - 0.064) | B(3.6,68.4) | pre-treatment loss to follow-up |
| d.ptltfu.soc | 0.180 (0.107 - 0.272) | 0.15 | pre-treatment loss to follow-up |
| d.ptltfu.idh | 0.180 (0.107 - 0.272) | 0.13 | pre-treatment loss to follow-up |
| d.ptltfu.iph | 0.180 (0.107 - 0.272) | 0.25 | pre-treatment loss to follow-up |
| d.idh.dh.7dltfu | 0.180 (0.107 - 0.272) | B(2,8) | DH-focused intervention, at DH: 7-day loss to follow-up, patients invited back for reassessment 7 days later (having initially been assessed as not having tuberculosis) |
| d.iph.dh.7dltfu | 0.092 (0.063 - 0.129) | B(2,8) | PHC-focused intervention, at DH: 7-day loss to follow-up, patients invited back for reassessment 7 days later (having initially been assessed as not having tuberculosis) |
| d.iph.phc.7dltfu | 0.092 (0.062 - 0.130) | B(2,8) | PHC-focused intervention, at PHC: 7-day loss to follow-up, patients invited back for reassessment 7 days later (having initially been assessed as not having tuberculosis) |
| d.soc.dh.test.o5 | 0.092 (0.063 - 0.129) | B(4.48,10.45) | SOC, at DH: children 5-14 years receiving Xpert Ultra testing [either sputum or GA], in those identified as having presumptive tuberculosis |
| d.soc.dh.test.u5 | 0.240 (0.176 - 0.314) | B(3.36,30.22) | SOC, at DH: children 0-4 years receiving Xpert Ultra testing [on GA], in those identified as having presumptive tuberculosis |
| d.soc.dh.fracsp.o5 | 0.397 (0.329 - 0.468) | B(24.63,12.31) | SOC, at DH: children 5-14 years receiving Xpert Ultra on a sputum sample, of those receiving any Xpert test [the remainder receiving it on a GA sample] |
| d.soc.phc.test.o5 | 0.092 (0.063 - 0.129) | B(3.5,31.5) | SOC, at PHC: children 5-14 years receiving Xpert Ultra testing [on sputum], in those identified as having presumptive tuberculosis |
| ES.poss.phc.o5 | 0.092 (0.063 - 0.129) | B(4.4375,13.3125) | % of children 5-14 years identified as having presumptive tuberculosis who give an expectorated sputum sample |
| ES.poss.dh.o5 | 0.139 (0.094 - 0.195) | B(9.2,13.8) | % of children 5-14 years identified as having presumptive tuberculosis who give an expectorated sputum sample |
| GA.poss.dh.o5 | 0.193 (0.145 - 0.247) | B(3.5,31.5) | % of children 5-14 years identified as having presumptive tuberculosis who give a GA sample |
| GA.poss.dh.u5 | 0.617 (0.561 - 0.672) | B(3.5,31.5) | % of children 0-4 years identified as having presumptive tuberculosis who give a GA sample |
| d.soc.phc.smear.o5 | 0.986 (0.982 - 0.989) | B(3.25,18.4166666666667) | % of children 5-14 years identified as having presumptive tuberculosis who receive smear microscopy on a sputum sample |
| d.soc.dh.smear.o5 | 0.456 (0.397 - 0.516) | B(5.48888888888889,21.9555555555556) | % of children 5-14 years identified as having presumptive tuberculosis who receive smear microscopy on a sputum sample |

| | | | |
|---|---|---|---|
| NPA.poss.dh | 0.978 (0.967 - 0.985) | 0.93 | % patients in whom collecting an NPA sample is possible |
| NPA.poss.phc | 0.736 (0.675 - 0.791) | 0.98 | % patients in whom collecting an NPA sample is possible |
| ST.poss.phc | 0.982 (0.976 - 0.988) | 0.61 | % patients in whom collecting a stool sample possible |
| ST.poss.dh | 0.729 (0.703 - 0.754) | 0.77 | % patients in whom collecting a stool sample possible |
| prr | 0.01 | 0.01 | prevalence of rifampicin resistance in people with tuberculosis |
| d.hivprev.u5 | 0.180 (0.107 - 0.272) | B(2,8) | HIV prevalence, children 0-4 years |
| d.hivprev.o5 | 0.180 (0.107 - 0.272) | B(2,8) | HIV prevalence, children 5-14 years |
| d.artcov | 0.820 (0.728 - 0.893) | B(8,2) | ART coverage |

**Table 4.** Diagnostic accuracy parameters based on literature

| NAME | MEDIAN (IQR) | DISTRIBUTION | DESCRIPTION | SOURCE |
|---|---|---|---|---|
| sens.xstool | 0.976 (0.971 - 0.980) | B(21.716911549042, 13.5951397502133) | sensitivity of Xpert Ultra on stool in bac+ children | Kay et al, 2020[15] |
| spec.xstool | 0.627 (0.615 - 0.639) | B(480.118305785123,7.31144628099173) | specificity of Xpert Ultra stool in bac+ children | Kay et al, 2020[15] |
| sens.xnpa | 0.627 (0.615 - 0.639) | B(14.876289210384, 17.67576595457) | sensitivity of Xpert Ultra on NPA in bac+ children | Kay et al, 2020[15] |
| spec.xnpa | 0.901 (0.894 - 0.908) | B(120.278188775511,3.08405612244899) | specificity of Xpert Ultra on NPA in bac+ children | Kay et al, 2020[15] |
| sens.xga | 0.019 (0.012 - 0.029) | B(19.4209751059137,7.1831003816393) | sensitivity of Xpert on GA in bac+ children | Kay et al, 2020[15] |
| spec.xga | 0.008 (0.006 - 0.011) | B(212.720785244704,4.11997443389335) | specificity of Xpert on GA in bac+ children | Kay et al, 2020[15] |
| sens.xsputum | 0.436 (0.413 - 0.460) | B(103.163282735012,38.5445232196748) | sensitivity for C+ of Xpert on sputum | Kay et al, 2020[15] |
| spec.xsputum | 0.149 (0.137 - 0.162) | B(520.629938271604,13.3494855967078) | specificity for C+ of Xpert on sputum | Kay et al, 2020[15] |
| sens.clin | 0.877 (0.852 - 0.899) | B(492.167344045368,292.788547574039) | SOC, at DH/PHC: sensitivity of clinical diagnosis without CXR | Marais et al, 2006[16] |

**Table 5.** Outcome parameters from literature

| NAME | MEDIAN (IQR) | DISTRIBUTION | DESCRIPTION | SOURCE |
|---|---|---|---|---|

| | | | | |
|---|---|---|---|---|
| ontx.u5 | 0.019 (0.012 - 0.029) | LN( -3.963316,0.6457913) | CFR children 0-4 on TB treatment (HIV-) | Jenkins et al 2017[17] |
| ontx.o5 | 0.008 (0.006 - 0.011) | LN(-4.828314,0.4817445) | CFR children 5-14 on TB treatment (HIV-) | Jenkins et al 2017[17] |
| hivartOR:mn | | MVN: [2.6375681, -0.5683867] | ORs of death on TB treatment, (OR HIV+ vs -) x (ART -/+): mean | Jenkins et al 2017[17], Dodd et al 2017[5] |
| hivartOR:sg | | MVN: [[0.2325509,-0.2325509],[-0.2325509,0.6367345]] | ORs of death on TB treatment, (OR HIV+ vs -) x (ART -/+): variance | Jenkins et al 2017[17] Dodd et al 2017[5] |
| notx.u5 | 0.436 (0.413 - 0.460) | LN(-0.830113,0.08035318) | CFR children 0-4 without TB treatment (HIV-) | Jenkins et al 2017[17] |
| notx.o5 | 0.149 (0.137 - 0.162) | LN(-1.903809,0.1285165) | CFR children 5-14 without TB treatment (HIV-) | Jenkins et al 2017[17] |
| notxH.u5 | 0.877 (0.852 - 0.899) | B(77.13050,11.10817) | CFR children 0-4 without TB treatment (HIV+/ART-) | Dodd et al 2017[5] |
| notxH.o5 | 0.746 (0.686 - 0.800) | B(19.59083,6.89700) | CFR children 5-14 without TB treatment (HIV+/ART-) | Dodd et al 2017[5] |
| notxHA.u5 | 0.542 (0.478 - 0.605) | B(15.18683,12.87500) | CFR children 0-4 without TB treatment (HIV+/ART+) | Dodd et al 2017[5] |
| notxHA.o5 | 0.484 (0.412 - 0.558) | B(10.43383,11.08417) | CFR children 5-14 without TB treatment (HIV+/ART+) | Dodd et al 2017[5] |

#### Calibration to cascade data

##### *Overview*

A number of parameters were calculated from data on cascades. Cascades were conceptualised as 4 stages (**Table 6**), each of which could occur at either PHC or DH level, with referral between them.

**Table 6**. Cascade of tuberculosis care stages

| A | B | C | D |
|---|---|---|---|
| Initial presentation | Screened | Presumptive TB | Treated for TB |

Count data from each stage and country were used to compute meta-analytic ratios, and a number of model parameters were derived from these ratios or other meta-analysed quantities. In particular, the parameters involved in these quantities are presented in **Table 7**.

**Table 7**. Parameters used for the calibration to cascade data

| Symbol | Meaning | Notes |
|---|---|---|
| $f$ | Fraction of care-seeking cohort age 0-4 years | Direct calculation |
| $p_p$ | Fraction of children initially going to PHC | Direct calculation, corrected by the fraction of PHCs in TB-Speed |
| $\pi_l^x$ | True TB prevalence, level $l$, stage $x$ | Ultimately used as prevalence by age at step A & an OR of going to DH \| TB.<br>Jointly sampled at D with $\phi$ then back-calculated |
| $\alpha$ | Coverage of screening | Direct calculation |
| $\sigma$ | Specificity of screening | Back-calculated |
| $\rho$ | Referral rate from PHC → DH, step $B \rightarrow C$ | Direct calculation |
| $\lambda$ | LTFU in the above referral | Direct calculation, making use of refusal data |
| $\phi$ | 1-specificity of clinical diagnosis | Jointly sampled with $\pi_l^D$ |
| $se$ | Sensitivity of diagnosis | These are calculated for the relevant algorithm $\phi$ given using the model |
| $sp$ | Specificity of diagnosis | |

*Assumptions*

A number of assumptions were used in order to perform these calculations:

1. Screening is 100% sensitive
2. Screening is random with respect to true tuberculosis status

Assumption 1 is essentially a definition: that the only type of tuberculosis we are modelling is that which can be detected by a systematic screen. The intervention at this step therefore increases the coverage of systematic screening, and tuberculosis that cannot be detected by systematic screening is neglected as not changing under intervention (and therefore cancels out from incremental comparisons).

Assumption 2 means that this screen is not more likely to be applied to those with true tuberculosis. The fraction of those screened who are considered to have presumptive tuberculosis is therefore determined by the specificity of screening and the prevalence of true tuberculosis among those at the screening step.

*Meta-analyses*

In all our cascade calculations, we use samples from the meta-analytic posteriors for relevant ratios rather than raw numbers because these better reflect across-country summary values and heterogeneity. Data were inspected and where intervention arm or age did not have an evident influence, we produced aggregate estimates. We used Bayesian hierarchical models with logit link functions fitted using Stan via the brms package (using default priors) - the Bayesian analogue of a GLMM meta-analysis with random effects for country. Samples from the posteriors were used in PSA-based calculations. Below we present forest plots for these analyses and a table of the parameters whose values were derived from the meta-analyses directly.

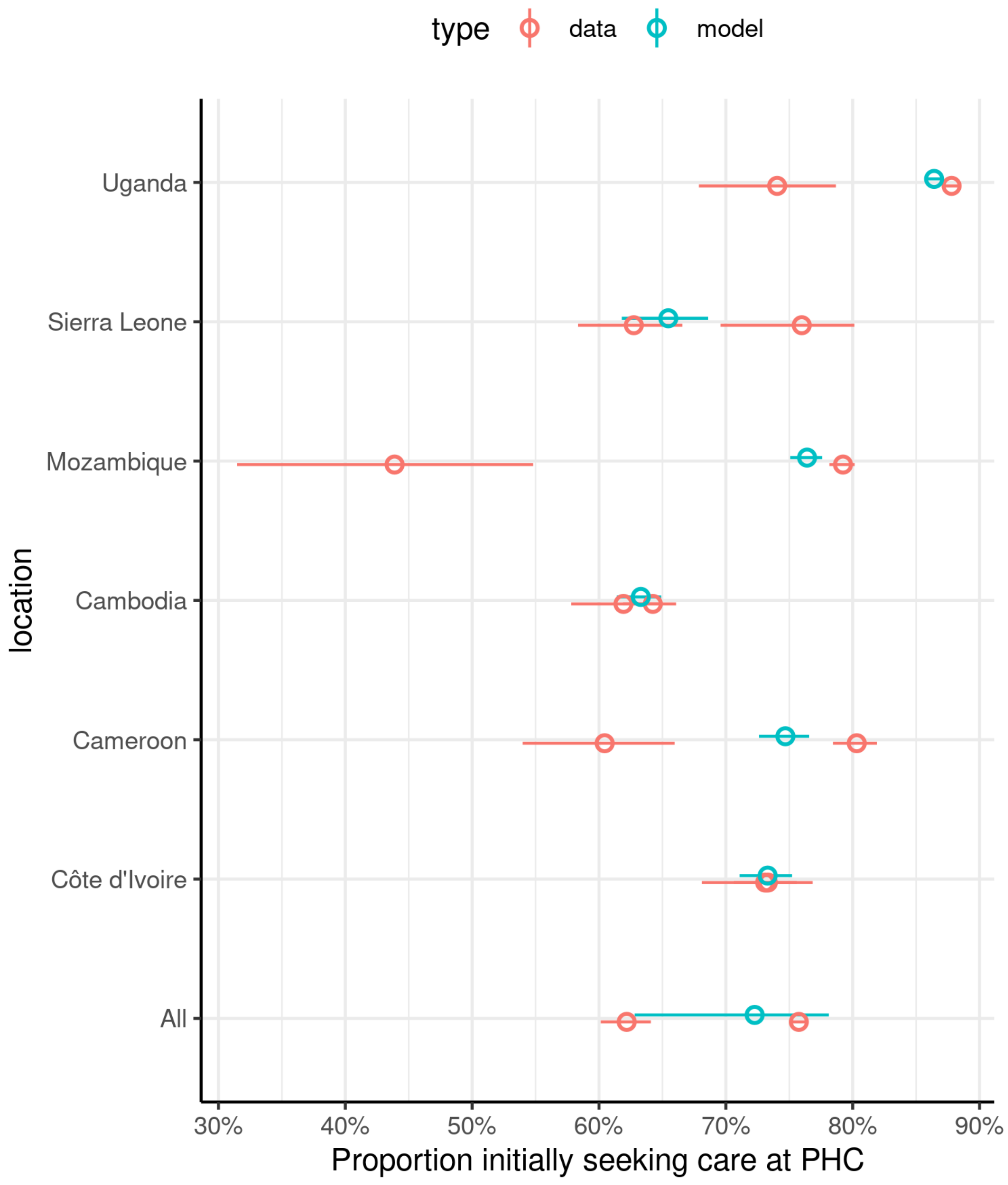


**Figure 8**. Initial care-seeking location corrected for fraction of district PHCs enrolled in TB-Speed. The two red data points for each location represent 0-4 year and 5-14 year age groups separately.

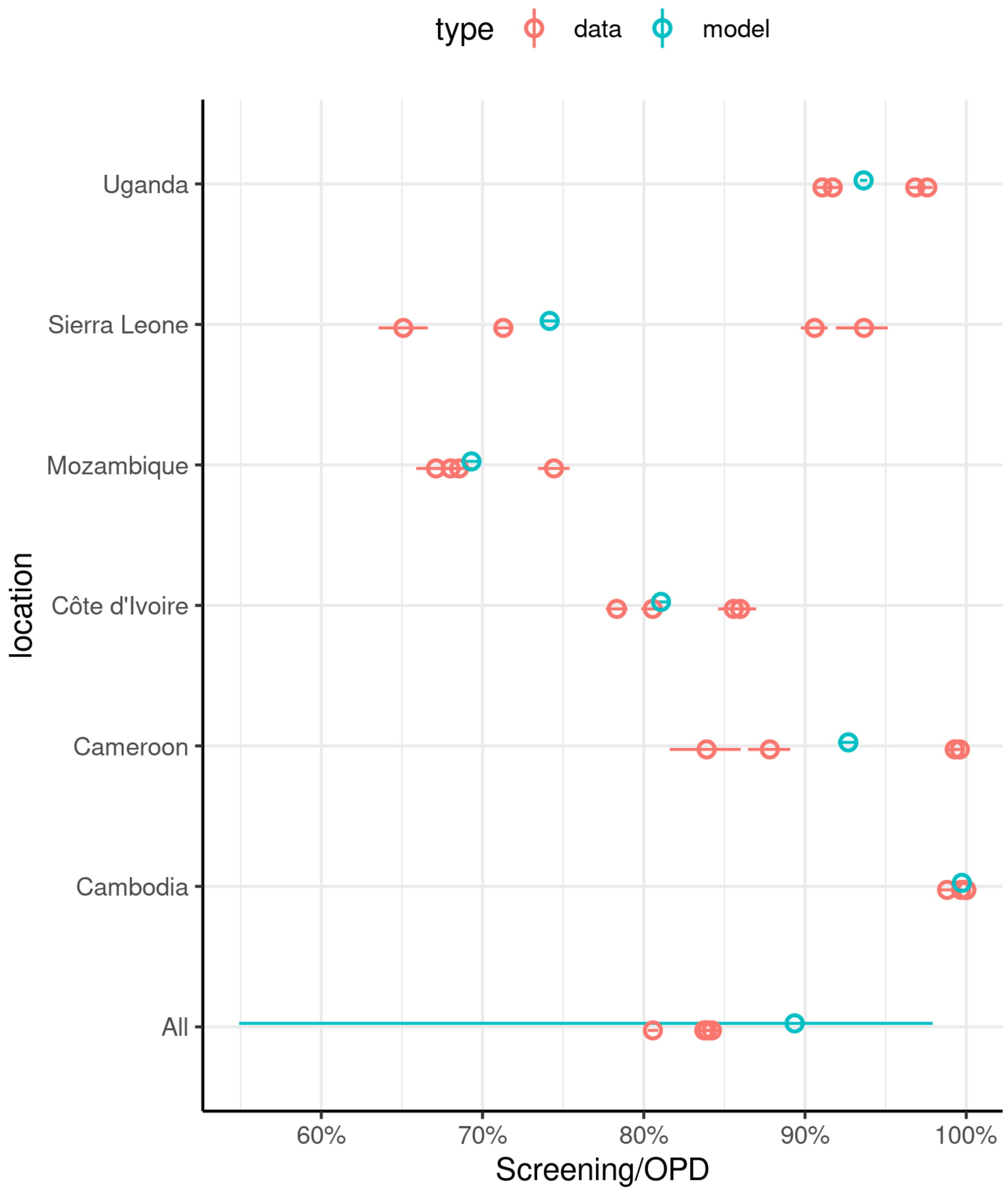


**Figure 9**. Meta-analysis of intervention screening coverage as a proportion of children presenting as outpatients (B/A). The four red data points for each location represent 0-4 year and 5-14 year age groups and both arms separately.

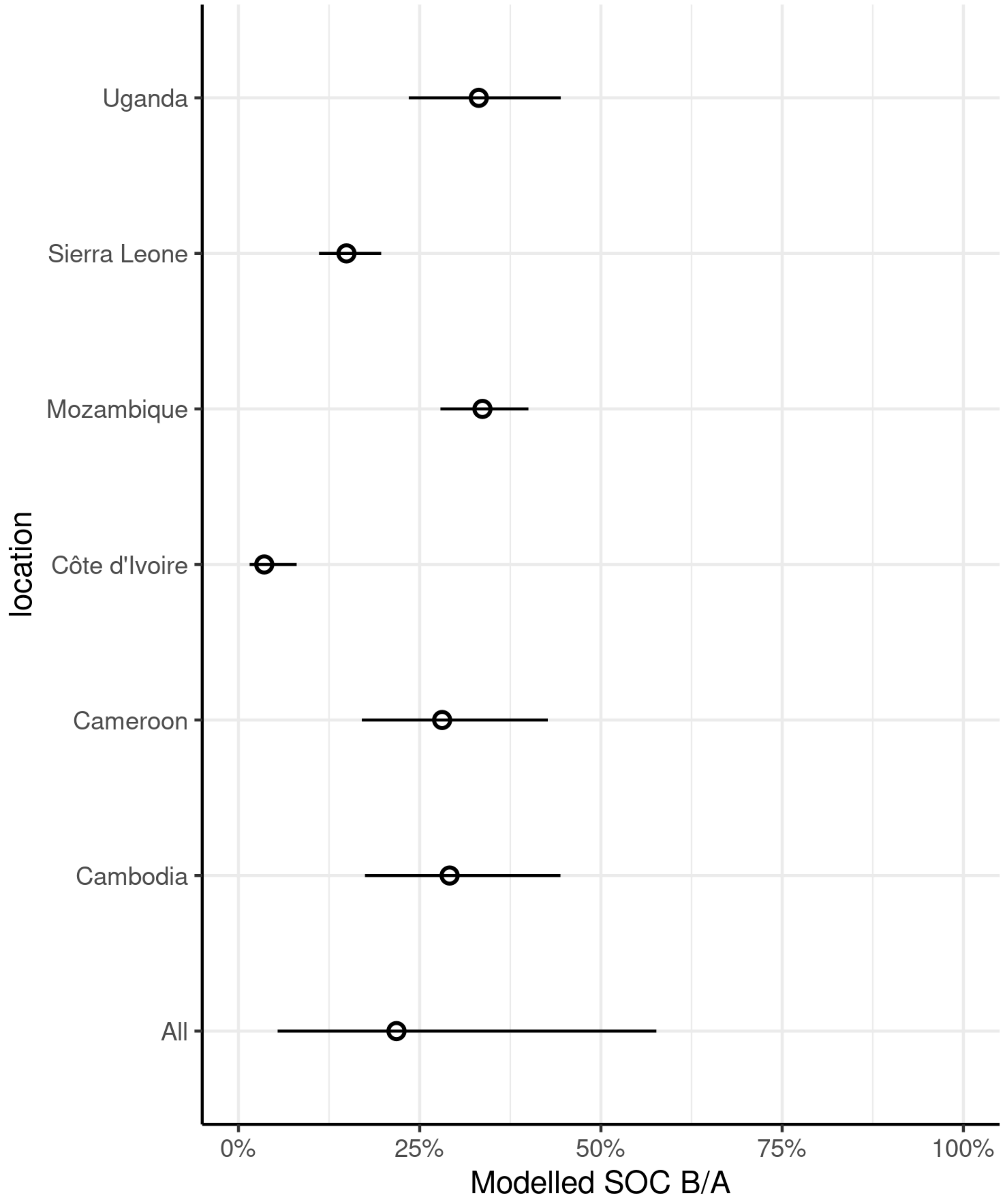


**Figure 10**. Standard of care screening coverage (B/A) modelled from the ratio of treatment to OPD attendances

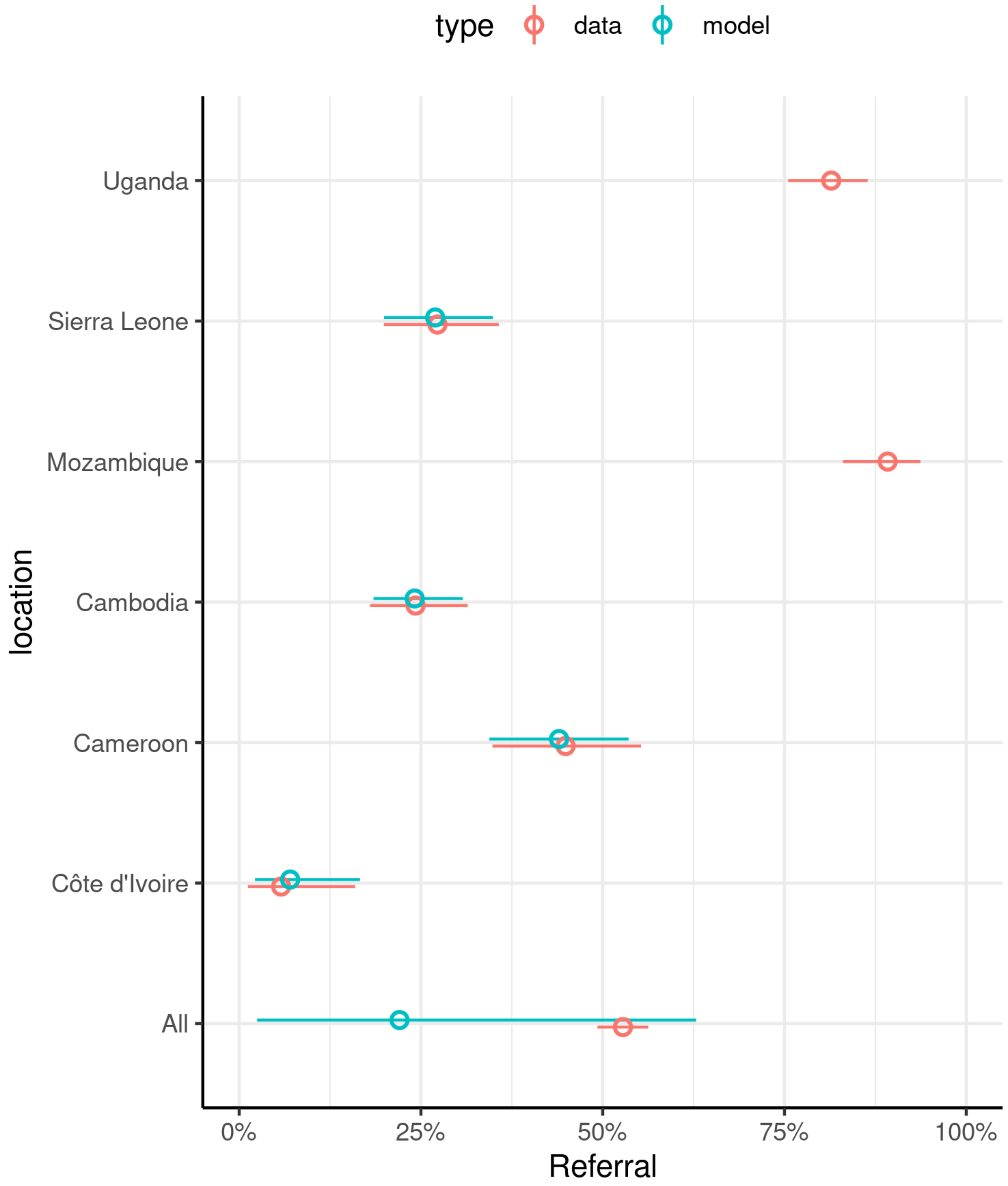


**Figure 11**. Percentage of children non-enrolled in the study at DH (DH-focused strategy), among children first presenting to PHC, then referred and identified with presumptive TB at DH (non-enrolment explained by either refusal or loss to follow-up during referral). Estimate excludes Mozambique and Uganda which had TB care services partially decentralised to PHC before the intervention.

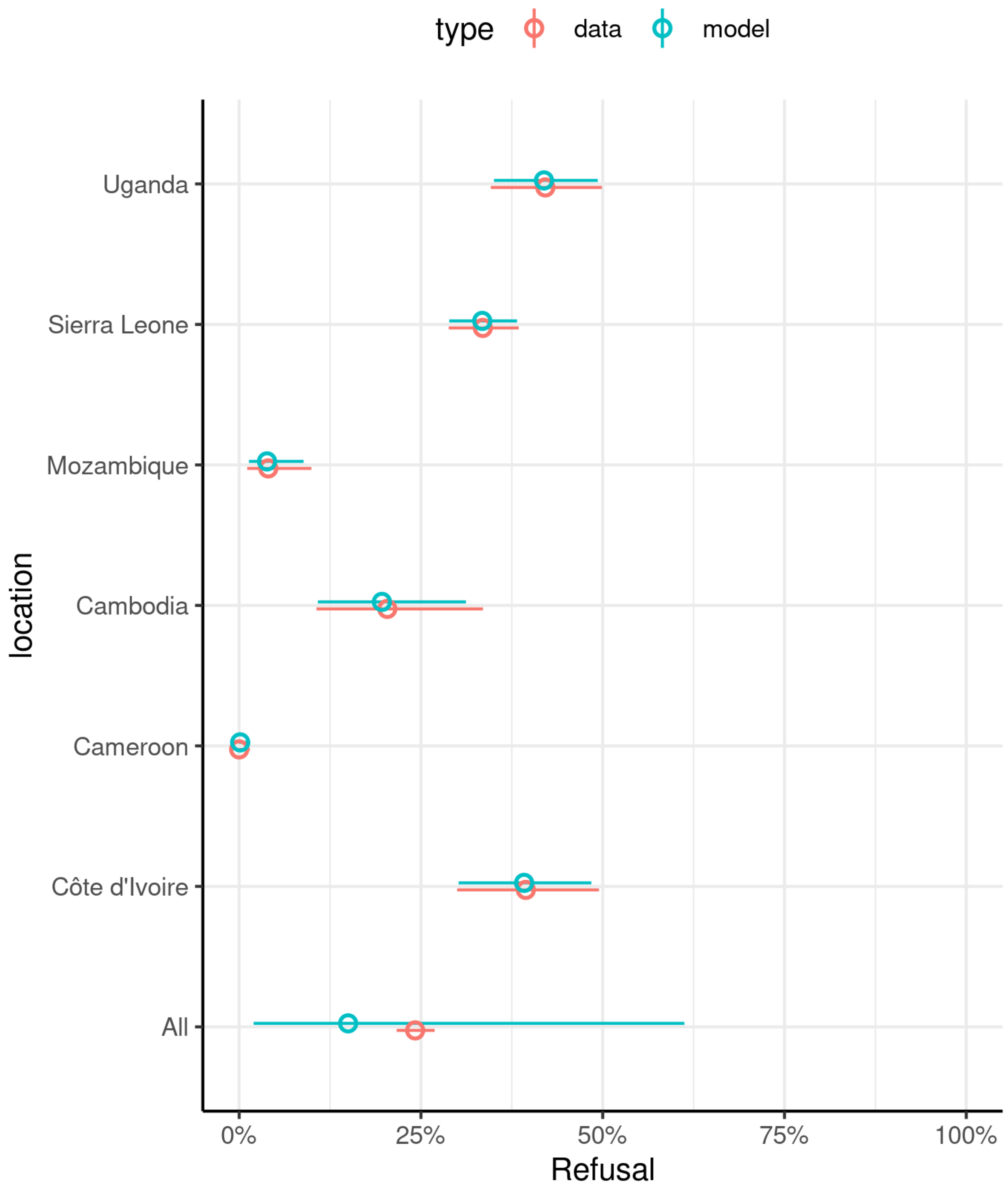


**Figure 12**. Percentage of children non-enrolled in the study at DH (DH-focused strategy), among children presenting to DH and identified with presumptive TB (refusal rate)

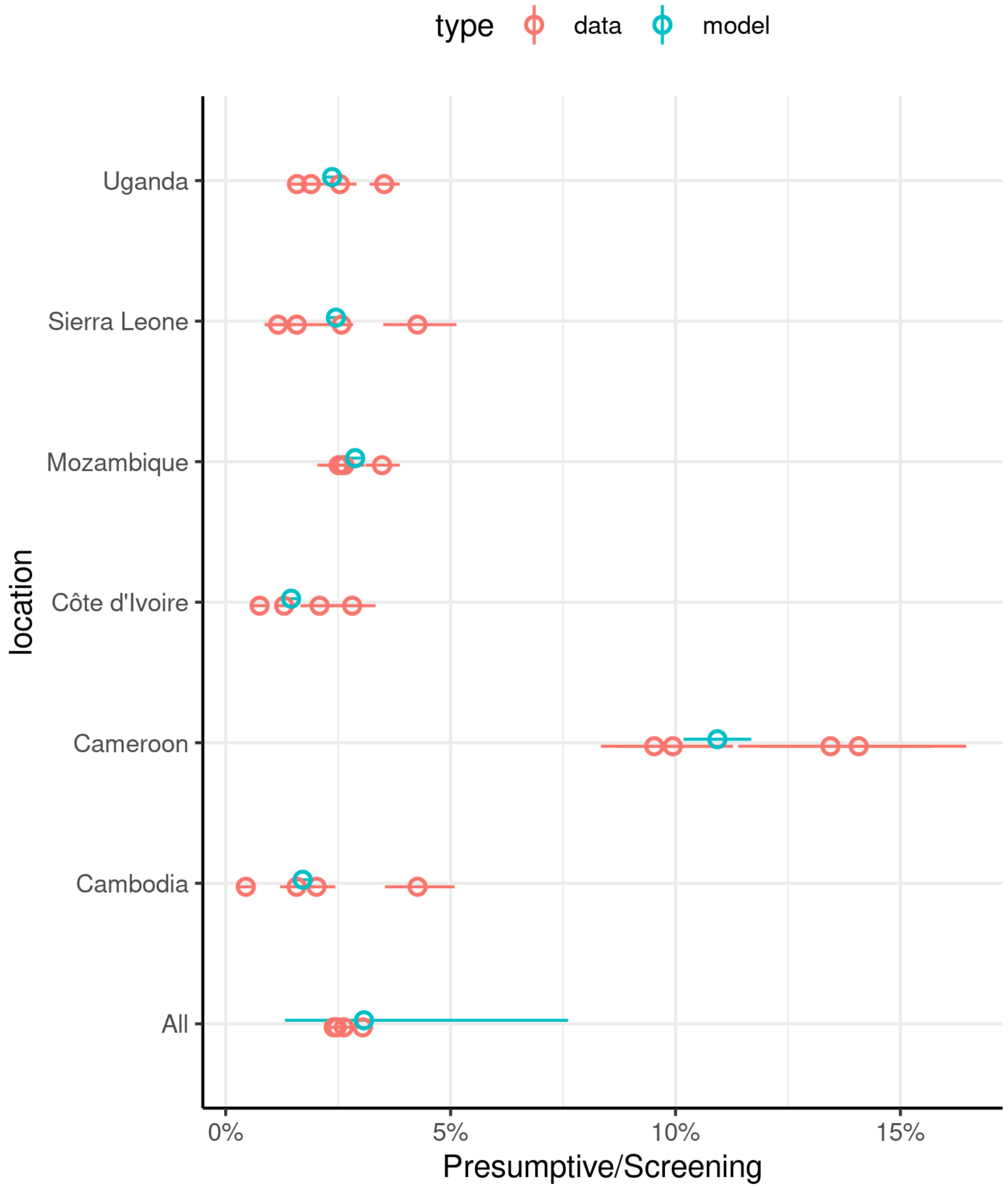


**Figure 13**. Presumptive tuberculosis identified among those screened (C/B). The four red data points for each location represent 0-4 year and 5-14 year age groups and both arms separately.

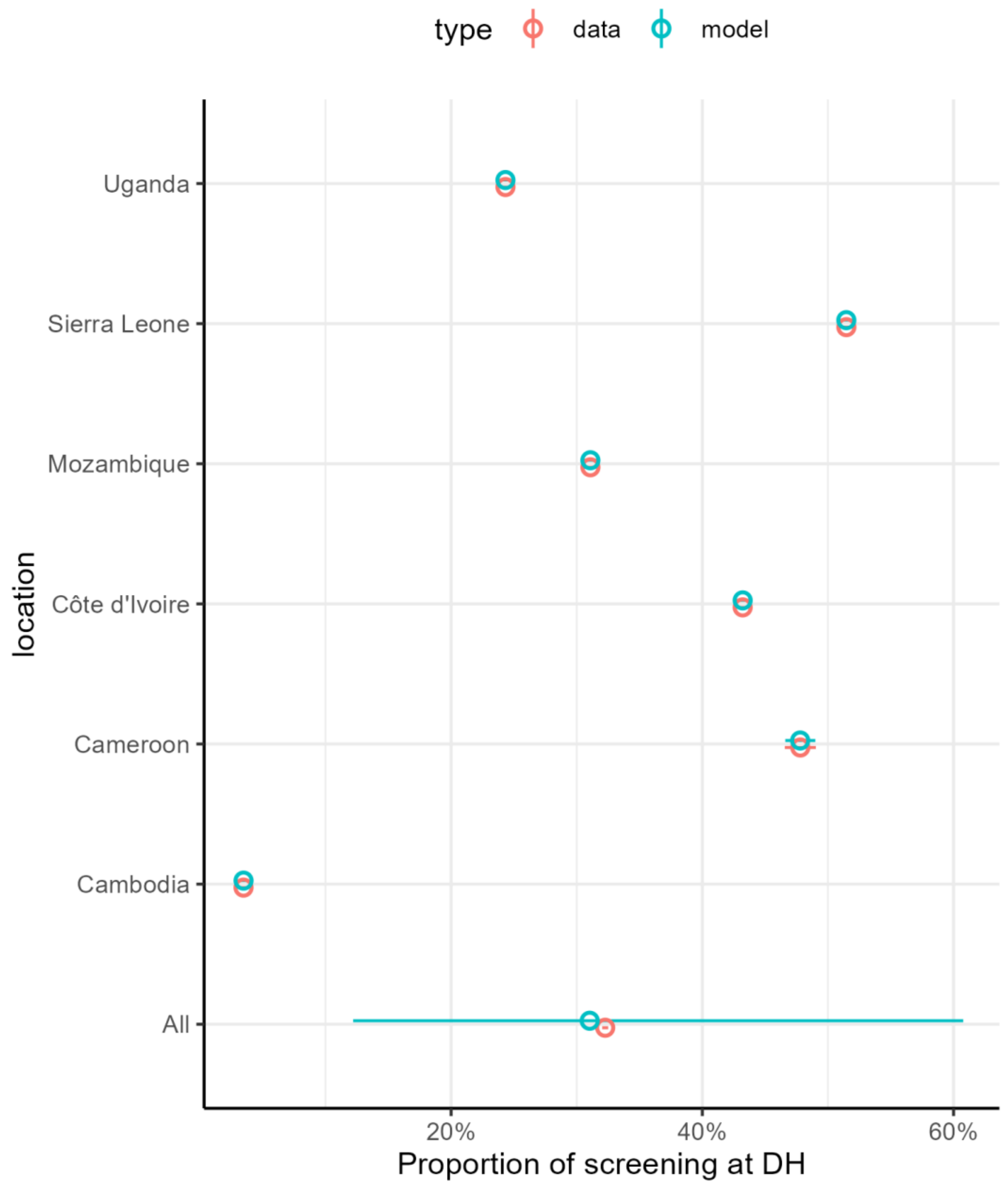


**Figure 14**. Proportion of screening percentage at DH compared to at PHC

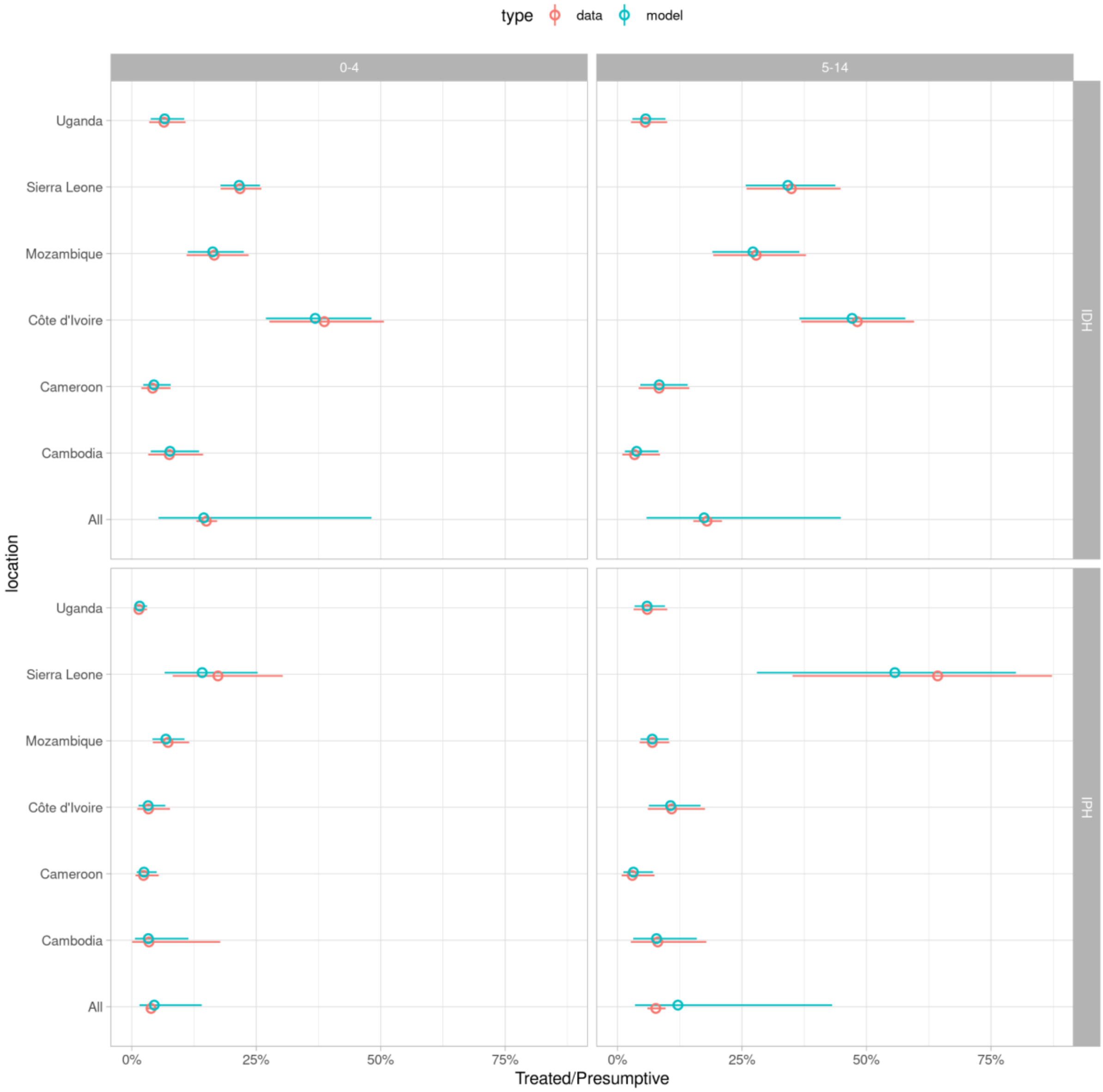


**Figure 15**. Proportion of children with presumptive tuberculosis receiving treatment (D/C). IPH=PHC-focused intervention; IDH=DH-focused intervention.

**Table 8**. Summary of parameters derived from meta-analyses

| NAME | DISTRIBUTION | MEDIAN (IQR) | MEANING |
|---|---|---|---|
| F_refs | B(1.37676464440336,4.86620425503129) | 0.190 (0.099 - 0.313) | Referral rate |
| F_refu | B(0.685514761946353,3.88961039796218) | 0.100 (0.032 - 0.222) | Refusal/LTFU on referral |
| F_presume | B(3.52045097800522,110.986717086738) | 0.028 (0.019 - 0.040) | Rate of presumptive TB among screened |
| F_ICS | B(94.2722891948688,36.1700858789189) | 0.724 (0.697 - 0.750) | Initial care-seeking at PHC vs DH |
| F_alpha | B(6.33480324077261,0.732420871081868) | 0.932 (0.850 - 0.977) | Intervention screening coverage |
| F_SOC_BoA_modelled | B(1.8735091532484,6.72208835298635) | 0.196 (0.115 - 0.299) | SOC screening coverage |
| DoC_0-4 | B(1.86216860060048,39.4126932387174) | 0.038 (0.021 - 0.061) | TB treatment among those identified with presumptive TB, 0-4 years |
| DoC_5-14 | B(1.14342204200909,8.2913550595211) | 0.095 (0.044 - 0.172) | TB treatment among those identified with presumptive TB, 5-14 years |
| Fu5 | B(55.4207078389574,32.9755206793714) | 0.628 (0.593 - 0.662) | Fraction of initial care-seeking cohort who are 0-4 years |
| F_omega_flat | G(1.14021779377055,0.35183741762051) | 0.292 (0.131 - 0.555) | Ratio of screening at DH to PHC |

##### *Sketch of calculations*

In this section, we describe more formally how the assumptions above can be used to back-calculate the quantities of interest (from right to left in the table above; D → A ).

First, consider the situation without referral between PHC and DH. Suppressing indices specifying age,

$$\pi^C = \frac{D/C + sp - 1}{se + sp - 1}$$

We can then use

$$B(\pi^B + (1-\sigma)(1-\pi^B)) = C$$

to find

$$\pi^C = \frac{B}{C}\pi^B = \frac{\pi^B}{\pi^B + (1-\sigma)(1-\pi^B)}$$

which can be rewritten as

$$\sigma = 1 - \frac{(C/B - \pi^B)}{1-\pi^B} = 1 - \frac{C}{B}.\frac{1-\pi^C}{1-(C/B)\pi^C}$$

where we have used

$\pi^B = \frac{C}{B}\pi^C$.

Similarly,

$$\pi^A = \frac{B}{A}\pi^B$$

Which means we have solved for $\pi^A$ and $\sigma$, given the numbers at stages A, …, D.

##### *Joint sampling of prevalence and the specificity of clinical diagnosis*

The specificity of diagnostic algorithms is uncertain, and in sampling across uncertainty it is easy to generate parameter sets where the first equation in the previous section results in an impossible negative prevalence. We therefore jointly sample the specificity of clinical diagnosis and prevalence of true tuberculosis at step C, which avoids this situation.

In more detail, this means we perform back calculations at the level of PSA replicate rather than in the mean, and must calculate how the specificity of diagnostic algorithms depends on the specificity of clinical diagnosis (*clinspec*). The specificity is empirically very close to linear in its dependence on *clinspec*, so for each PSA replicate we calculate the specificity of the diagnostic algorithm (by setting true tuberculosis prevalence to zero) at two values of *clinspec*, which we then use as the basis of a linear interpolation. We then use a bivariate normal approximation to analytically generate posteriors for true tuberculosis prevalence and clinical specificity, conditional on the proportion of those presumed to have tuberculosis who are diagnosed with tuberculosis (and other parameter values). We used a prior of $N(10\%,3\%^2)$ for true tuberculosis prevalence among those presumed to have tuberculosis and a prior of $N(10\%,2.5\%^2)$ for *(1 - clinspec)*, informed by Marais et al.[16]

##### *Back-calculations with referral*

With referral, we have

$$C_d^+ = B_d^+ + B_p^+\rho(1-\lambda)$$

$$C_d^- = B_d^-(1-\sigma_d) + B_p^-(1-\sigma_p)\rho(1-\lambda)$$

$$C_p^+ = B_p^+(1-\rho)$$

$$C_p^- = B_p^-(1-\sigma_p)(1-\rho)$$

where +/- superscripts denote positive or negative for true tuberculosis.

This implies

$$\pi_p^C = \frac{\pi_p^B(1-\rho)}{\pi_p^B(1-\rho)+(1-\pi_p^B)(1-\sigma_p)(1-\rho)}$$

$$\pi_d^C = \frac{\omega\pi_d^B+\pi_p^B\rho(1-\lambda)}{\omega[\pi_d^B+(1-\pi_d^B)(1-\sigma_d)]+[\pi_p^B\rho(1-\lambda)+(1-\pi_p^B)(1-\sigma_p)\rho(1-\lambda)]}$$

where we have written $\omega = B_d/B_p$.

We can again solve for $\pi_p^B$, $\pi_d^B$, $\sigma_p$, and $\sigma_d$ with references to the first equations in this section as follows:

$$\pi_p^B = (C_p/B_p)\pi_p^C/(1-\rho)$$

$$\pi_d^B = (C_d/B_d)\pi_d^C - (C_p/B_p)\frac{\rho(1-\lambda)}{\omega(1-\rho)}\pi_p^C$$

and then for the specificities

$$\sigma_p = 1-(C_p/B_p)\frac{1-\pi_p^C}{(1-\pi_p^B)(1-\rho)}$$

$$\sigma_d = 1-(C_d/B_d)\frac{1-\pi_d^C}{1-\pi_d^B}+\frac{1-\pi_p^B}{1-\pi_d^B}(1-\sigma_p)\rho(1-\lambda)/\omega$$

Our assumptions imply

$$\pi_l^A = B/A\,\pi_l^B$$

The prevalence of true tuberculosis in those initially seeking care ($\pi$) is then:

$$\pi = \frac{\sum_l \quad \pi_l^A A_l}{\sum_l \quad A_l}$$

and we can also calculate odds ratios quantifying the tendency for children with true tuberculosis to be among those initially seeking care at DH- rather than PHC-level, thus enriching true tuberculosis prevalence at DHs.

##### *Derived cascade parameters*

**Table 9**. Summary of cascade parameters derived from meta-analyses

| NAME | MEAN (95% CrI) | DESCRIPTION |
|---|---|---|
| tbu5 | 107.9 (10.5 - 387.7) | True TB prevalence per 100K at initial care-seeking, 0-4 years |
| tbo5 | 212.6 (14.4 - 693.7) | True TB prevalence per 100K at initial care-seeking, 5-14 years |
| ORu5 | 10.8 (0.1 - 33.6) | OR for initial care-seeking at DH-level given true TB, 0-4 years |
| ORo5 | 2.1 (0.1 - 10.1) | OR for initial care-seeking at DH-level given true TB, 5-14 years |
| sigmap_iph_0-4 | 97 (93.1 - 99.3) | % specificity of TB presumption, intervention, PHC, 0-4 years |
| sigmap_iph_5-14 | 97.1 (93.3 - 99.3) | % specificity of TB presumption, intervention, PHC, 5-14 years |
| sigmap_soc_0-4 | 97 (93.1 - 99.3) | % specificity of TB presumption, SOC, PHC, 0-4 years |
| sigmap_soc_5-14 | 97.1 (93.3 - 99.3) | % specificity of TB presumption, SOC, PHC, 5-14 years |
| sigmad_iph_0-4 | 97.3 (93.4 - 99.5) | % specificity of TB presumption, intervention, PHC, 0-4 years |
| sigmad_iph_5-14 | 97.4 (93.8 - 99.6) | % specificity of TB presumption, intervention, DH, 5-14 years |
| sigmad_soc_0-4 | 97.3 (93.4 - 99.5) | % specificity of TB presumption, SOC, DH, 0-4 years |
| sigmad_soc_5-14 | 97.4 (93.8 - 99.6) | % specificity of TB presumption, SOC, DH, 5-14 years |
| clinspec | 89.9 (85.3 - 94.8) | % specificity of clinical TB diagnosis |

CHEERS 2022 - Checklist

| Topic | No. | Item | Location where item is reported |
|---|---|---|---|
| **Title** | | | |
| | 1 | Identify the study as an economic evaluation and specify the interventions being compared. | Title, Page 1 |
| **Abstract** | | | |
| | 2 | Provide a structured summary that highlights context, key methods, results, and alternative analyses. | Abstract, Page 2 |
| **Introduction** | | | |
| **Background and objectives** | 3 | Give the context for the study, the study question, and its practical relevance for decision making in policy or practice. | Introduction, Paragraphs 4-6 |
| **Methods** | | | |
| **Health economic analysis plan** | 4 | Indicate whether a health economic analysis plan was developed and where available. | Methods, Modelling approach |
| **Study population** | 5 | Describe characteristics of the study population (such as age range, demographics, socioeconomic, or clinical characteristics). | Methods, Modelling approach |
| **Setting and location** | 6 | Provide relevant contextual information that may influence findings. | Introduction, Fifth Paragraph |
| **Comparators** | 7 | Describe the interventions or strategies being compared and why chosen. | Introduction, Fifth Paragraph; Figure 1 and Appendix |
| **Perspective** | 8 | State the perspective(s) adopted by the study and why chosen. | Methods, Modelling approach |
| **Time horizon** | 9 | State the time horizon for the study and why appropriate. | Methods, Modelling approach |
| **Discount rate** | 10 | Report the discount rate(s) and reason chosen. | Methods, Costing and Modelling approach |
| **Selection of outcomes** | 11 | Describe what outcomes were used as the measure(s) of benefit(s) and harm(s). | Methods, Modelling approach |
| **Measurement of outcomes** | 12 | Describe how outcomes used to capture benefit(s) and harm(s) were measured. | Methods, Modelling approach |
| **Valuation of outcomes** | 13 | Describe the population and methods used to measure and value outcomes. | Methods, Modelling approach |
| **Measurement and valuation of resources and costs** | 14 | Describe how costs were valued. | Methods, Costing approach |

| | | | |
|---|---|---|---|
| **Currency, price date, and conversion** | 15 | Report the dates of the estimated resource quantities and unit costs, plus the currency and year of conversion. | Methods, Costing approach |
| **Rationale and description of model** | 16 | If modelling is used, describe in detail and why used. Report if the model is publicly available and where it can be accessed. | Methods, Patient pathways & Modelling approach |
| **Analytics and assumptions** | 17 | Describe any methods for analysing or statistically transforming data, any extrapolation methods, and approaches for validating any model used. | Methods, Modelling approach |
| **Characterising heterogeneity** | 18 | Describe any methods used for estimating how the results of the study vary for subgroups. | Methods, Modelling approach First paragraph |
| **Characterising distributional effects** | 19 | Describe how impacts are distributed across different individuals or adjustments made to reflect priority populations. | Methods , Last paragraph |
| **Characterising uncertainty** | 20 | Describe methods to characterise any sources of uncertainty in the analysis. | Appendix |
| **Approach to engagement with patients and others affected by the study** | 21 | Describe any approaches to engage patients or service recipients, the general public, communities, or stakeholders (such as clinicians or payers) in the design of the study. | Methods, Patient pathways |
| **Results** | | | |
| **Study parameters** | 22 | Report all analytic inputs (such as values, ranges, references) including uncertainty or distributional assumptions. | Appendix |
| **Summary of main results** | 23 | Report the mean values for the main categories of costs and outcomes of interest and summarise them in the most appropriate overall measure. | Results, Pages 13-14 |
| **Effect of uncertainty** | 24 | Describe how uncertainty about analytic judgments, inputs, or projections affect findings. Report the effect of choice of discount rate and time horizon, if applicable. | Results, Pages 13-14, Table 1 & Figure 1 |
| **Effect of engagement with patients and others affected by the study** | 25 | Report on any difference patient/service recipient, general public, community, or stakeholder involvement made to the approach or findings of the study | Methods, Patient pathways |
| **Discussion** | | | |
| **Study findings, limitations, generalisability, and current knowledge** | 26 | Report key findings, limitations, ethical or equity considerations not captured, and how these could affect patients, policy, or practice. | Discussion |
| **Other relevant information** | | | |
| **Source of funding** | 27 | Describe how the study was funded and any role of the funder in the identification, design, conduct, and reporting of the analysis | Abstract, Page 3 |

| | | | |
|---|---|---|---|
| **Conflicts of interest** | 28 | Report authors conflicts of interest according to journal or International Committee of Medical Journal Editors requirements. | End of manuscript |

*From:* Husereau D, Drummond M, Augustovski F, et al. Consolidated Health Economic Evaluation Reporting Standards 2022 (CHEERS 2022) Explanation and Elaboration: A Report of the ISPOR CHEERS II Good Practices Task Force. Value Health 2022;25. doi:10.1016/j.jval.2021.10.008

Country level probabilities of cost-effectiveness and cost-effectiveness thresholds

**Table 10.** Country level probabilities of cost-effectiveness and cost-effectiveness thresholds in cost per DALY averted (2021 US$)

| Country | Threshold and probability value | Threshold 1 (Ochalek, 2018) | | Threshold 2 (Woods, 2016) | | GDP per capita* |
|---|---|---|---|---|---|---|
| | | Low | High | Low | High | |
| Cambodia | Threshold (% GDP per capita) | 260 (16%) | 390 (24%) | 70 (4%) | 829 (51%) | 1,625 |
| | Probability of being cost-effective - DH-focused | 40% | 69% | 0% | 94% | |
| | Probability of being cost-effective - PHC-focused | 7% | 27% | 0% | 77% | |
| Cameroon | Threshold (% GDP per capita) | 150 (9%) | 200 (12%) | 52 (3%) | 699 (42%) | 1,667 |
| | Probability of being cost-effective - DH-focused | 2% | 10% | 0% | 85% | |
| | Probability of being cost-effective - PHC-focused | 0% | 0% | 0% | 55% | |
| Côte d’Ivoire | Threshold (% GDP per capita) | 382 (15%) | 484 (19%) | 82 (3%) | 987 (39%) | 2,549 |
| | Probability of being cost-effective - DH- | 49% | 65% | 0% | 92% | |

| | | | | | | |
|---|---|---|---|---|---|---|
| | focused | | | | | |
| | Probability of being cost-effective - PHC-focused | 11% | 26% | 0% | 73% | |
| Mozambique | Threshold (% GDP per capita) | 177 (36%) | 226 (46%) | 6 (1%) | 215 (44%) | 492 |
| | Probability of being cost-effective - DH-focused | 10% | 22% | 0% | 19% | |
| | Probability of being cost-effective - PHC-focused | 0% | 2% | 0% | 1% | |
| Sierra Leone | Threshold (% GDP per capita) | 77 (16%) | 101 (21%) | 16 (3%) | 296 (62%) | 480 |
| | Probability of being cost-effective - DH-focused | 0% | 1% | 0% | 39% | |
| | Probability of being cost-effective - PHC-focused | 0% | 0% | 0% | 6% | |
| Uganda | Threshold (% GDP per capita) | 150 (17%) | 194 (22%) | 12 (1%) | 316 (36%) | 884 |
| | Probability of being cost-effective - DH-focused | 3% | 11% | 0% | 42% | |
| | Probability of being cost-effective - PHC-focused | 0% | 0% | 0% | 8% | |

*given for reference, not as a suggested threshold option

Supplementary results - Screening to diagnosis cascades during the intervention period by decentralisation approach and age group

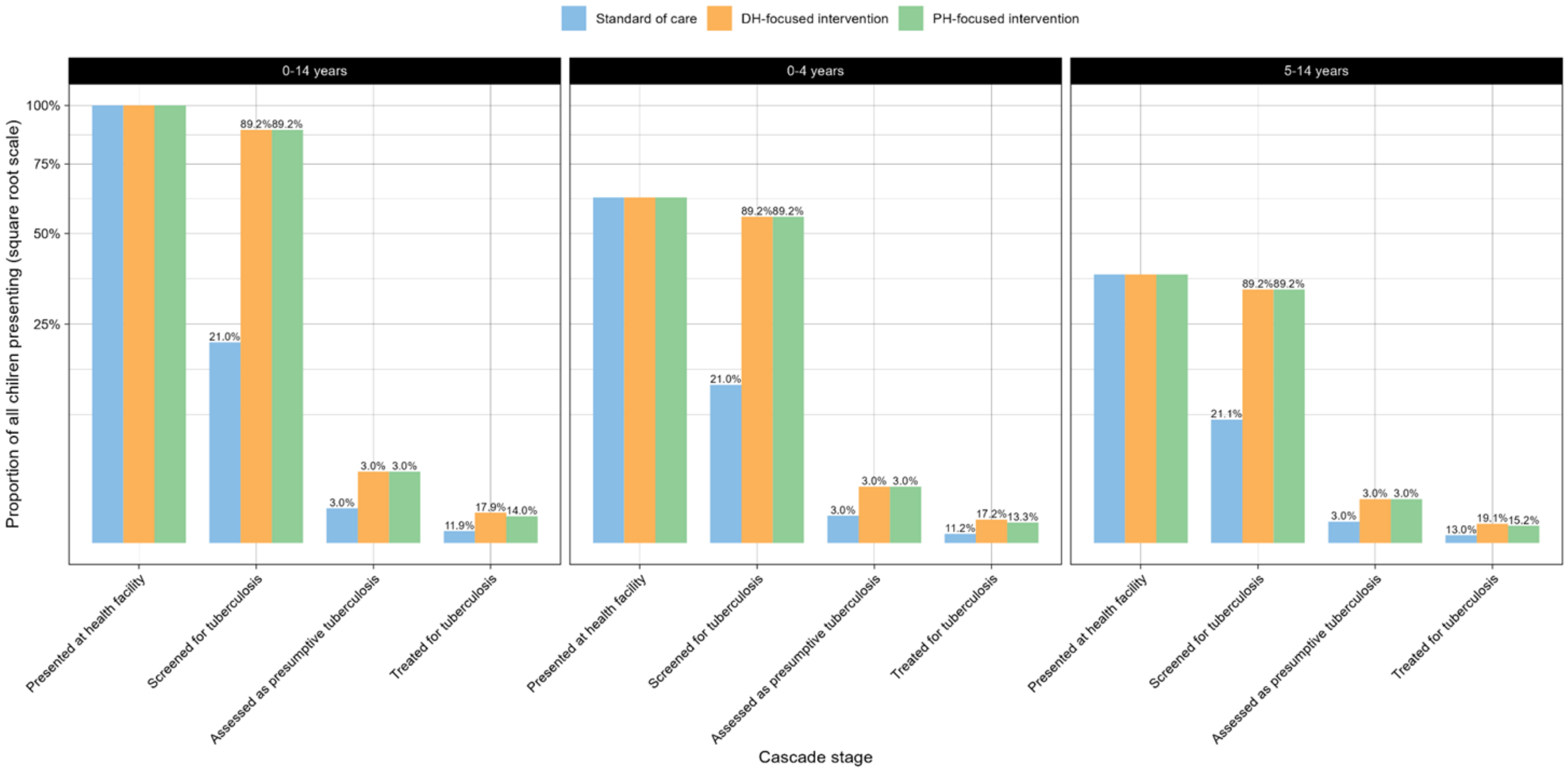


**Figure 16**. Screening to diagnosis cascades during the intervention period by decentralisation approach and age group. Percentages are bar height as a proportion of the corresponding bar at the previous stage.

Supplementary results - Other modelled health outcomes and cost per health outcome

**Table 11.** Undiscounted DALYs and ICERs

| Arm | Type of outcome | Per 100,000 OPD initial visits | Cambodia | Cameroon | Côte d'Ivoire | Mozambique | Sierra Leone | Uganda |
|---|---|---|---|---|---|---|---|---|
| Standard of care | Full | Cost (2021 US$) | 10,449 (1,164 to 32,054) | 13,430 (1,453 to 41,195) | 15,574 (1,797 to 46,247) | 10,359 (1,073 to 33,665) | 10,164 (1,100 to 32,066) | 13,581 (1,575 to 40,263) |
| | | DALY | 3,053 (440 to 9,108) | 2,752 (397 to 8,209) | 2,662 (384 to 7,942) | 2,868 (414 to 8,558) | 2,618 (377 to 7,810) | 2,906 (419 to 8,670) |
| DH-focused | Full | Cost (2021 US$) | 168,490 (48,731 to 375,050) | 198,455 (57,910 to 435,159) | 205,544 (64,514 to 440,572) | 181,355 (49,499 to 409,446) | 176,682 (46,093 to 398,673) | 193,742 (59,315 to 415,612) |
| | | DALY | 1,570 (212 to 5,222) | 1,415 (191 to 4,704) | 1,369 (185 to 4,556) | 1,475 (199 to 4,907) | 1,346 (182 to 4,475) | 1,495 (202 to 4,970) |
| | Incremental to SOC | DALY averted | 1,483 (148 to 4,705) | 1,337 (133 to 4,244) | 1,293 (129 to 4,097) | 1,393 (139 to 4,418) | 1,272 (127 to 4,038) | 1,412 (141 to 4,480) |
| | | ICER ($/DALY) | 107 | 138 | 147 | 123 | 131 | 128 |
| PHC-focused | Full | Cost (2021 US$) | 257,022 (71,606 to 583,728) | 295,105 (84,794 to 661,929) | 303,124 (92,655 to 677,697) | 276,615 (73,237 to 639,476) | 273,622 (72,754 to 628,440) | 286,493 (82,536 to 644,522) |
| | | DALY | 1,774 (250 to 5,488) | 1,599 (226 to 4,945) | 1,547 (218 to 4,778) | 1,667 (235 to 5,154) | 1,521 (215 to 4,704) | 1,689 (238 to 5,223) |

| | | | | | | | | |
|---|---|---|---|---|---|---|---|---|
| | Incremental to SOC | DALY averted | 1,279 (129 to 3,958) | 1,153 (116 to 3,569) | 1,115 (112 to 3,449) | 1,202 (121 to 3,717) | 1,097 (110 to 3,396) | 1,218 (123 to 3,767) |
| | | ICER ($/DALY) | 193 | 244 | 258 | 222 | 240 | 224 |

SOC: standard of care, TB: tuberculosis, OPD: patients presenting at outpatient department, DH: district hospital, PHC: primary health centre, DALY: disability-adjusted life year, ICER: incremental cost-effectiveness ratio

### Supplementary results - Cost per child treated by cost input and by care stage

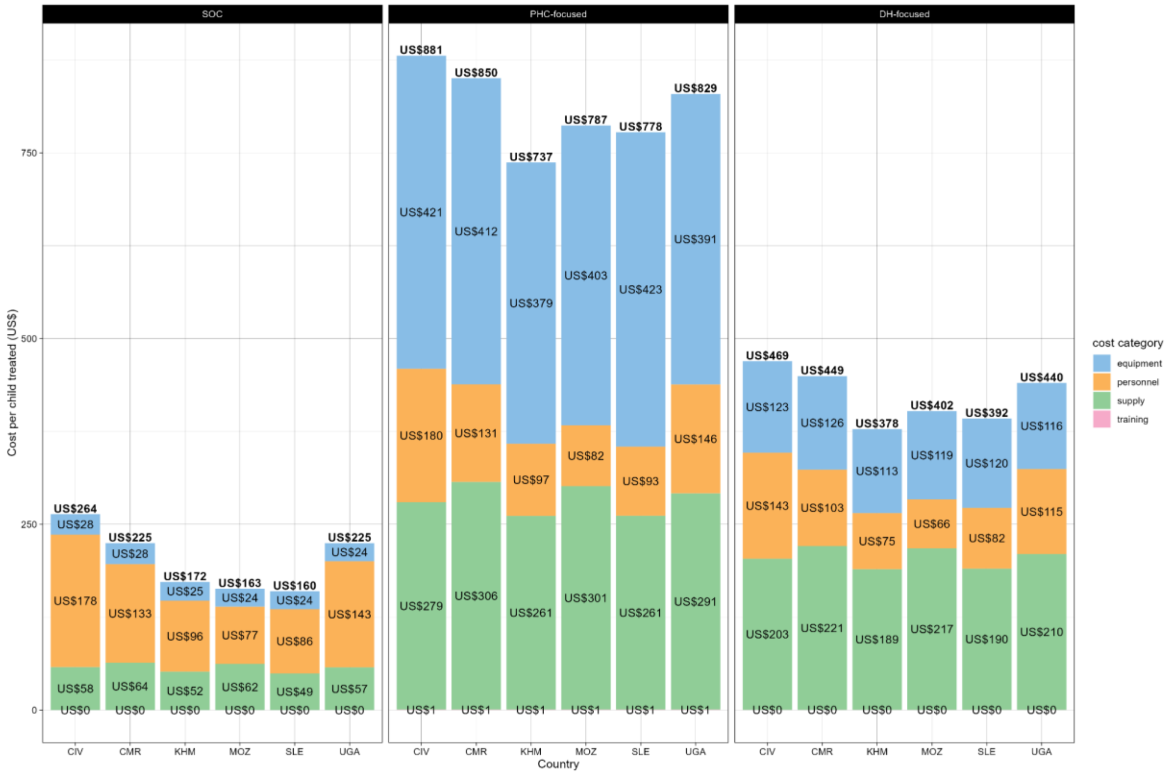


**Figure 17**. Cost per child treated by cost input and by strategy - absolute values

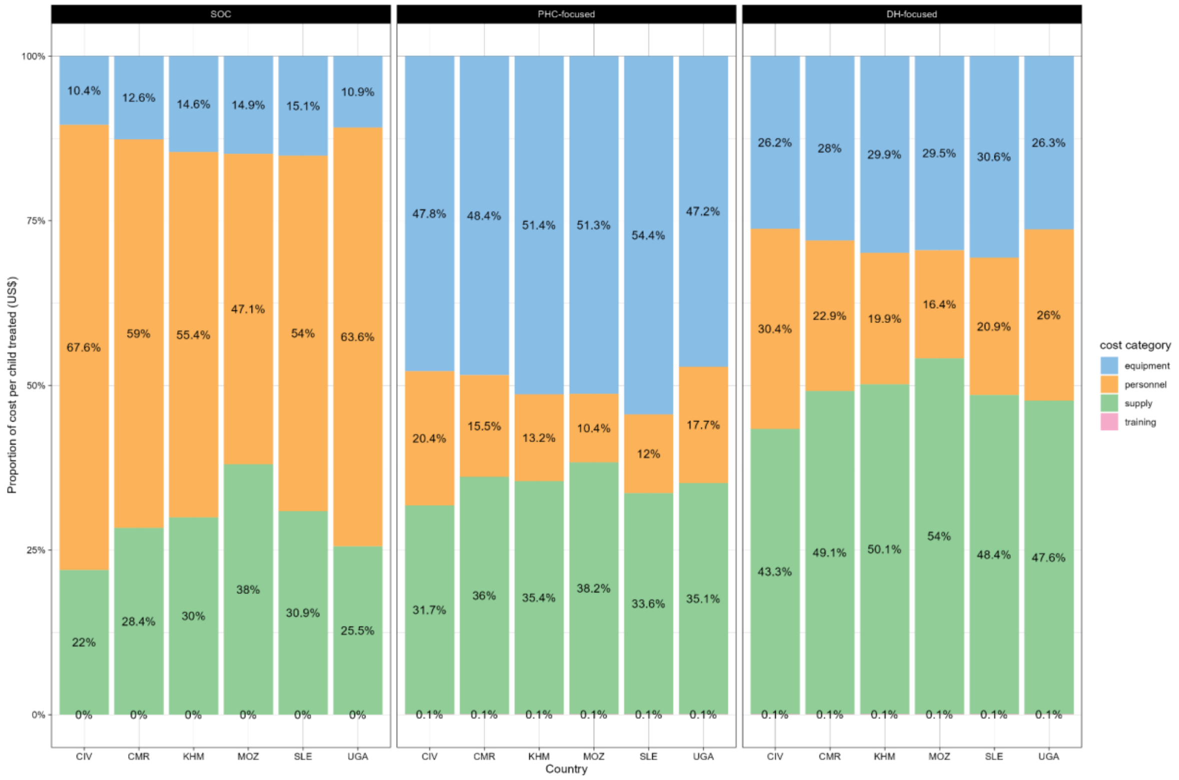


**Figure 18**. Cost per child treated by cost input and by strategy - percentage values

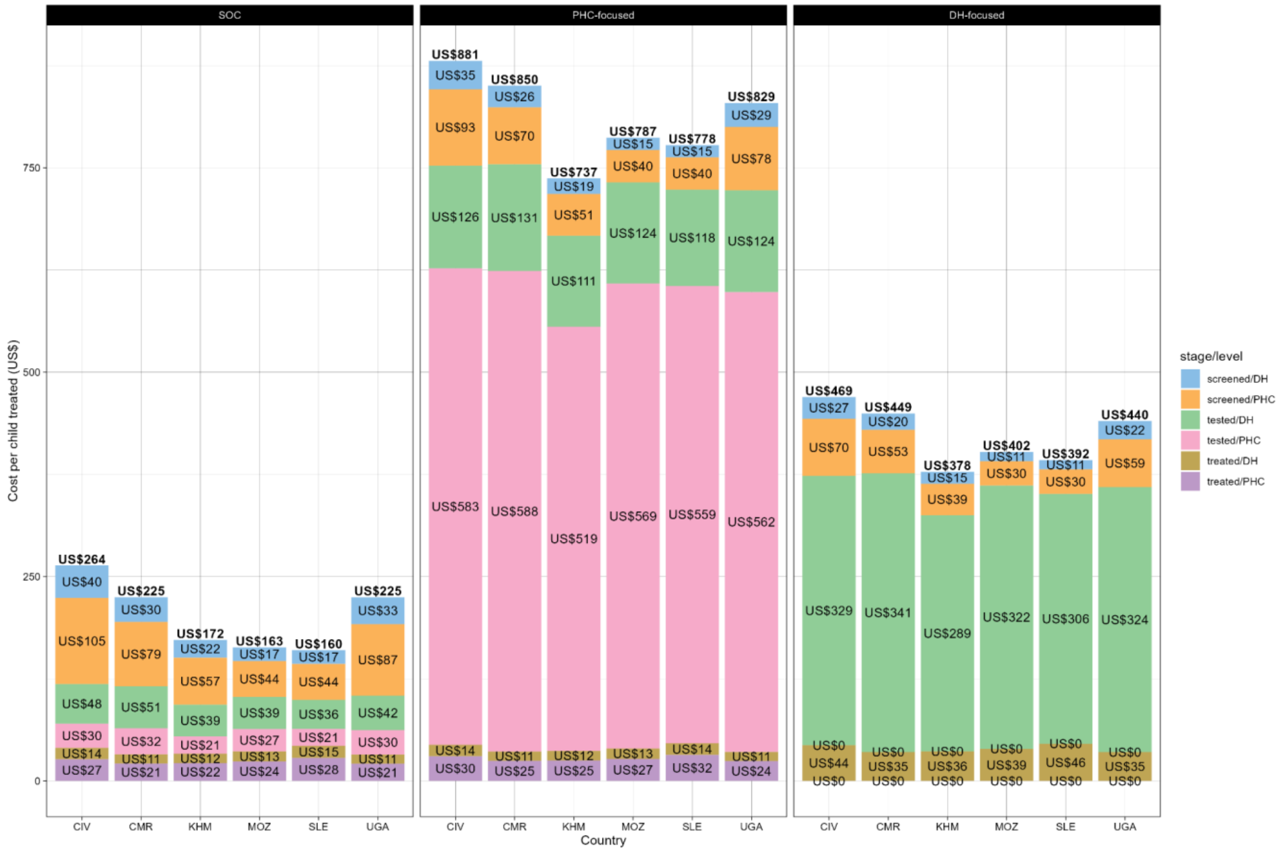


**Figure 19**. Cost per child treated by care stage and by strategy - absolute values

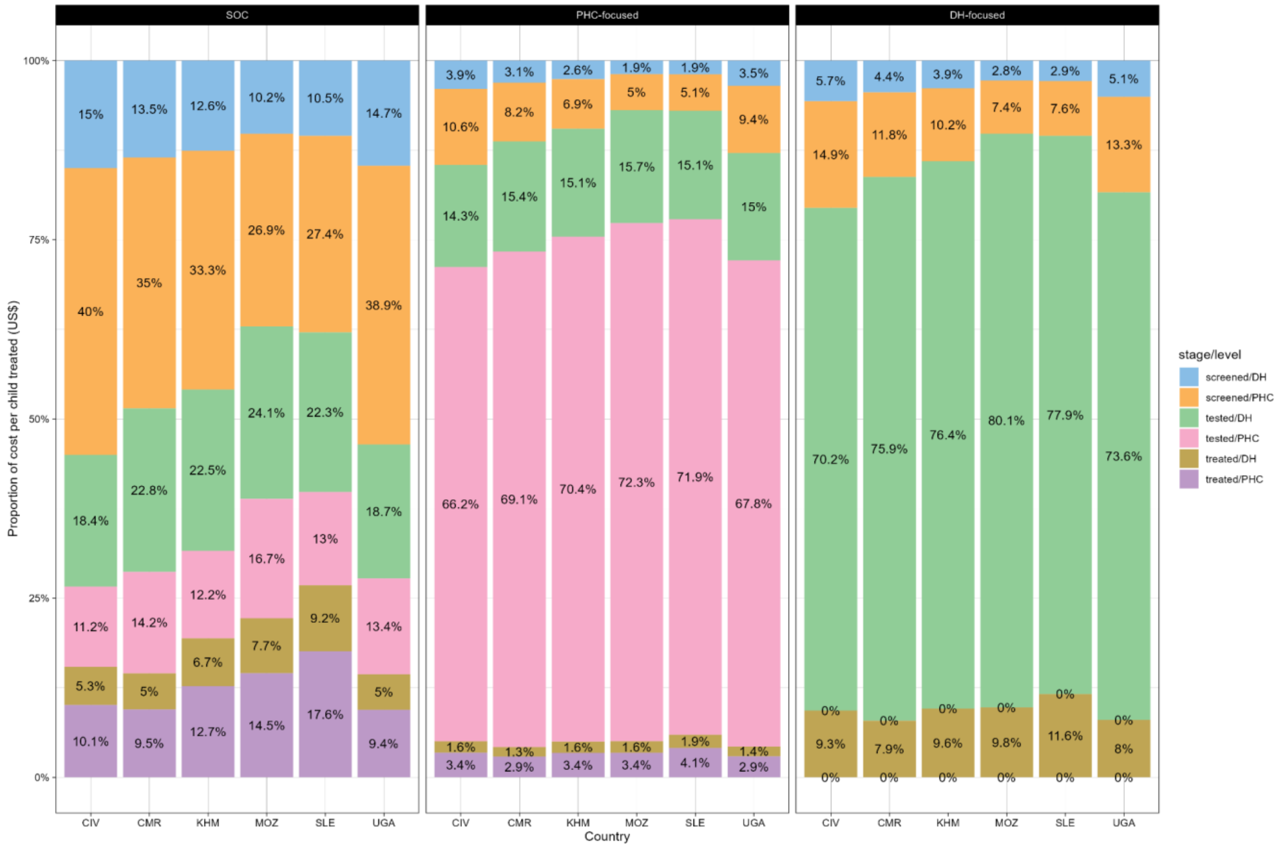


**Figure 20**. Cost per child treated by care stage and by strategy - percentage values

#### Supplementary results - Sensitivity analysis

**Table 12.** Sensitivity analysis of the effect of paediatric tuberculosis prevalence and discount rate applied on life years on the estimated incremental cost-effectiveness ratios

| | | | Incremental cost-effectiveness ratios (2021 US$/DALY averted) | | | | | |
|---|---|---|---|---|---|---|---|---|
| Comparison | Tuberculosis prevalence | Discount rate | Cambodia | Cameroon | Côte d'Ivoire | Mozambique | Sierra Leone | Uganda |
| DH-focused strategy versus standard of care | 500/100,000 | 0% | 31 | 40 | 42 | 35 | 38 | 36 |
| | | 3% | 76 | 94 | 98 | 86 | 89 | 89 |
| | | 6% | 138 | 167 | 173 | 153 | 156 | 159 |
| | 200/100,000 | 0% | 69 | 89 | 95 | 79 | 85 | 82 |
| | | 3% | 171 | 213 | 222 | 192 | 198 | 200 |
| | | 6% | 309 | 377 | 390 | 343 | 348 | 359 |

| | | | | | | | | |
|---|---|---|---|---|---|---|---|---|
| | 50/100,000 | 0% | 259 | 338 | 358 | 298 | 318 | 311 |
| | | 3% | 644 | 806 | 839 | 723 | 745 | 759 |
| | | 6% | 1,164 | 1,427 | 1,473 | 1,290 | 1,307 | 1,358 |
| PHC-focused strategy versus standard of care | 500/100,000 | 0% | 55 | 70 | 73 | 63 | 69 | 64 |
| | | 3% | 137 | 166 | 172 | 154 | 162 | 156 |
| | | 6% | 247 | 294 | 302 | 274 | 283 | 278 |
| | 200/100,000 | 0% | 125 | 158 | 167 | 144 | 156 | 145 |
| | | 3% | 311 | 378 | 391 | 348 | 365 | 354 |
| | | 6% | 561 | 670 | 688 | 621 | 641 | 634 |
| | 50/100,000 | 0% | 474 | 602 | 635 | 545 | 591 | 553 |
| | | 3% | 1,179 | 1,438 | 1,488 | 1,320 | 1,385 | 1,347 |
| | | 6% | 2,130 | 2,546 | 2,614 | 2,357 | 2,429 | 2,410 |

DH: district hospital, PHC: primary health centre

### Budget impact analysis

#### Methodological approach

The budget impact of implementing each strategy was calculated as follows:

$$C_{ijk} = c1 \cdot s1_{ijk} + c2 \cdot s2_{ijk} + c3_i \cdot s3_{ijk} + c4 \cdot s4_{ijk} + c5_i \cdot s5_{ijk} + c6_{ij} \cdot s6_{ijk}$$

i: country (Cambodia, Cameroon, Côte d'Ivoire, Mozambique, Sierra Leone, Uganda),

j: arm (SOC, DH-focused, PHC-focused),

k: year of implementation (Year 1, Year 2, Year 3, Year 4, Year 5)

**Table 13**. Budget impact analysis - parameters

| # | Area of resources/costs | Cost parameters (c) | Scale parameters (s) | Data sources |
|---|---|---|---|---|
| 1 | Staff training | Cost per trained HCW | Number of HCW | TB-Speed primary costing, WHO Global Health Observatory |
| 2 | Equipment at DH | Cost of equipment items (including maintenance) at DH | Number of DH | TB-Speed primary costing, Ministries of Health |
| 3 | Equipment delivery to DH | Cost of equipment delivery at DH | Number of DH | TB-Speed primary costing, Ministries of Health |
| 4 | Equipment at PHC | Cost of equipment items (including maintenance) at PHC | Number of PHC | TB-Speed primary costing, Ministries of Health |
| 5 | Equipment delivery to PHC | Cost of equipment delivery at PHC | Number of PHC | TB-Speed primary costing, |

| | | | | |
|---|---|---|---|---|
| | | | | Ministries of Health |
| 6 | TB assessment | Cost per TB case treated (including personnel and supplies costs, excluding training, equipment & maintenance, and delivery chain costs) | Annual rate of TB notification in children 0-14 years | TB-Speed CE modelling, WHO Global Tuberculosis Programme |

We accounted for the additional costs of implementing the intervention compared to current health spending for the standard of care. National projections were based on the number of healthcare workers involved in tuberculosis care, the number and type of existing health facilities, and reported tuberculosis notifications. Budget impact was calculated over an horizon of 5 years. Costs were not discounted and are presented in US dollars. The approach was conservative and ignored existing tuberculosis screening/assessment activities.

Across countries, a TB-Speed baseline assessment survey showed that 69% of DHs had a functioning CXR, and that GeneXpert machine availability was at 46% and 1% for DH and PHC sites, respectively (sample sizes per country were too small to produce representative country-specific estimates). Almost all DHs were equipped with analogue CXR, so we budgeted for a digital detector and computing equipment in all DH. We assumed that all health facilities would need new NPA sample collection equipment. To allocate national HCW numbers to DH and PHC levels, we assume that each PHC has at most 1 doctor, 1 nurse, and 1 lab technician, therefore the share of HCW at DH level is the national total HCW - PHC-based HCW × 3. This suggests that tertiary hospital staff are retrained, but as they compose a minor share of the total HCWs, we expected it would not affect our estimates. Data to inform scale-up, such as the number of DHs and PHCs were collected during the study or found in the literature.[18]

The impact of the intervention was assessed using the number of child tuberculosis cases treated. For the SOC, we used the annual rate of tuberculosis notification in children 0-14 years old from the WHO Global TB Programme database (year 2019 to avoid distortions related to the COVID-19 pandemic), and it was assumed to be constant over the 5-year period. Before and during the COVID-19 pandemic (period 2018-2022), we did not observe a noticeable drop in tuberculosis case notifications. For the DH-focused strategy, we used the expected annual rate of tuberculosis notification in children 0-14 years old, based on the TB-Speed data on the proportion of children treated for tuberculosis among those presenting at OPD at DHs in DH-focused districts. We used the data on 0-14 year old OPD volumes and the number of tuberculosis cases treated from the TB-Speed observation phase (pre-intervention), and WHO tuberculosis notification rates, to retrieve national OPD visits among 0-14 year olds (also assumed constant over time). To account for a phased intervention scale-up, we assumed a 3-year period for rollout of the equipment (CXR machines, Xpert, etc) to all DHs and/or PHCs, and training of all staff. Consequently, we assumed a progressive increase of impact (Y1:20%; Y2:40%; Y3:60%; Y4:80%; Y5:100%) for the two decentralisation strategies.

We conducted sensitivity analyses on the following sources of uncertainty. The main cost drivers were equipment, so most of the assumptions tested are related to this cost input. Each analysis compares our model assumptions 'base' case versus alternative scenarios.

*1.a. Accounting for potential economies of scale on delivery chain costs (base: no economies of scale; economies of scale)*

We evaluated potential economies of scale on the intervention in collaboration with experts at MSF-logistique. At scale-up, the cost of purchasing each item of equipment (for example, CXR or GeneXpert machines) would remain the same. Field-based personnel costs would be proportional with scale. Supplies (for example, stool containers or gloves) and training costs only contributed to a small proportion of total costs, so were not explored as potential sources of economies of scale. The only exception was the GeneXpert cartridge kit price (~$10/per kit), which was assumed to remain constant at scale-up (as for equipment items) - and tested in *1.c.* Therefore, the only potential sources for economies of scale were equipment international delivery chain costs. For each strategy, economies of scale were estimated by our partner MSF-logistique based on the total number of equipment items that would be purchased for a national scale-up of the interventions. The experts used a country-specific average cost per air kilogram (as of 30th December, 2022), and did not assess maritime costs because they are highly variable.

*1.b. MSF-logistique market screening - reduced mucus aspirator cost (base: $1,850; $440)*

Currently NPA, which requires mucus aspirators, is seldom used at district level, compared to higher health facility level. For its decentralised use at DH and PHC level, simple and robust equipment or alternative methods are needed. Several battery-operated suction machines are available on the market. MSF-Logistique conducted a market screening for mucus aspirators. They found two eligible and cheaper machines. The cost of the cheapest machine was assessed in this analysis.

*1.c. Negotiated price of cartridge kit (base: $10; $5)*

The literature raises the issue of Xpert test cartridges for tuberculosis having a high cost. There is an ongoing debate on negotiated prices for humanitarian organisations, decided on a case-by-case basis by the manufacturer Cepheid in consultation with local and global stakeholders. We assessed here the impact of a reduction to a unit price of US$5.

*1.d. Change in % of DH sites equipped with CXR and Xpert machines (base: 69% CXR & 46% GeneXpert; 35% & 20%)*

Related to the first point, reliable data on health facilities equipped with 'functioning' or usable machines can be challenging to obtain. Such data was collected on TB-Speed sites but may not be representative of each country. To account for this uncertainty, we halved the % of sites equipped in the main analysis. This source of uncertainty is of interest because, in real life, sites already equipped with analogue CXR might continue using them, and new digital CXR equipment might be purchased for facilities without CXR.

*1.e. Change in equipment installation, repair and/or maintenance costs (base: 15%; 10%; 20%)*

To our knowledge, there is no data in the literature on the total installation, repair and/or maintenance costs over the useful life of equipment such as Xpert and CXR machines. We assumed an additional 15% of purchase price cost to account for this critical expense, too often overlooked, in budget impact analysis. In the sensitivity analysis, we evaluate a 5% variation to estimate its impact on total costs. We did not account for repair and/or maintenance costs of other equipment, because it was either not expensive or not subject to such service.

*1.f. Best and worst case scenario*

Finally, we combined the above assumptions into best and worst case scenarios to evaluate the overall impact of all assumptions on total scale-up costs.

#### Supplementary results - Scale-up costs by intervention, year, cost input, and country

We present below the results from the BIA disaggregated by country, intervention, implementation versus routine costs, and by cost inputs (**Table 14**).

**Table 14.** Scale-up costs by intervention, year, cost input, and country (2021 US$)

| | | Cambodia | | | | | | |
|---|---|---|---|---|---|---|---|---|
| | | Implementation costs | | Running costs | | Total | Additional costs compared to SOC | % |
| Intervention | Expenditure | Training | Equipment | Personnel | Supply | | | |
| Standard of care | 2022 | | | 176,817 | 95,557 | 272,373 | | |
| | 2023 | | | 176,817 | 95,557 | 272,373 | | |
| | 2024 | | | 176,817 | 95,557 | 272,373 | | |
| | 2025 | | | 176,817 | 95,557 | 272,373 | | |
| | 2026 | | | 176,817 | 95,557 | 272,373 | | |
| | Total | | | 884,083 | 477,783 | 1,361,867 | | |
| DH-focused | 2022 | 137,747 | 2,167,135 | 229,709 | 576,865 | 3,111,456 | 2,839,082 | 1042% |
| | 2023 | 137,747 | 2,167,135 | 320,031 | 803,690 | 3,428,603 | 3,156,229 | 1159% |

| | | | | | | | | |
|---|---|---|---|---|---|---|---|---|
| | 2024 | 137,747 | 2,167,135 | 410,353 | 1,030,515 | 3,745,750 | 3,473,376 | 1275% |
| | 2025 | 0 | 0 | 500,675 | 1,257,340 | 1,758,015 | 1,485,641 | 545% |
| | 2026 | 0 | 0 | 590,997 | 1,484,165 | 2,075,162 | 1,802,788 | 662% |
| | Total | 413,242 | 6,501,404 | 2,051,763 | 5,152,575 | 14,118,984 | 12,757,118 | 937% |
| | % | 3% | 46% | 15% | 36% | 100% | | |
| PHC-focused | 2022 | 182,803 | 9,354,287 | 193,969 | 520,744 | 10,251,803 | 9,979,430 | 3664% |
| | 2023 | 182,803 | 9,354,287 | 208,355 | 559,366 | 10,304,811 | 10,032,438 | 3683% |
| | 2024 | 182,803 | 9,354,287 | 222,741 | 597,987 | 10,357,818 | 10,085,445 | 3703% |
| | 2025 | 0 | 0 | 237,127 | 636,609 | 873,736 | 601,362 | 221% |
| | 2026 | 0 | 0 | 251,513 | 675,230 | 926,743 | 654,370 | 240% |
| | Total | 548,409 | 28,062,861 | 1,113,704 | 2,989,937 | 32,714,911 | 31,353,045 | 2302% |
| | % | 2% | 86% | 3% | 9% | 100% | | |

| | | Côte d'Ivoire | | | | | | |
|---|---|---|---|---|---|---|---|---|
| | | Implementation costs | | Running costs | | Total | Additional costs compared to SOC | % |
| Intervention | Expenditure | Training | Equipment | Personnel | Supply | | | |
| Standard of care | 2022 | | | 181,302 | 58,950 | 240,252 | | |
| | 2023 | | | 181,302 | 58,950 | 240,252 | | |
| | 2024 | | | 181,302 | 58,950 | 240,252 | | |
| | 2025 | | | 181,302 | 58,950 | 240,252 | | |
| | 2026 | | | 181,302 | 58,950 | 240,252 | | |
| | Total | | | 906,510 | 294,750 | 1,201,260 | | |
| DH-focused | 2022 | 211,366 | 4,978,121 | 239,403 | 341,068 | 5,769,959 | 5,529,707 | 2302% |
| | 2023 | 211,366 | 4,978,121 | 333,537 | 475,177 | 5,998,202 | 5,757,950 | 2397% |
| | 2024 | 211,366 | 4,978,121 | 427,671 | 609,286 | 6,226,444 | 5,986,192 | 2492% |

| | | | | | | | | |
|---|---|---|---|---|---|---|---|---|
| | 2025 | 0 | 0 | 521,805 | 743,395 | 1,265,200 | 1,024,948 | 427% |
| | 2026 | 0 | 0 | 615,939 | 877,503 | 1,493,443 | 1,253,191 | 522% |
| | Total | 634,098 | 14,934,364 | 2,138,356 | 3,046,429 | 20,753,247 | 19,551,987 | 1628% |
| | % | 3% | 72% | 10% | 15% | 100% | | |
| PHC-focused | 2022 | 273,362 | 29,379,640 | 197,425 | 306,996 | 30,157,424 | 29,917,172 | 12452% |
| | 2023 | 273,362 | 29,379,640 | 212,068 | 329,764 | 30,194,834 | 29,954,583 | 12468% |
| | 2024 | 273,362 | 29,379,640 | 226,710 | 352,533 | 30,232,245 | 29,991,993 | 12484% |
| | 2025 | 0 | 0 | 241,352 | 375,301 | 616,654 | 376,402 | 157% |
| | 2026 | 0 | 0 | 255,994 | 398,070 | 654,065 | 413,813 | 172% |
| | Total | 820,086 | 88,138,921 | 1,133,550 | 1,762,664 | 91,855,222 | 90,653,962 | 7547% |
| | % | 1% | 96% | 1% | 2% | 100% | | |
| | | Sierra Leone | | | | | | |

| | | Implementation costs | | Running costs | | Total | Additional costs compared to SOC | % |
|---|---|---|---|---|---|---|---|---|
| Intervention | Expenditure | Training | Equipment | Personnel | Supply | | | |
| Standard of care | 2022 | | | 202,789 | 116,157 | 318,946 | | |
| | 2023 | | | 202,789 | 116,157 | 318,946 | | |
| | 2024 | | | 202,789 | 116,157 | 318,946 | | |
| | 2025 | | | 202,789 | 116,157 | 318,946 | | |
| | 2026 | | | 202,789 | 116,157 | 318,946 | | |
| | Total | | | 1,013,945 | 580,783 | 1,594,728 | | |
| DH-focused | 2022 | 80,806 | 1,606,766 | 317,081 | 735,917 | 2,740,570 | 2,421,624 | 759% |
| | 2023 | 80,806 | 1,606,766 | 441,758 | 1,025,282 | 3,154,612 | 2,835,666 | 889% |
| | 2024 | 80,806 | 1,606,766 | 566,435 | 1,314,646 | 3,568,653 | 3,249,708 | 1019% |
| | 2025 | 0 | 0 | 691,113 | 1,604,011 | 2,295,124 | 1,976,178 | 620% |

| | | | | | | | | |
|---|---|---|---|---|---|---|---|---|
| | 2026 | 0 | 0 | 815,790 | 1,893,376 | 2,709,165 | 2,390,220 | 749% |
| | Total | 242,417 | 4,820,298 | 2,832,177 | 6,573,232 | 14,468,123 | 12,873,395 | 807% |
| | % | 2% | 33% | 20% | 45% | 100% | | |
| PHC-focused | 2022 | 89,217 | 3,351,833 | 235,901 | 662,389 | 4,339,339 | 4,020,393 | 1261% |
| | 2023 | 89,217 | 3,351,833 | 253,397 | 711,515 | 4,405,961 | 4,087,016 | 1281% |
| | 2024 | 89,217 | 3,351,833 | 270,892 | 760,642 | 4,472,584 | 4,153,638 | 1302% |
| | 2025 | 0 | 0 | 288,388 | 809,769 | 1,098,157 | 779,211 | 244% |
| | 2026 | 0 | 0 | 305,884 | 858,895 | 1,164,779 | 845,834 | 265% |
| | Total | 267,650 | 10,055,499 | 1,354,462 | 3,803,210 | 15,480,820 | 13,886,092 | 871% |
| | % | 2% | 65% | 9% | 25% | 100% | | |
| | | Cameroon | | | | | | |
| | | Implementation costs | | Running costs | | Total | Additional | % |

| Intervention | Expenditure | Training | Equipment | Personnel | Supply | | costs compared to SOC | |
|---|---|---|---|---|---|---|---|---|
| Standard of care | 2022 | | | 166,513 | 80,030 | 246,544 | | |
| | 2023 | | | 166,513 | 80,030 | 246,544 | | |
| | 2024 | | | 166,513 | 80,030 | 246,544 | | |
| | 2025 | | | 166,513 | 80,030 | 246,544 | | |
| | 2026 | | | 166,513 | 80,030 | 246,544 | | |
| | Total | | | 832,567 | 400,152 | 1,232,719 | | |
| DH-focused | 2022 | 114,068 | 5,534,774 | 212,428 | 456,049 | 6,317,318 | 6,070,775 | 2462% |
| | 2023 | 114,068 | 5,534,774 | 295,956 | 635,368 | 6,580,165 | 6,333,621 | 2569% |
| | 2024 | 114,068 | 5,534,774 | 379,483 | 814,688 | 6,843,012 | 6,596,468 | 2676% |
| | 2025 | 0 | 0 | 463,011 | 994,007 | 1,457,018 | 1,210,474 | 491% |
| | 2026 | 0 | 0 | 546,538 | 1,173,327 | 1,719,865 | 1,473,321 | 598% |

| | | | | | | | | |
|---|---|---|---|---|---|---|---|---|
| | Total | 342,203 | 16,604,321 | 1,897,416 | 4,073,439 | 22,917,378 | 21,684,659 | 1759% |
| | % | 1% | 72% | 8% | 18% | 100% | | |
| PHC-focused | 2022 | 201,494 | 20,863,303 | 178,138 | 415,307 | 21,658,242 | 21,411,698 | 8685% |
| | 2023 | 201,494 | 20,863,303 | 191,349 | 446,109 | 21,702,255 | 21,455,711 | 8703% |
| | 2024 | 201,494 | 20,863,303 | 204,561 | 476,910 | 21,746,269 | 21,499,725 | 8720% |
| | 2025 | 0 | 0 | 217,773 | 507,712 | 725,485 | 478,941 | 194% |
| | 2026 | 0 | 0 | 230,985 | 538,514 | 769,498 | 522,955 | 212% |
| | Total | 604,482 | 62,589,909 | 1,022,806 | 2,384,552 | 66,601,749 | 65,369,030 | 5303% |
| | % | 1% | 94% | 2% | 4% | 100% | | |

| | | Mozambique | | | | | | |
|---|---|---|---|---|---|---|---|---|
| | | Implementation costs | | Running costs | | Total | Additional costs compared to SOC | % |
| Intervention | Expenditure | Training | Equipment | Personnel | Supply | | | |

| | | | | | | | | |
|---|---|---|---|---|---|---|---|---|
| Standard of care | 2022 | | | 990,514 | 798,750 | 1,789,264 | | |
| | 2023 | | | 990,514 | 798,750 | 1,789,264 | | |
| | 2024 | | | 990,514 | 798,750 | 1,789,264 | | |
| | 2025 | | | 990,514 | 798,750 | 1,789,264 | | |
| | 2026 | | | 990,514 | 798,750 | 1,789,264 | | |
| | Total | | | 4,952,570 | 3,993,748 | 8,946,319 | | |
| DH-focused | 2022 | 166,350 | 1,727,834 | 1,399,862 | 4,603,498 | 7,897,544 | 6,108,280 | 341% |
| | 2023 | 166,350 | 1,727,834 | 1,950,291 | 6,413,606 | 10,258,081 | 8,468,817 | 473% |
| | 2024 | 166,350 | 1,727,834 | 2,500,721 | 8,223,713 | 12,618,618 | 10,829,354 | 605% |
| | 2025 | 0 | 0 | 3,051,150 | 10,033,821 | 13,084,971 | 11,295,707 | 631% |
| | 2026 | 0 | 0 | 3,601,579 | 11,843,928 | 15,445,508 | 13,656,244 | 763% |
| | Total | 499,049 | 5,183,503 | 12,503,603 | 41,118,566 | 59,304,720 | 50,358,402 | 563% |

| | | | | | | | | |
|---|---|---|---|---|---|---|---|---|
| | % | 1% | 9% | 21% | 69% | 100% | | |
| PHC-focused | 2022 | 215,947 | 12,362,337 | 1,135,269 | 4,178,131 | 17,891,683 | 16,102,420 | 900% |
| | 2023 | 215,947 | 12,362,337 | 1,219,467 | 4,488,006 | 18,285,757 | 16,496,493 | 922% |
| | 2024 | 215,947 | 12,362,337 | 1,303,665 | 4,797,881 | 18,679,830 | 16,890,566 | 944% |
| | 2025 | 0 | 0 | 1,387,864 | 5,107,756 | 6,495,620 | 4,706,356 | 263% |
| | 2026 | 0 | 0 | 1,472,062 | 5,417,631 | 6,889,693 | 5,100,430 | 285% |
| | Total | 647,840 | 37,087,011 | 6,518,327 | 23,989,406 | 68,242,584 | 59,296,265 | 663% |
| | % | 1% | 54% | 10% | 35% | 100% | | |
| | | Uganda | | | | | | |
| | | Implementation costs | | Running costs | | Total | Additional costs compared to SOC | % |
| Intervention | Expenditure | Training | Equipment | Personnel | Supply | | | |
| Standard of | 2022 | | | 1,174,511 | 471,955 | 1,646,467 | | |

| | | | | | | | | |
|---|---|---|---|---|---|---|---|---|
| care | 2023 | | | 1,174,511 | 471,955 | 1,646,467 | | |
| | 2024 | | | 1,174,511 | 471,955 | 1,646,467 | | |
| | 2025 | | | 1,174,511 | 471,955 | 1,646,467 | | |
| | 2026 | | | 1,174,511 | 471,955 | 1,646,467 | | |
| | Total | | | 5,872,557 | 2,359,777 | 8,232,335 | | |
| DH-focused | 2022 | 743,572 | 5,254,484 | 1,550,974 | 2,837,768 | 10,386,798 | 8,740,331 | 531% |
| | 2023 | 743,572 | 5,254,484 | 2,160,821 | 3,953,586 | 12,112,463 | 10,465,996 | 636% |
| | 2024 | 743,572 | 5,254,484 | 2,770,667 | 5,069,404 | 13,838,128 | 12,191,661 | 740% |
| | 2025 | 0 | 0 | 3,380,514 | 6,185,223 | 9,565,737 | 7,919,270 | 481% |
| | 2026 | 0 | 0 | 3,990,361 | 7,301,041 | 11,291,402 | 9,644,935 | 586% |
| | Total | 2,230,715 | 15,763,453 | 13,853,337 | 25,347,022 | 57,194,528 | 48,962,193 | 595% |
| | % | 4% | 28% | 24% | 44% | 100% | | |

| | | | | | | | | |
|---|---|---|---|---|---|---|---|---|
| PHC-focused | 2022 | 853,704 | 39,316,515 | 1,300,367 | 2,583,327 | 44,053,913 | 42,407,446 | 2576% |
| | 2023 | 853,704 | 39,316,515 | 1,396,810 | 2,774,922 | 44,341,951 | 42,695,484 | 2593% |
| | 2024 | 853,704 | 39,316,515 | 1,493,253 | 2,966,517 | 44,629,989 | 42,983,522 | 2611% |
| | 2025 | 0 | 0 | 1,589,696 | 3,158,112 | 4,747,808 | 3,101,341 | 188% |
| | 2026 | 0 | 0 | 1,686,139 | 3,349,706 | 5,035,846 | 3,389,379 | 206% |
| | Total | 2,561,111 | 117,949,546 | 7,466,266 | 14,832,583 | 142,809,507 | 134,577,172 | 1635% |
| | % | 2% | 83% | 5% | 10% | 100% | | |

#### Supplementary results - Sensitivity analysis

We also report the findings from the sensitivity analysis by country and by intervention (**Figure 21**, **Figure 22**, **Figure 23**, **Figure 24**, **Figure 25**, **Figure 26**, **Figure 27**, **Figure 28**, **Figure 29**, **Figure 30**, **Figure 31**, **Figure 32**).

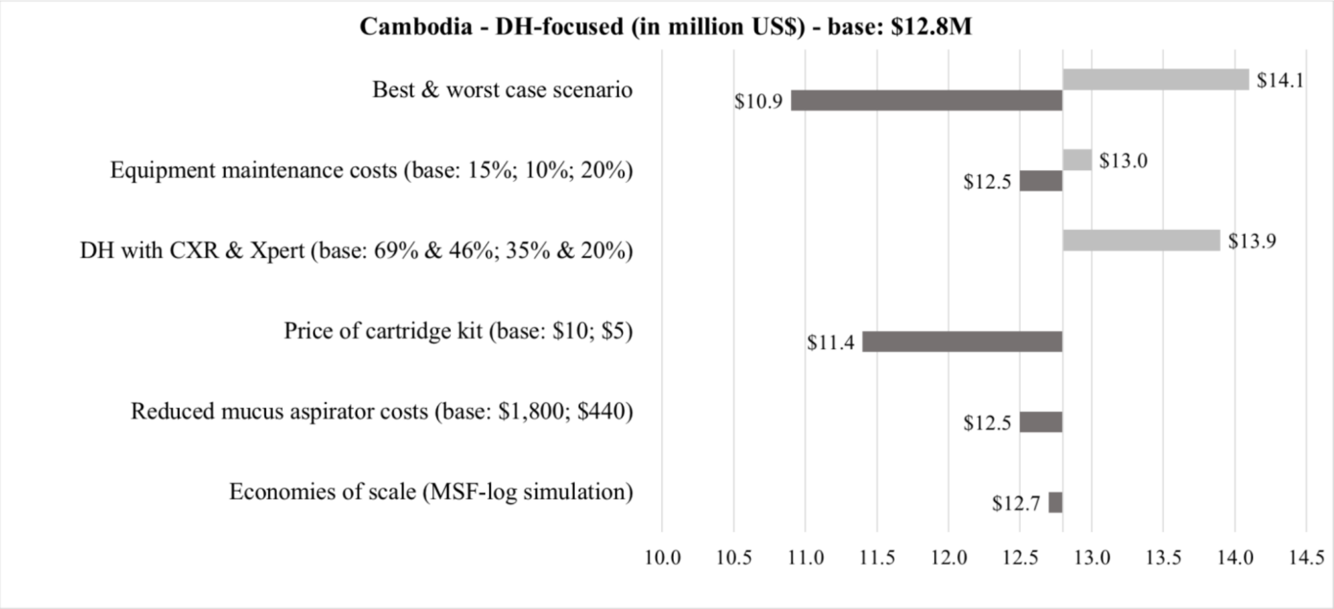


**Figure 21**. Tornado diagram on findings from sensitivity analysis on total scale-up costs of the DH-focused strategy in Cambodia

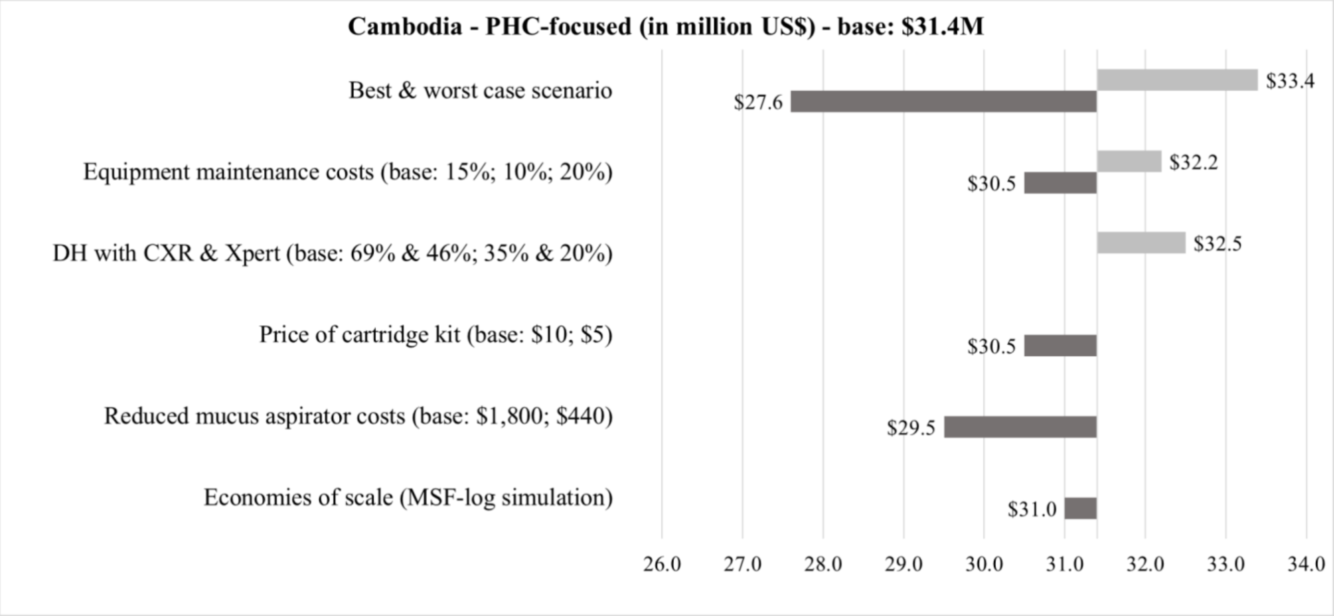


**Figure 22**. Tornado diagram on findings from sensitivity analysis on total scale-up costs of the PHC-focused strategy in Cambodia

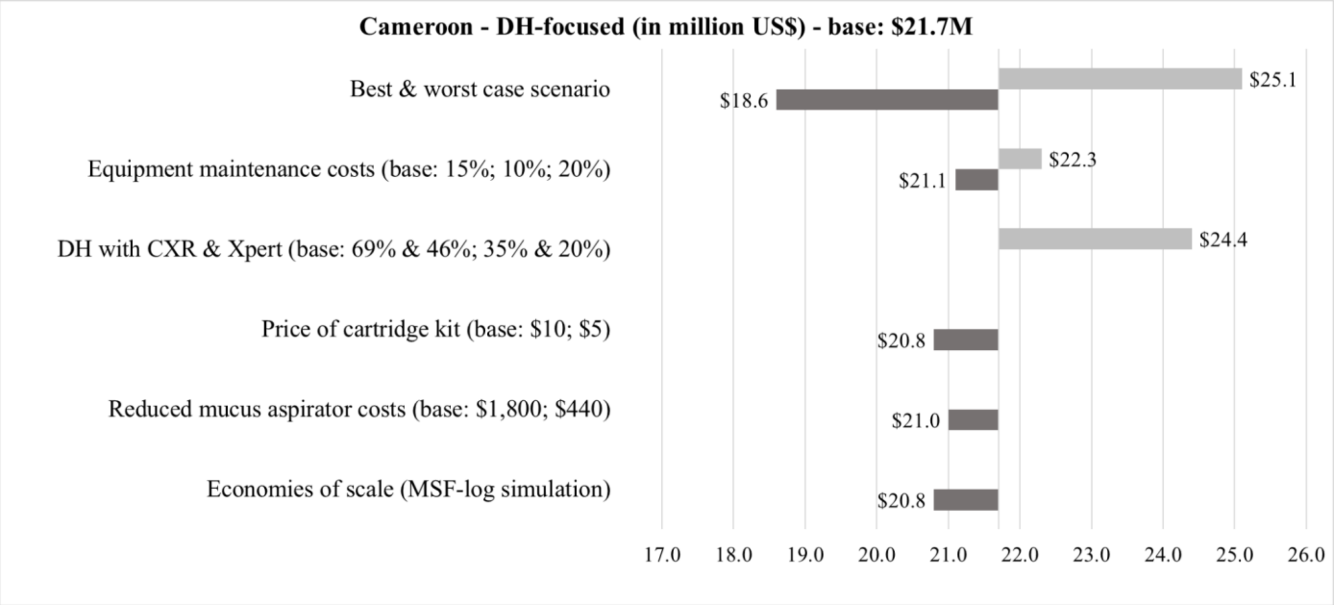


**Figure 23**. Tornado diagram on findings from sensitivity analysis on total scale-up costs of the DH-focused strategy in Cameroon

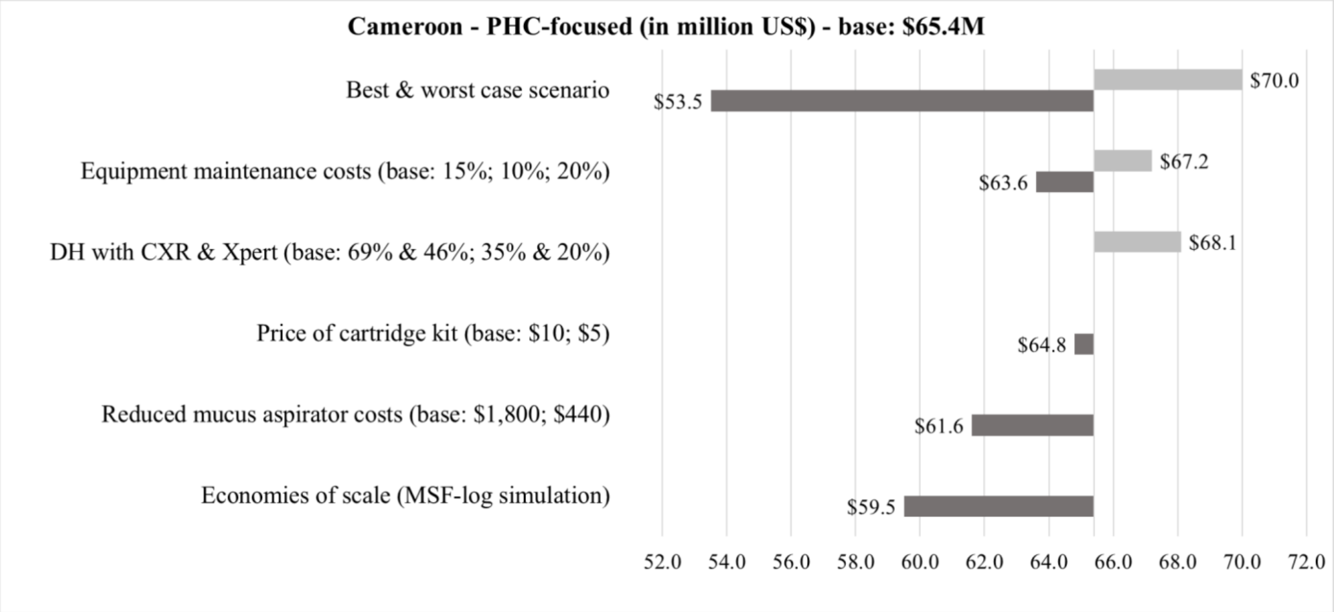


**Figure 24**. Tornado diagram on findings from sensitivity analysis on total scale-up costs of the PHC-focused strategy in Cameroon

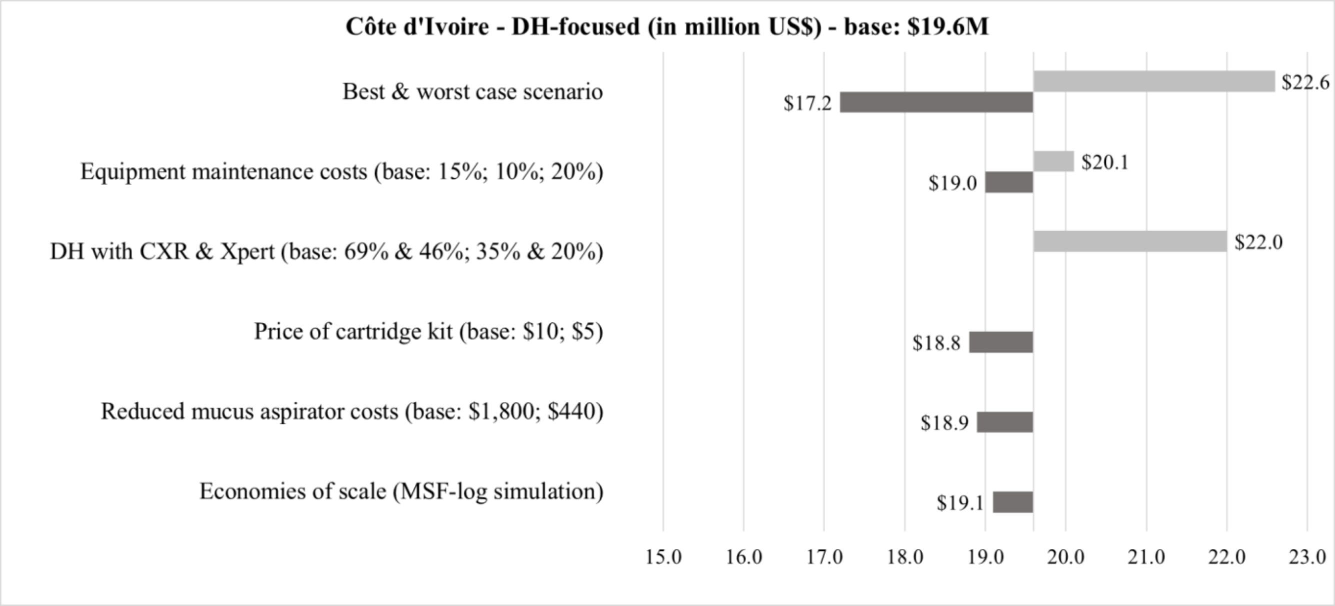


**Figure 25**. Tornado diagram on findings from sensitivity analysis on total scale-up costs of the DH-focused strategy in Côte d’Ivoire

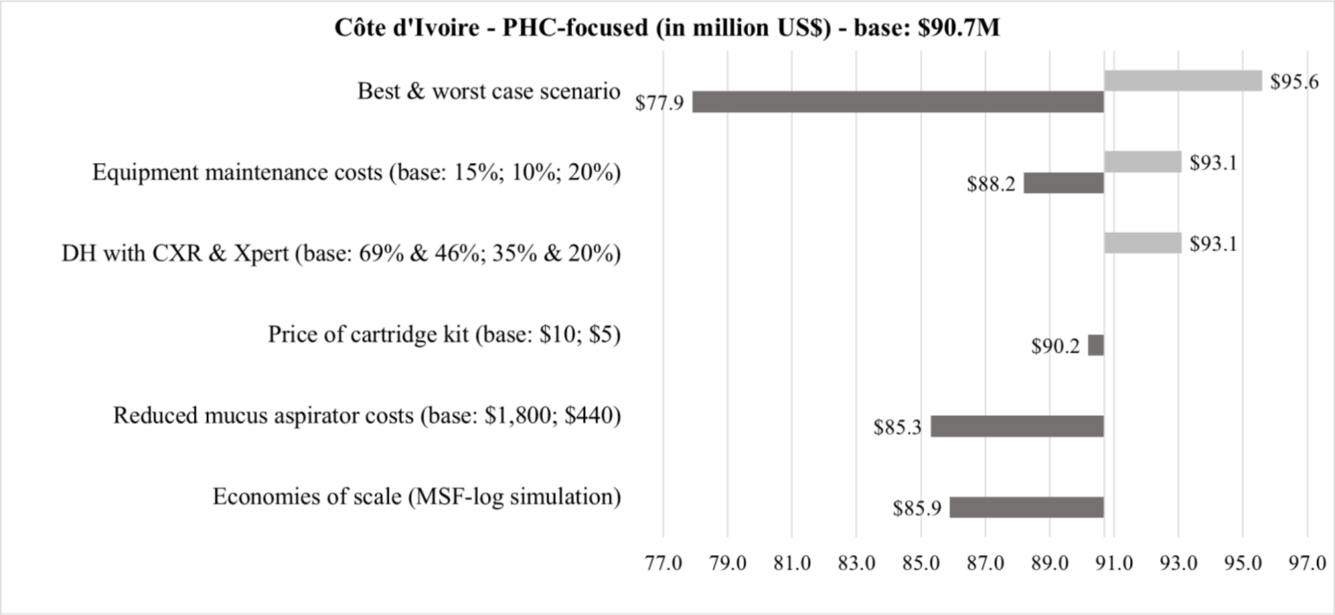


**Figure 26**. Tornado diagram on findings from sensitivity analysis on total scale-up costs of the PHC-focused strategy in Côte d’Ivoire

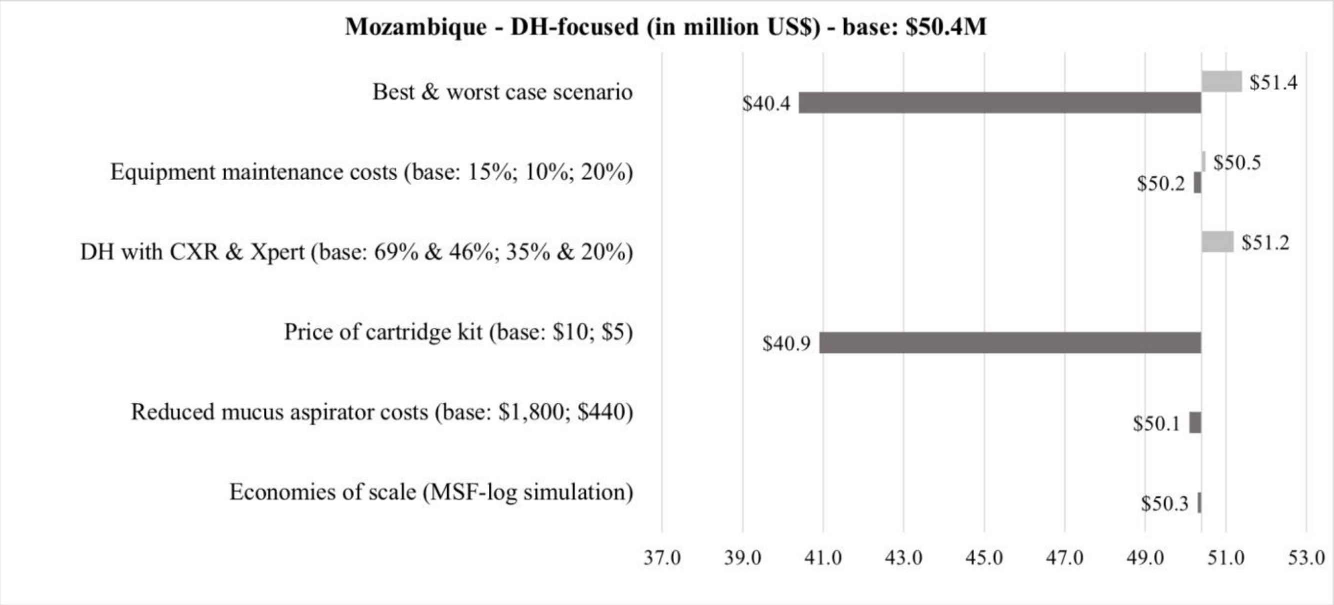


**Figure 27**. Tornado diagram on findings from sensitivity analysis on total scale-up costs of the DH-focused strategy in Mozambique

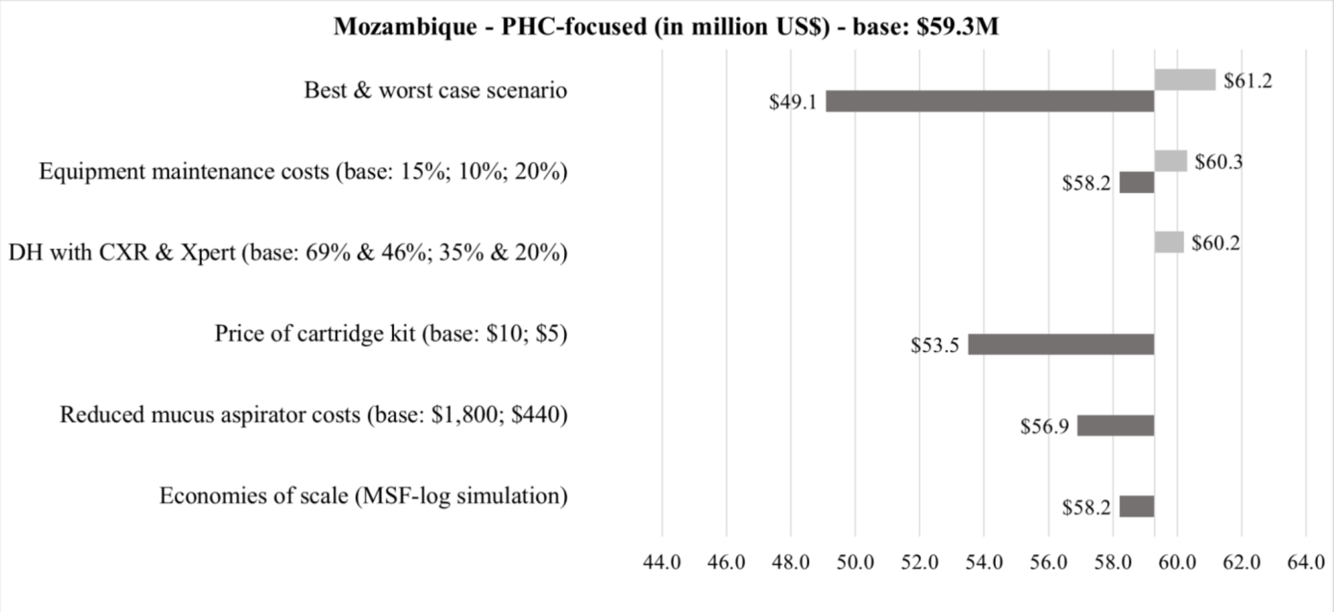


**Figure 28**. Tornado diagram on findings from sensitivity analysis on total scale-up costs of the PHC-focused strategy in Mozambique

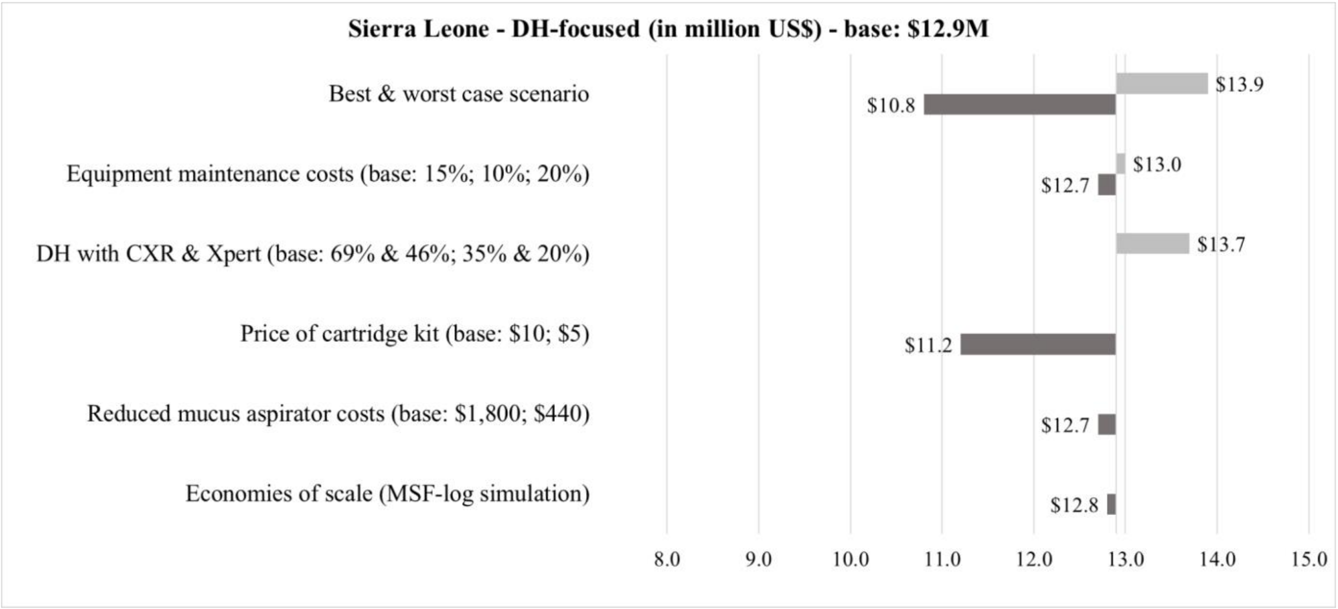


**Figure 29**. Tornado diagram on findings from sensitivity analysis on total scale-up costs of the DH-focused strategy in Sierra Leone

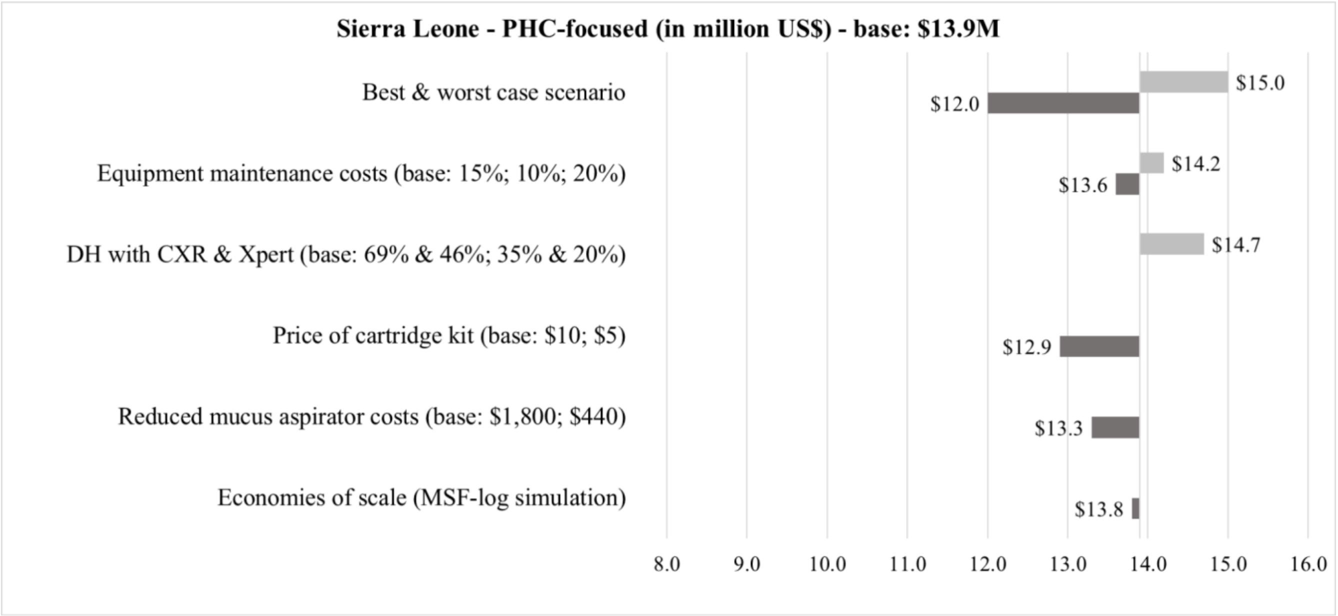


**Figure 30**. Tornado diagram on findings from sensitivity analysis on total scale-up costs of the PHC-focused strategy in Sierra Leone

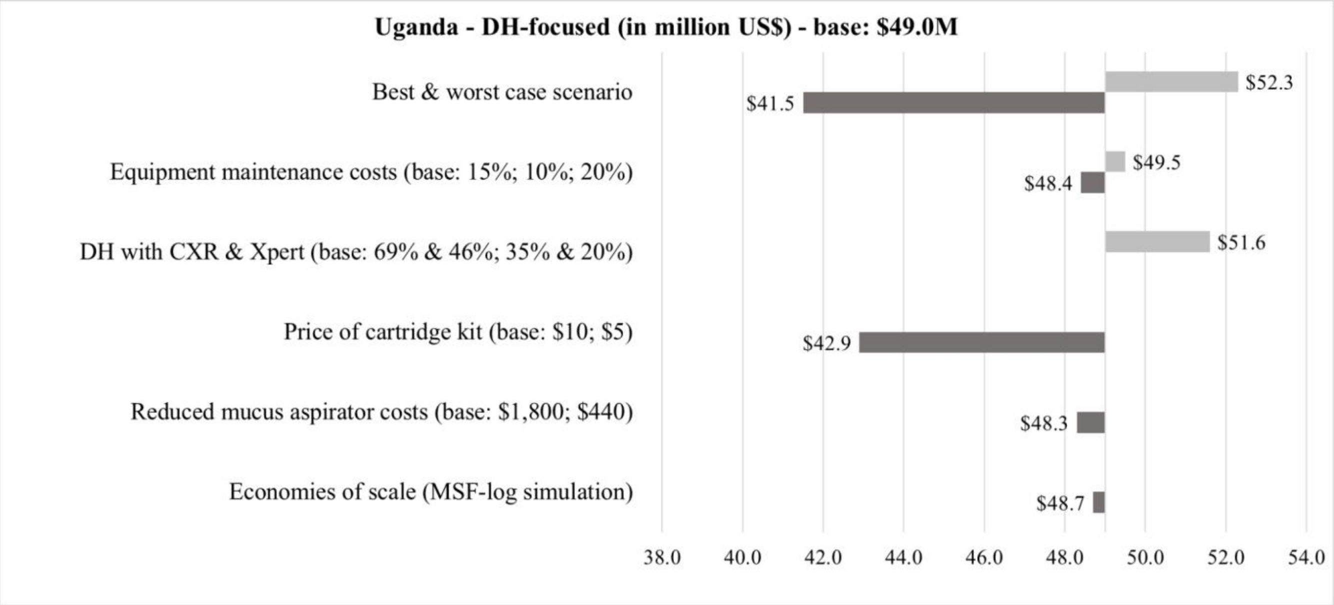


**Figure 31**. Tornado diagram on findings from sensitivity analysis on total scale-up costs of the DH-focused strategy in Uganda

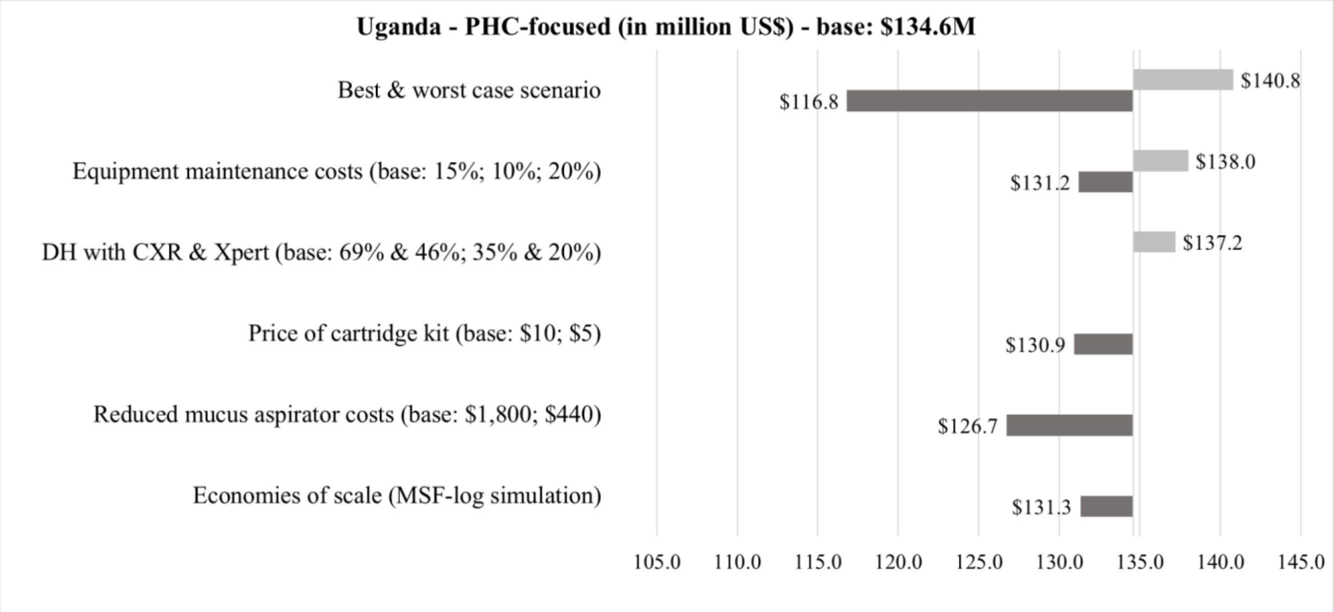


**Figure 32**. Tornado diagram on findings from sensitivity analysis on total scale-up costs of the PHC-focused strategy in Uganda

### Ethics committees and review boards for the study

**Table 15**. Ethics committees and review boards for the study

| | |
|---|---|
| Global | WHO Research Ethics Review Committee (WHO ERC) |
| Sponsor | Inserm - Comité d'évaluation éthique de l'Inserm (CEEI-IRB) |
| Cambodia | National Ethics Committee for Health Research |
| Cameroon | National Ethics Committee on Human Health Research |
| Côte d’Ivoire | Comité National d'Ethique de la Recherche |
| Mozambique | Comité Institucional de Bioética para Saude do INS<br>Comité Nacional de Bioética para a Saude (CNBS) |
| Uganda | Mbarara University of Science and Technology REC<br>Uganda National Council for Science and Technology |
| Sierra Leone | Sierra Leone Ethics and Scientific review committee |

### References

1 Wobudeya E, Nanfuka M, Huyen Ton Nu Nguyet M, *et al.* Impact of decentralizing childhood tuberculosis diagnosis at primary health center and district hospital level - A pre-post study in six high tuberculosis burden countries. .

2 Dodd PJ. HEdtree: A Package For Decision Tree Modelling. 2023 https://github.com/petedodd/HEdtree.

3 Dodd PJ, Gardiner E, Coghlan R, Seddon JA. Burden of childhood tuberculosis in 22 high-burden countries: a mathematical modelling study. *Lancet Glob Health* 2014; **2**: e453–9.

4 Dodd PJ, Sismanidis C, Seddon JA. Global burden of drug-resistant tuberculosis in children: a mathematical modelling study. *Lancet Infect Dis* 2016; **16**: 1193–201.

5 Dodd PJ, Yuen CM, Sismanidis C, Seddon JA, Jenkins HE. The global burden of tuberculosis mortality in children: a mathematical modelling study. *Lancet Glob Health* 2017; **5**: e898–906.

6 Dodd PJ, Prendergast AJ, Beecroft C, Kampmann B, Seddon JA. The impact of HIV and antiretroviral therapy on TB risk in children: a systematic review and meta-analysis. *Thorax* 2017; **72**: 559–75.

7 Disability-adjusted life years (DALYs). https://www.who.int/data/gho/indicator-metadata-registry/imr-details/158 (accessed April 20, 2023).

8 Wilkinson T, Sculpher MJ, Claxton K, *et al.* The International Decision Support Initiative Reference Case for Economic Evaluation: An Aid to Thought. *Value Health* 2016; **19**: 921–8.

9 Tan-Torres Edejer T, Baltussen R, Adam T, *et al.*, editors. Making choices in health: WHO guide to cost-effectiveness. World Health Organization.

10 Chi Y-L, Blecher M, Chalkidou K, *et al.* What next after GDP-based cost-effectiveness thresholds? *Gates Open Res* 2020; **4**: 176.

11 Bertram MY, Lauer JA, De Joncheere K, *et al.* Cost-effectiveness thresholds: pros and cons. *Bull World Health Organ* 2016; **94**: 925–30.

12 Cameron D, Ubels J, Norström F. On what basis are medical cost-effectiveness thresholds set? Clashing opinions and an absence of data: a systematic review. *Glob Health Action* 2018; **11**: 1447828.

13 Ochalek J, Lomas J, Claxton K. Estimating health opportunity costs in low-income and middle-income countries: a novel approach and evidence from cross-country data. *BMJ Glob Health* 2018; **3**: e000964.

14 Woods B, Revill P, Sculpher M, Claxton K. Country-Level Cost-Effectiveness Thresholds: Initial Estimates and the Need for Further Research. *Value Health* 2016; **19**:

929–35.

15 Kay AW, González Fernández L, Takwoingi Y, *et al.* Xpert MTB/RIF and Xpert MTB/RIF Ultra assays for active tuberculosis and rifampicin resistance in children. *Cochrane Database Syst Rev* 2020; **8**: CD013359.

16 Marais BJ, Gie RP, Hesseling AC, *et al.* A refined symptom-based approach to diagnose pulmonary tuberculosis in children. *Pediatrics* 2006; **118**: e1350–9.

17 Jenkins HE, Yuen CM, Rodriguez CA, *et al.* Mortality in children diagnosed with tuberculosis: a systematic review and meta-analysis. *Lancet Infect Dis* 2017; **17**: 285–95.

18 South A, Dicko A, Herringer M, *et al.* A rapid and reproducible picture of open access health facility data in Africa to support the COVID-19 response. *Wellcome Open Res* 2020; **5**: 157.
